# Perceived indoor temperature extremes are associated with sleep health among women in the United States

**DOI:** 10.1101/2025.09.12.25335650

**Authors:** Symielle A. Gaston, Dayna T. Neo, W. Braxton Jackson, Dale P. Sandler, Chandra L. Jackson

**Affiliations:** Epidemiology Branch, National Institute of Environmental Health Sciences, National Institutes of Health, Research Triangle Park, NC, USA; DLH LLC, Bethesda, MD, USA; Division of Intramural Research, National Institute on Minority Health and Health Disparities, National Institutes of Health, Bethesda, MD, USA

**Keywords:** hot temperature, cold temperature, sleep, women, health status disparities, race factors

## Abstract

**Introduction:** Indoor temperature extremes contribute to thermal discomfort and can threaten sleep health. Few studies have investigated indoor temperature-sleep associations, especially across differentially-exposed sociodemographic groups.

**Methods:** Using cross-sectional Sister Study data (2017-2019), we estimated associations between perceived sleep-disrupting indoor temperature extremes (SDITE) and sleep dimensions. Eligible women reported frequent vs. infrequent SDITE considered ‘too hot’ or ‘too cold’ and sleep dimensions (sleep duration; Pittsburgh Sleep Quality Index-derived sleep disturbances; healthcare professional-diagnosed sleep apnea). Adjusting for trouble sleeping for reasons other than temperature, sociodemographic characteristics, and clinical characteristics, we used Poisson regression with robust variance to estimate prevalence ratios (PRs) and 95% confidence intervals (CIs).

**Results:** Among 33,545 women (mean±SD age = 67±8.5 years), 90% self-identified as non-Hispanic White, 6.6% as non-Hispanic Black, and 3.7% as Latina, and 27% reported an annual household income (AHI) ≤$49,999 while 37% reported an AHI $50,000-$99,999 and 37% reported an AHI ≥$100,000. Prevalence of ‘too hot’ SDITE was highest among non-Hispanic Black women (15% vs. 9.0% overall) and women with an AHI ≥$100,000 (11%); ‘too cold’ SDITE prevalence was highest among Latina women (3.8% vs. 1.4% overall) and women with an AHI ≤$49,999 (2.2%). ‘Too hot’ and ‘too cold’ SDITE were consistently associated with long sleep onset latency (LSOL) (PR_hot_=1.89 [1.75-2.04] and PR_cold_=1.99 [1.70-2.33]) and daytime dysfunction (PR_hot_=1.76 [1.23-2.51] and PR_cold_=4.69 [2.83-7.76]).

**Conclusions:** Perceived SDITE were associated with insomnia symptoms and may contribute to sleep disparities given the higher burdens often observed among Black women, Latina women, and women with lower annual household incomes.

## INTRODUCTION

Due to the long-standing under-recognition of sleep as an essential pillar of health, a global call to action was issued to prioritize sleep health.^1^ Fortunately, sleep health is increasingly recognized as a public health problem in need of immediate attention given the importance of sleep as a likely determinant of overall health and well-being.^2^ Despite the critical role of sleep in relation to many costly and potentially-devasting health outcomes,^3^ one in three adults in the United States (US) do not achieve the recommendation of at least seven hours of quality sleep.^4,5^ Sleep disorders such as insomnia are also prevalent.^2^ Furthermore, certain segments of the population are even more likely to experience poorer sleep health than their counterparts.^5–9^ For instance, men compared to women report more established risk factors for obstructive sleep apnea,^10^ and women more often report certain sleep disturbances (e.g., insomnia symptoms).^11^ In the US, most of the other racial and ethnic groups exhibit poorer sleep health compared to their non-Hispanic White counterparts.^8^ These sleep disparities as well as general sleep disturbances in the overall population are influenced by modifiable environmental factors, such as light at night, noise, and temperatures that impede thermoregulation.^12,13^ One study suggests noise and ambient temperatures are perceived as the most influential factors.^14^ Relatedly, increases in adverse environmental conditions such as temperature extremes are now considered a dire threat to human sleep health, globally.^15–18^ Therefore, it is important to investigate sleep in the context of long-term changes in weather patterns.^19,20^ Moreover, sleep quality is expected to inequitably decrease among populations with higher susceptibility to temperature extremes and temperature-related sleep disturbances (e.g., women, lower income populations),^16,18^ thereby widening existing sleep disparities.

Sleep disparities may be addressed in several ways, including by evaluating the immediate conditions (i.e., the bedroom/indoor environment) in which people sleep.^21^ Suboptimal outdoor temperatures that are either too hot or too cold have been associated with unfavorable (i.e., too short or too long) sleep duration and poor sleep quality.^13,22–24^ These relationships may be explained by both cold and hot temperatures likely altering sleep architecture (e.g., fragmented and less physiologically-restorative slow wave sleep).^13^ Distinctively, indoor temperatures are more directly experienced, have been shown to be both colder and warmer than outdoor ambient temperatures,^25,26^ and, according to a recent review, have had stronger associations with sleep than outdoor temperatures.^22^ However, empirical studies of indoor temperature extremes and multiple sleep dimensions are sparse, and – of these few studies – most include populations outside of the US.^22,23,27–40^ Furthermore, while most findings suggest indoor temperatures considered too hot are related to more disrupted sleep,^22,26–28,30–32,37^ studies of cold indoor temperatures are fewer and yielded mixed results.^23,30,34,35,38,41^ Moreover, indoor temperature extremes are especially important to consider when temperature control is inadequate, such as in residences that have limited to no capacity to control heating and cooling^13,42^ Investigations of disparities in associations between indoor temperatures and sleep health are sparse although socioeconomic burdens – that disproportionately impact socially-relegated groups – can lead to, for instance, a routine inability to meet basic household energy (heating/cooling) needs.^16–19,42,43^

To address gaps in the current literature, we sought to determine the prevalence of perceived sleep-disrupting indoor temperature extremes (SDITE), to characterize multiple sleep dimensions by reports of perceived SDITE, and to determine whether relationships between perceived SDITE and sleep dimensions differ by race and ethnicity as well as socioeconomic status (SES) among a multiethnic cohort of US women. Findings from this large cohort of women is important because women tend to have poorer sleep quality than men at all temperatures and because women have been shown to be physiologically more sensitive to temperatures during sleep than men.^44^ Additionally, perceived and observed temperatures are related, with one study suggesting almost three times the odds of perceiving temperature in a higher category such as ‘hot’ or ‘very hot’ with each one degree Celsius increase in measured temperature.^41^ Perceived temperatures also likely reflect subjective, personalized thermal comfort,^32,38,45^ which is important but irregularly assessed in studies of temperature and sleep.^13^ We hypothesized that perceptions of both too hot and too cold SDITE, expected to reflect actual indoor temperatures, are more frequently reported by minoritized racial and ethnic groups compared to non-Hispanic White women and by women with lower compared to higher annual household incomes (AHIs). We also hypothesized that perceptions of both extremely hot and extremely cold SDITE are associated with poorer subjective sleep duration and quality. Further, we hypothesized that relationships are stronger among historically socially-relegated groups,^8^ including racially and ethnically minoritized women compared to non-Hispanic White women and women with lower compared to higher AHIs. Moreover, relationships may vary by geographical region of residence due to factors like geographic variation in both access to central air conditioning and sleep.^5,46^ Relationships may also vary by menopausal status due to differences related to biological and hormonal changes that can affect both perceived temperature as well as thermal comfort and sleep quality among women.^47,48^ Therefore, as exploratory aims, we also sought to determine whether associations varied by region of residence and menopausal status.

## METHODS

### Data source: The Sister Study

We used data from the Sister Study, an ongoing cohort study of 50,884 US women (Release 11.1 with follow-up until September 30, 2021). Details about the Sister Study are described elsewhere.^49^ Briefly, eligible women aged 35 to 74 years who had a biological sister with a prior diagnosis of breast cancer but who were themselves without a breast cancer diagnosis were enrolled between 2003-2009. Participants completed home visits, self-administered questionnaires, computer assisted telephone interviews, and computer-assisted web interviews at enrollment. Brief follow-up interviews are completed annually, and detailed follow-up data collection occurs approximately every 2-3 years. All participants provided written informed consent. The National Institute of Environmental Health Sciences Institutional Review Board approved and oversees Sister Study protocols. Review is waived for this analysis of de-identified, secondary data.

### Study population

Data on perceived SDITE were collected in a follow-up interview during 2017-2019. Therefore, only eligible participants who completed the follow-up interview were included in the current cross-sectional analysis. As illustrated in Supplemental Figure 1, we applied the following additional exclusions in a stepwise manner: withdrew from the study (n=5); did not complete the follow-up questionnaire (n=10,168); missing data for perceived SDITE (n=5,459); missing data for potential modifiers – race and ethnicity (n=7) or AHI (n=282); self-identified as Asian, American Indian/Alaska Native, multiracial, Native Hawaiian/Pacific Islander, or some other race and ethnicity [due to small sample size within groups and within-group heterogeneity if groups were combined (n=794)]; and underweight (body mass index <18.5 kg/m^2^) [due to small sample size and its previously observed association with morbidity and mortality (n=624)],^50^ which could bias results. The final analytic sample comprised 33,545 eligible participants. Comparing characteristics at enrollment among ineligible to eligible participants (Supplemental Table 1), ineligible participants were less likely to self-identify as non-Hispanic [NH] White; had lower SES; and were more likely to not be married or living as though married, reside in the South or Puerto Rico, have obesity, and be postmenopausal.

### Exposure assessment: Perceived sleep-disrupting indoor temperature extremes (SDITE)

We captured frequency of perceived SDITE. Prior literature demonstrates that perceived or subjective assessments of temperature are strongly associated with measured temperature and that perceived comfortable temperatures align with the optimal objective temperature for sleep onset and sleep maintenance.^32,38,41,45^ In an abbreviated version of the Pittsburgh Sleep Quality Index (PSQI),^51^ participants used a 4-point Likert scale ranging from ‘not during the past month’ to ‘three or more times/week’ to respond to the following questions, “*During the past month, how often have you had trouble sleeping because you…*” (1) *“felt too hot”* and (2) *“felt too cold”*. To capture frequently perceiving SDITE, we dichotomized (1) ‘too hot’ as yes (reports of “*had trouble sleeping because you felt too hot”* ≥3 times/week) vs. no (reports of both “*had trouble sleeping because you felt too hot’ 0 to <3 times/week”* and “*had trouble sleeping because you felt too cold”* 0 to <3 times/week) and (2) ‘too cold’ as yes (reports of “*had trouble sleeping because you felt too cold”* ≥3 times/week) vs. no (reports of both “*had trouble sleeping because you felt too hot’ 0 to <3 times/week”* and “*had trouble sleeping because you felt too cold”* 0 to <3 times/week). To isolate each type of perceived SDITE (i.e., ‘too hot’ and ‘too cold’ separately), participants who endorsed trouble sleeping due to being both ‘too hot’ ≥3 times/week and ‘too cold’ ≥3 times/week (n=438, 1.3%) were not included in analyses.

### Outcome assessment: Multiple sleep health dimensions

Participants either reported (1) daily bed and wake times on workdays/weekdays and on non-workdays/weekends or (2) average sleep duration in the past year, which we used to derive average weekly duration over 7-days. Average values of ≤2 hours or ≥23 hours were considered implausible; therefore, participants with these values (n=404) were excluded from sleep duration analyses. We then categorized sleep duration as short (<7 hours), recommended (7-9 hours), and long (>9 hours) based on recommendations by the National Sleep Foundation.^52^ We also assessed the following PSQI-derived individual sleep disturbances in the past month,^51^ separately: long sleep onset latency (LSOL; taking ≥30 minutes to fall asleep), poor sleep maintenance (wake up in the middle of the night or early morning), insomnia symptoms (either LSOL or poor sleep maintenance), sleep medication use, and daytime dysfunction (i.e., trouble staying awake while driving, eating meals, or engaging in social activity). Each were defined as yes (≥3 times/week) vs. no (<3 times/week) rather than with continuous scores to identify meaningful categories reflecting potential sleep problems that could serve as possible targets for intervention. Lastly, participants reported whether they were (yes vs. no) ever diagnosed by a healthcare professional with and currently have sleep apnea.

### Potential confounders

We chose *a priori* potential confounders based on prior literature.^5,8,53–56^ Sociodemographic characteristics included age (years) and age^2^, self-identified race and ethnicity (Hispanic/Latina, NH Black, NH White) based on US Office of Management and Budget categories,^57^ AHI (<$20,000-$49,999; $50,000-$99,999; ≥$100,000), marital status (as a proxy for likelihood of bed sharing: married or living as though married; never married, divorced, widowed, or separated), and region of residence (Northeast, Midwest, South, West, Puerto Rico). Clinical characteristics included body mass index category (recommended [18.5 – 24.9 kg/m^2^], overweight [25.0 – 29.9 kg/m^2^], obesity [≥30.0 kg/m^2^) and menopausal status (premenopausal, postmenopausal). We also included participant reports on the PSQI of whether (yes vs. no) they had trouble sleeping for any other reason ≥ 3 times/week during the past month.^51^ These other reasons included *getting up to use the bathroom*; *unable to breathe comfortably*; *cough or snore loudly*; *bad dreams*; *pain*; or *other reason(s)*. Additionally, any other reasons could include *unable to fall asleep within 30 minutes* or *waking in the middle of the night or early morning* when considering the following sleep health dimensions as outcomes: weekly sleep duration, sleep medication use, daytime dysfunction, and healthcare professional diagnosed (HPD) sleep apnea.

### Potential modifiers

Race and ethnicity as well as AHI were potential modifiers in the main analysis. As an exploratory analysis, we also considered region of residence and menopausal status as potential modifiers since prior evidence suggests that temperature extremes, extreme temperature mitigation resources (e.g., air conditioning), and sleep vary by US region of residence^5,46,58^ and that thermal comfort and sleep vary by menopausal status.^47,48^

### Statistical analysis

We calculated descriptive statistics as means ± standard deviations for continuous variables and as frequencies and percentages for categorical variables in the total population as well as by categories of perceived SDITE. We also further stratified descriptive statistics by race and ethnicity, AHI, region of residence, and menopausal status. Using Chi-square or ANOVA, we tested for differences in SDITE and other characteristics by strata (e.g., by racial and ethnic group). Using Poisson regression with robust variance estimation, we calculated prevalence ratios (PRs) and 95% confidence intervals (CIs) for indoor ‘too hot’ and ‘too cold’ vs. ‘infrequent temperature extremes’ in relation to each poor sleep characteristic, separately. In the total population, potential confounders were added to models in a stepwise manner. Model 1 was adjusted for whether participants reported trouble sleeping for any other reasons than indoor temperature extremes. Model 2 was additionally adjusted for age, age^2^, race and ethnicity, AHI, and region of residence. Lastly, Model 3 was adjusted for Model 2 confounders as well as body mass index category and menopausal status. We then assessed effect modification on the multiplicative scale by race and ethnicity, AHI, region of residence, and menopausal status, separately by inserting temperature extreme *potential modifier interaction terms in fully adjusted models and examining Wald p-values. We then reported results stratified by the potential modifiers. The false discovery rate (FDR) was used for multiple comparisons correction. To assess effect modification on the additive scale, we estimated the relative excess risk due to interaction (RERI) and 95% CIs based on standard errors obtained using the Hosmer and Lemeshow method.^59^ In a sensitivity analysis among only participants who reported trouble sleeping for any reason other than SDITE, we calculated descriptive statistics and re-estimated all fully-adjusted, FDR-corrected models. All analyses were conducted using SAS, 9.4 (Cary, NC), and a two-sided p-value of 0.05 determined statistical significance. Figures were created in Python version 3.11.12 within the Spyder Integrated Development Environment (IDE) version 6.0.7.

## RESULTS

### Study population characteristics

Among 33,545 eligible participants with a mean ± standard deviation age of 67±8.5 years, 3.7% identified as Latina, 6.6% as NH Black, and 90% as NH White (Table 1). Most attained a Bachelor’s degree or higher (55%), had an AHI of $50,000 or more (74%), and were currently married or living as married (68%). The highest proportion of participants resided in the South (33%) while the fewest resided in Puerto Rico (1.1%), and most were postmenopausal (96%). While 88% of participants reported infrequent SDITE, 9.0% reported frequently perceiving ‘too hot’ SDITE, and 1.4% reported frequently perceiving ‘too cold’ SDITE (1.3% reported both and were excluded from analyses). Additionally, 71% of participants reported frequent trouble sleeping for any other reason than temperature. Most participants reported their average weekly sleep duration within the recommended range of 7 to 9 hours (73%) while 11% reported short sleep and 15% reported long sleep. Prevalence of LSOL was 13%, poor sleep maintenance was 40%, insomnia symptoms was 43%, sleep medication use was 19%, daytime dysfunction was <1%, and 10% reported HPD sleep apnea.

**Table 1.**
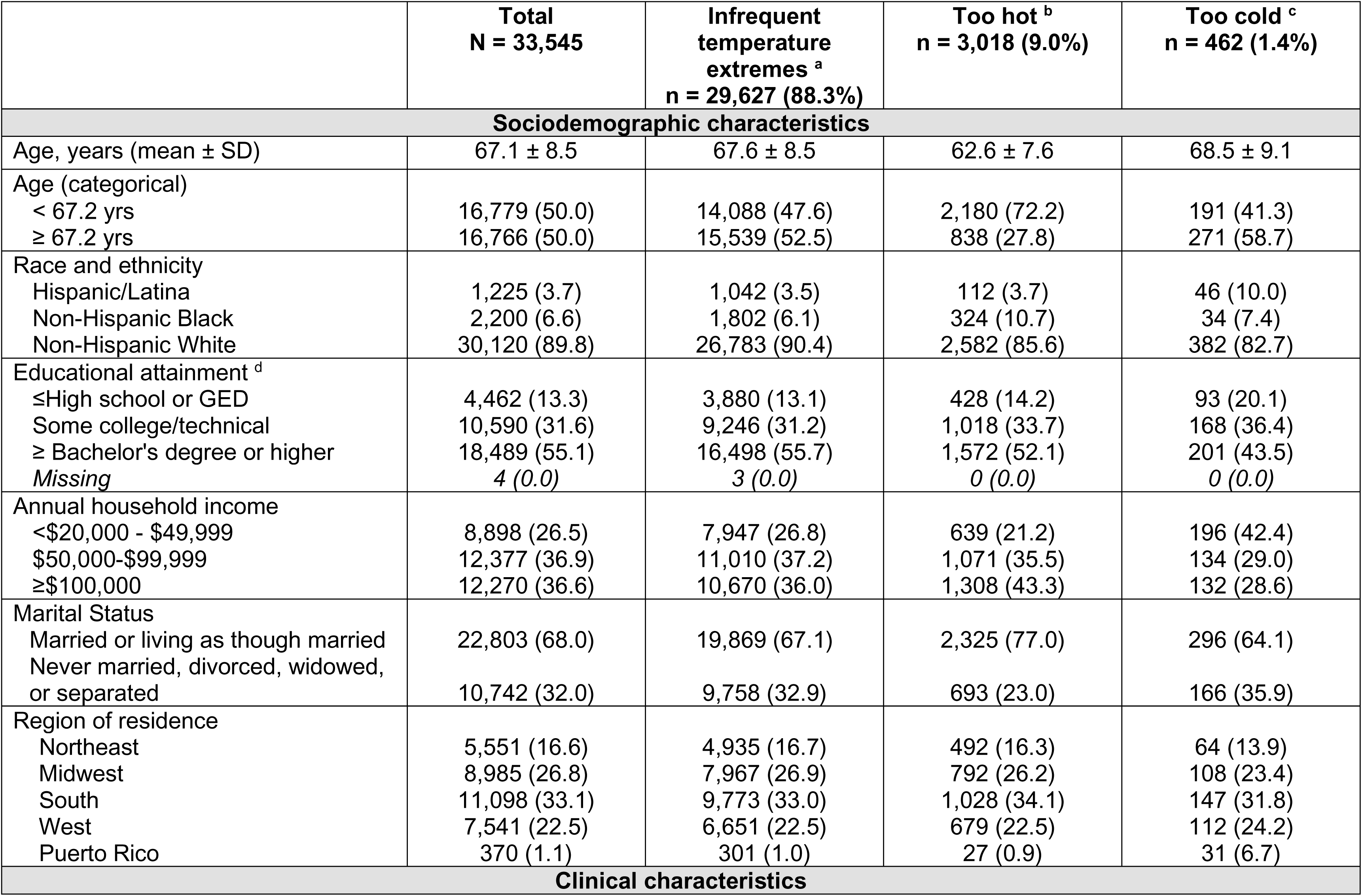
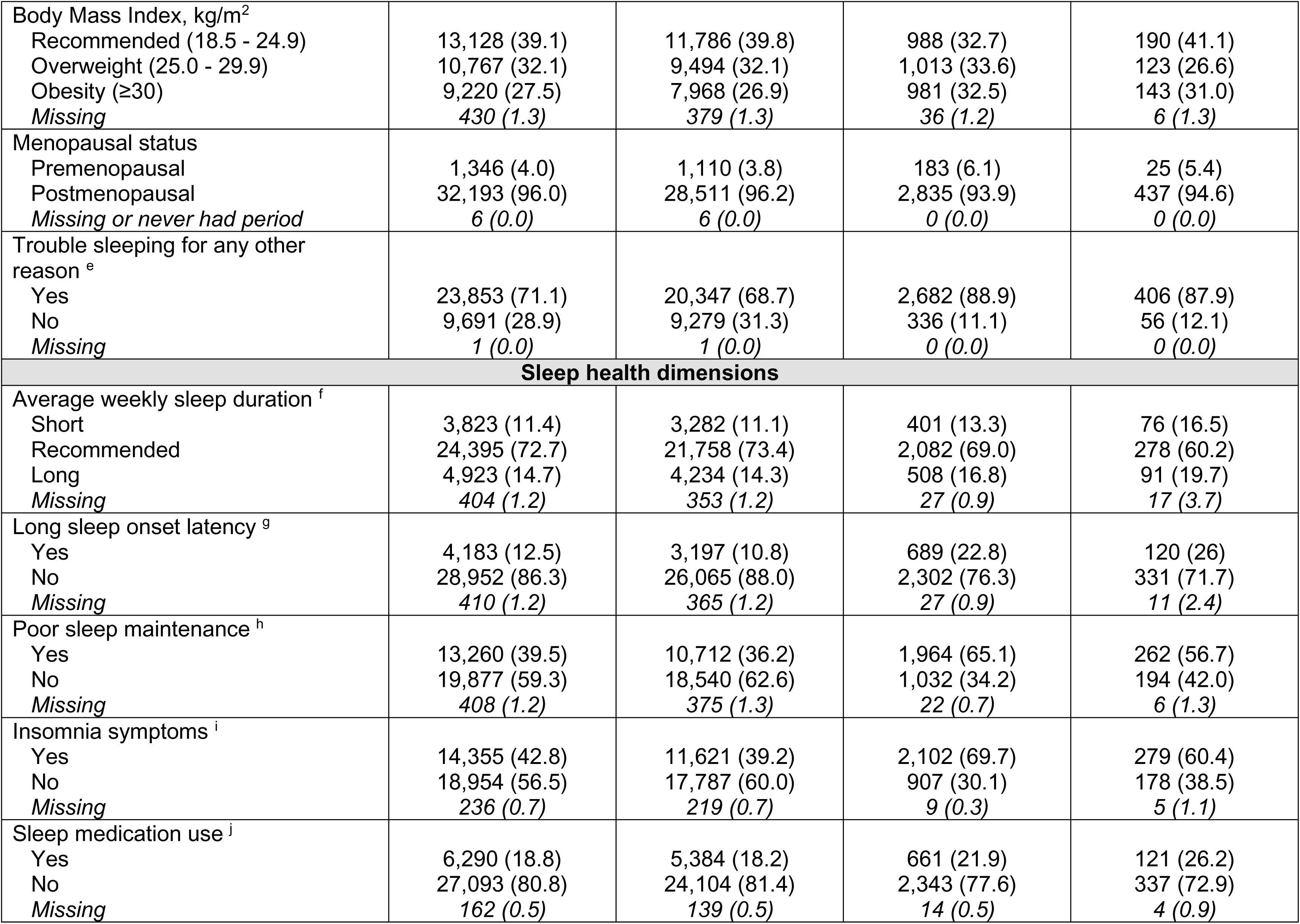
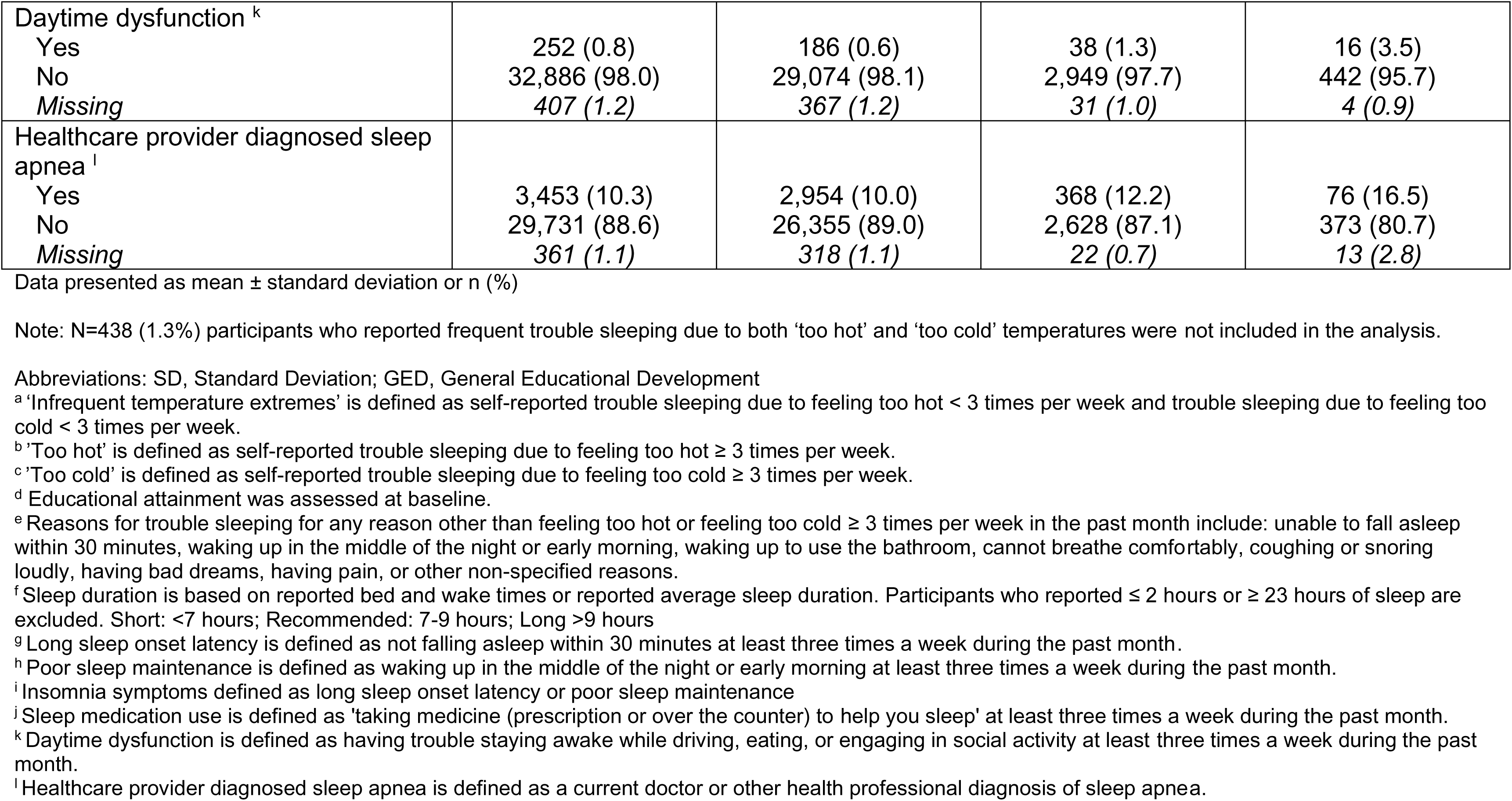
Sociodemographic, clinical, and sleep health characteristics among eligible participants, overall and by perceived indoor temperature extremes, Sister Study, 2017-2019, N=33,545.

|  | **Total**<br>**N = 33,545** | **Infrequent**<br>**temperature**<br>**extremes <sup>a</sup>**<br>**n = 29,627 (88.3%)** | **Too hot <sup>b</sup>**<br>**n = 3,018 (9.0%)** | **Too cold <sup>c</sup>**<br>**n = 462 (1.4%)** |
| --- | --- | --- | --- | --- |
| **Sociodemographic characteristics** |  |  |  |  |
| Age, years (mean ± SD) | 67.1 ± 8.5 | 67.6 ± 8.5 | 62.6 ± 7.6 | 68.5 ± 9.1 |
| Age (categorical) |  |  |  |  |
| < 67.2 yrs | 16,779 (50.0) | 14,088 (47.6) | 2,180 (72.2) | 191 (41.3) |
| ≥ 67.2 yrs | 16,766 (50.0) | 15,539 (52.5) | 838 (27.8) | 271 (58.7) |
| Race and ethnicity |  |  |  |  |
| Hispanic/Latina | 1,225 (3.7) | 1,042 (3.5) | 112 (3.7) | 46 (10.0) |
| Non-Hispanic Black | 2,200 (6.6) | 1,802 (6.1) | 324 (10.7) | 34 (7.4) |
| Non-Hispanic White | 30,120 (89.8) | 26,783 (90.4) | 2,582 (85.6) | 382 (82.7) |
| Educational attainment <sup>d</sup> |  |  |  |  |
| ≤High school or GED | 4,462 (13.3) | 3,880 (13.1) | 428 (14.2) | 93 (20.1) |
| Some college/technical | 10,590 (31.6) | 9,246 (31.2) | 1,018 (33.7) | 168 (36.4) |
| ≥ Bachelor's degree or higher | 18,489 (55.1) | 16,498 (55.7) | 1,572 (52.1) | 201 (43.5) |
| Missing | 4 (0.0) | 3 (0.0) | 0 (0.0) | 0 (0.0) |
| Annual household income |  |  |  |  |
| <\$20,000 - \$49,999 | 8,898 (26.5) | 7,947 (26.8) | 639 (21.2) | 196 (42.4) |
| \$50,000-\$99,999 | 12,377 (36.9) | 11,010 (37.2) | 1,071 (35.5) | 134 (29.0) |
| ≥\$100,000 | 12,270 (36.6) | 10,670 (36.0) | 1,308 (43.3) | 132 (28.6) |
| Marital Status |  |  |  |  |
| Married or living as though married | 22,803 (68.0) | 19,869 (67.1) | 2,325 (77.0) | 296 (64.1) |
| Never married, divorced, widowed, or separated | 10,742 (32.0) | 9,758 (32.9) | 693 (23.0) | 166 (35.9) |
| Region of residence |  |  |  |  |
| Northeast | 5,551 (16.6) | 4,935 (16.7) | 492 (16.3) | 64 (13.9) |
| Midwest | 8,985 (26.8) | 7,967 (26.9) | 792 (26.2) | 108 (23.4) |
| South | 11,098 (33.1) | 9,773 (33.0) | 1,028 (34.1) | 147 (31.8) |
| West | 7,541 (22.5) | 6,651 (22.5) | 679 (22.5) | 112 (24.2) |
| Puerto Rico | 370 (1.1) | 301 (1.0) | 27 (0.9) | 31 (6.7) |
| **Clinical characteristics** |  |  |  |  |
| Body Mass Index, kg/m<sup>2</sup><br>Recommended (18.5 - 24.9)<br>Overweight (25.0 - 29.9)<br>Obesity (≥30)<br>*Missing* | 13,128 (39.1)<br>10,767 (32.1)<br>9,220 (27.5)<br>430 (1.3) | 11,786 (39.8)<br>9,494 (32.1)<br>7,968 (26.9)<br>379 (1.3) | 988 (32.7)<br>1,013 (33.6)<br>981 (32.5)<br>36 (1.2) | 190 (41.1)<br>123 (26.6)<br>143 (31.0)<br>6 (1.3) |
| Menopausal status<br>Premenopausal<br>Postmenopausal<br>*Missing or never had period* | 1,346 (4.0)<br>32,193 (96.0)<br>6 (0.0) | 1,110 (3.8)<br>28,511 (96.2)<br>6 (0.0) | 183 (6.1)<br>2,835 (93.9)<br>0 (0.0) | 25 (5.4)<br>437 (94.6)<br>0 (0.0) |
| Trouble sleeping for any other reason <sup>e</sup><br>Yes<br>No<br>*Missing* | 23,853 (71.1)<br>9,691 (28.9)<br>1 (0.0) | 20,347 (68.7)<br>9,279 (31.3)<br>1 (0.0) | 2,682 (88.9)<br>336 (11.1)<br>0 (0.0) | 406 (87.9)<br>56 (12.1)<br>0 (0.0) |
| **Sleep health dimensions** |  |  |  |  |
| Average weekly sleep duration <sup>f</sup><br>Short<br>Recommended<br>Long<br>*Missing* | 3,823 (11.4)<br>24,395 (72.7)<br>4,923 (14.7)<br>404 (1.2) | 3,282 (11.1)<br>21,758 (73.4)<br>4,234 (14.3)<br>353 (1.2) | 401 (13.3)<br>2,082 (69.0)<br>508 (16.8)<br>27 (0.9) | 76 (16.5)<br>278 (60.2)<br>91 (19.7)<br>17 (3.7) |
| Long sleep onset latency <sup>g</sup><br>Yes<br>No<br>*Missing* | 4,183 (12.5)<br>28,952 (86.3)<br>410 (1.2) | 3,197 (10.8)<br>26,065 (88.0)<br>365 (1.2) | 689 (22.8)<br>2,302 (76.3)<br>27 (0.9) | 120 (26)<br>331 (71.7)<br>11 (2.4) |
| Poor sleep maintenance <sup>h</sup><br>Yes<br>No<br>*Missing* | 13,260 (39.5)<br>19,877 (59.3)<br>408 (1.2) | 10,712 (36.2)<br>18,540 (62.6)<br>375 (1.3) | 1,964 (65.1)<br>1,032 (34.2)<br>22 (0.7) | 262 (56.7)<br>194 (42.0)<br>6 (1.3) |
| Insomnia symptoms <sup>i</sup><br>Yes<br>No<br>*Missing* | 14,355 (42.8)<br>18,954 (56.5)<br>236 (0.7) | 11,621 (39.2)<br>17,787 (60.0)<br>219 (0.7) | 2,102 (69.7)<br>907 (30.1)<br>9 (0.3) | 279 (60.4)<br>178 (38.5)<br>5 (1.1) |
| Sleep medication use <sup>j</sup><br>Yes<br>No<br>*Missing* | 6,290 (18.8)<br>27,093 (80.8)<br>162 (0.5) | 5,384 (18.2)<br>24,104 (81.4)<br>139 (0.5) | 661 (21.9)<br>2,343 (77.6)<br>14 (0.5) | 121 (26.2)<br>337 (72.9)<br>4 (0.9) |
| Daytime dysfunction <sup>k</sup> |  |  |  |  |
| Yes | 252 (0.8) | 186 (0.6) | 38 (1.3) | 16 (3.5) |
| No | 32,886 (98.0) | 29,074 (98.1) | 2,949 (97.7) | 442 (95.7) |
| Missing | 407 (1.2) | 367 (1.2) | 31 (1.0) | 4 (0.9) |
| Healthcare provider diagnosed sleep apnea <sup>l</sup> |  |  |  |  |
| Yes | 3,453 (10.3) | 2,954 (10.0) | 368 (12.2) | 76 (16.5) |
| No | 29,731 (88.6) | 26,355 (89.0) | 2,628 (87.1) | 373 (80.7) |
| Missing | 361 (1.1) | 318 (1.1) | 22 (0.7) | 13 (2.8) |

Data presented as mean ± standard deviation or n (%)
Note: N=438 (1.3%) participants who reported frequent trouble sleeping due to both 'too hot' and 'too cold' temperatures were not included in the analysis.
Abbreviations: SD, Standard Deviation; GED, General Educational Development
<sup>a</sup> 'Infrequent temperature extremes' is defined as self-reported trouble sleeping due to feeling too hot < 3 times per week and trouble sleeping due to feeling too cold < 3 times per week.
<sup>b</sup> 'Too hot' is defined as self-reported trouble sleeping due to feeling too hot ≥ 3 times per week.
<sup>c</sup> 'Too cold' is defined as self-reported trouble sleeping due to feeling too cold ≥ 3 times per week.
<sup>d</sup> Educational attainment was assessed at baseline.
<sup>e</sup> Reasons for trouble sleeping for any reason other than feeling too hot or feeling too cold ≥ 3 times per week in the past month include: unable to fall asleep within 30 minutes, waking up in the middle of the night or early morning, waking up to use the bathroom, cannot breathe comfortably, coughing or snoring loudly, having bad dreams, having pain, or other non-specified reasons.
<sup>f</sup> Sleep duration is based on reported bed and wake times or reported average sleep duration. Participants who reported ≤ 2 hours or ≥ 23 hours of sleep are excluded. Short: <7 hours; Recommended: 7-9 hours; Long >9 hours

By race and ethnicity, the prevalence of frequently perceiving ‘too hot’ SDITE was highest among NH Black women (15% vs. 9.1% Latina and 8.6% NH White; p<0.05); reports of frequently perceiving ‘too cold’ SDITE were highest among Latina participants (3.8% vs.1.6% NH Black and 1.3% NH White; p<0.05; Supplemental Table 2). Short sleep prevalence was highest among NH Black participants; long sleep prevalence was highest among Latina participants; LSOL was more prevalent among Latina followed by NH Black and NH White participants; poor sleep maintenance, insomnia symptoms, and sleep medication use were highest among NH White participants; and HPD sleep apnea prevalence was highest among NH Black participants (all p<0.05). Latina and NH Black participants reported lower AHIs compared with NH White participants (p<0.05). Thirty percent of Latina participants resided in Puerto Rico while 0% of NH Black and NH White participants resided in Puerto Rico.

By AHI, frequent reports of ‘too hot’ SDITE were highest among participants with AHI ≥$100,000 (11%), followed by participants with $50,000-$99,999 (8.7%) and with ≤$49,999 (7.2%) (p<0.05; Supplemental Table 3). Frequent reports of ‘too cold’ SDITE were highest (p<0.05) among participants with AHIs ≤$49,999 (2.2%) but were similar between participants with AHIs of $50,000-$99,999 (1.1%) and ≥$100,000 (1.1%). All poor sleep dimensions were most prevalent among participants with AHIs ≤$49,999.

Both perceived SDITE and sleep health also varied by region of residence as well as menopausal status (all p<0.05; Supplemental Tables 4-5). Perceptions of ‘too hot’ SDITE were highest among participants residing in the South (9.3%) and West (9.0%) but perceptions of ‘too cold’ SDITE were highest among residents of Puerto Rico (8.4% vs. range 1.2%-1.5% in other regions). Premenopausal compared to postmenopausal participants were more likely to report both ‘too hot’ (14% vs. 8.8%) and ‘too cold’ (1.9% vs. 1.4%) SDITE. Conversely, sleep disturbances and HPD sleep apnea were more prevalent among postmenopausal versus premenopausal women.

### Frequently perceiving ‘too hot’ SDITE and multiple sleep dimensions

#### Overall

Compared to participants who reported infrequent SDITE, participants who reported frequent SDITE that were ‘too hot’ had a higher prevalence of all poor sleep characteristics even after adjustment for trouble sleeping for other reasons and FDR correction (Supplemental Table 6, Model 1). After full adjustment for sociodemographic and clinical characteristics along with FDR correction, frequently perceiving ‘too hot’ SDITE was associated with a higher prevalence of long sleep duration (PR=1.25 [95% CI:1.15,1.36]), LSOL (PR=1.88 [1.74-2.03]), poor sleep maintenance (PR=1.57 [1.52-1.62]), insomnia symptoms (PR=1.56 [1.52-1.61]), sleep medication use (PR=1.18 [1.09, 1.27]), daytime dysfunction (PR=1.76 [1.23, 2.52]), and HPD sleep apnea (PR = 1.20 [1.09, 1.33]). (Figure 1; Supplemental Table 6, Model 3).

**Figure 1.**
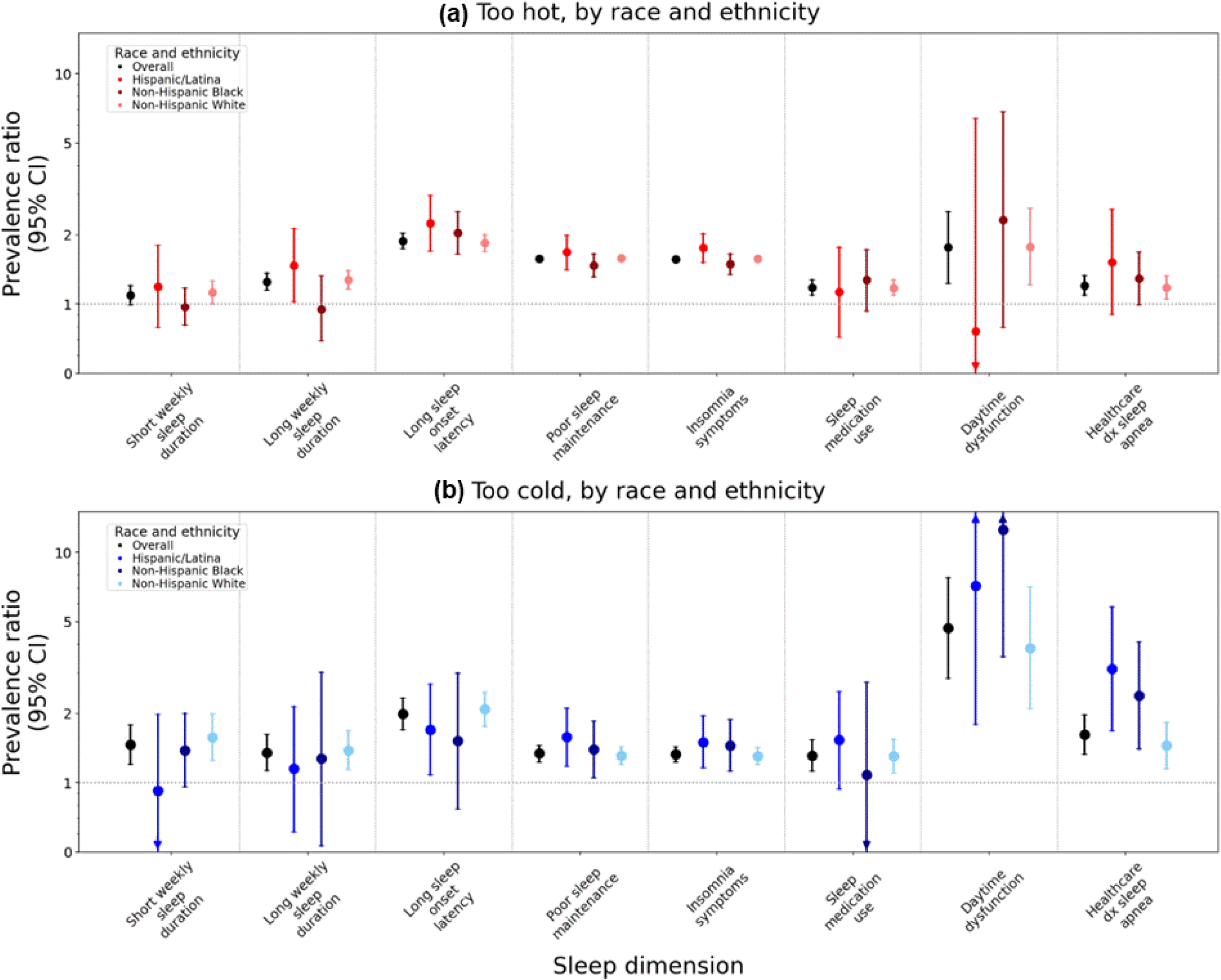
Prevalence ratios and 95% confidence intervals for associations between perceived sleep disrupting temperature extremes and sleep dimensions, overall and by race and ethnicity. Models are adjusted for trouble sleeping for any reason other than feeling too hot or feeling too cold, age (continuous), age^2^, race and ethnicity (in overall model: Hispanic/Latina, non-Hispanic Black, non-Hispanic White), annual household income (<$20,000 - $49,999, $50,000 - $99,999, ≥$100,000), marital status (married or living as though married, divorced/widowed/separated/never married), region of residence (Northeast, Midwest, South, West, and Puerto Rico), body mass index (BMI: underweight, recommended, overweight, obesity), and menopausal status (premenopausal, postmenopausal).

#### By Race and Ethnicity

Although magnitudes of association varied by race and ethnicity (Figure 1), we did not observe effect modification (all Wald p-values >0.05; Supplemental Table 7A). However, there was evidence of additive interaction among Latina compared to NH White participants for LSOL (RERI=0.79 [95% CI: 0.06,1.52]), suggesting synergism: the combination of frequently perceiving ‘too hot’ SDITE and self-identification as Latina was more strongly associated with higher prevalence of LSOL than expected if each was reported alone and summed (Supplemental Table 7B). Conversely, the combination of ‘too hot’ SDITE and self-identification as NH Black was less associated with both poor sleep maintenance and insomnia symptoms than expected if each was reported alone and summed.

#### By Annual Household Income

AHI did not modify relative associations between reports of frequent SDITE that were ‘too hot’ and the measured sleep health dimensions (all Wald p-values >0.05; Figure 2, Supplemental Table 8A). However, there was evidence of synergism between frequent SDITE that were ‘too hot’ and AHI on associations with LSOL as well as sleep medication use (Supplemental Table 8B). Specifically, the combination of low (RERI_<$20,000 -$49,999_=0.58 [0.19,0.97]) or middle AHI (RERI_$50,000 - $99,999_=0.51 [0.18,0.84]) with frequently perceiving ‘too hot’ SDITE had stronger than expected joint associations with higher prevalence of LSOL than if each was reported alone and summed. Additionally, the combination of low versus high AHI along with frequently perceiving ‘too hot’ SDITE also had a stronger than expected associations (RERI_<$20,000 - $49,999_=0.24 [0.01,0.47]) with sleep medication use than the sum of each alone.

**Figure 2.**
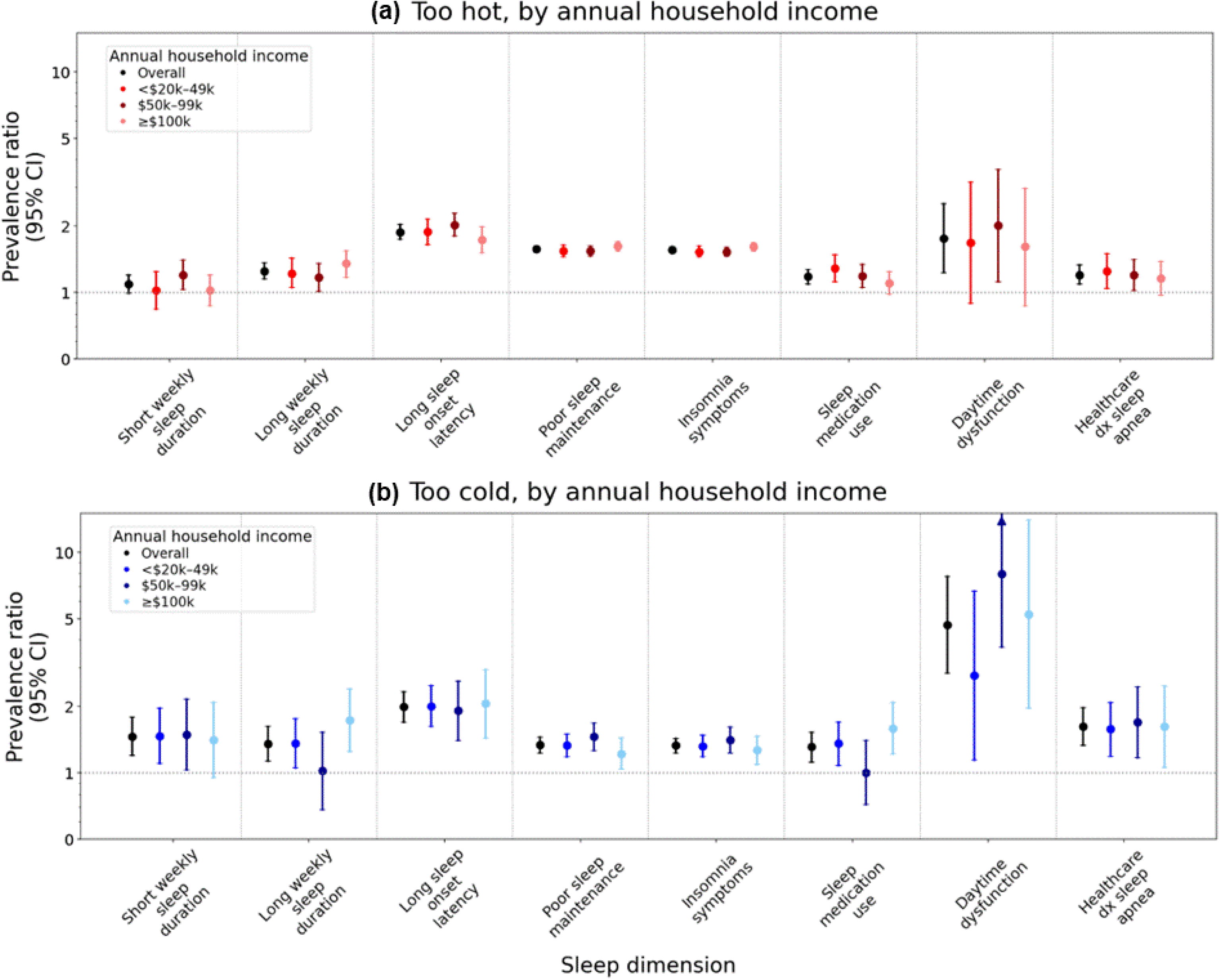
Prevalence ratios and 95% confidence intervals for associations between perceived sleep disrupting temperature extremes and sleep dimensions, overall and by annual household income. Models are adjusted for trouble sleeping for any reason other than feeling too hot or feeling too cold, age (continuous), age^2^, race and ethnicity (Hispanic/Latina, non-Hispanic Black, non-Hispanic White), annual household income (in overall model: <$20,000 - $49,999, $50,000 - $99,999, ≥$100,000), marital status (married or living as though married, divorced/widowed/separated/never married), region of residence (Northeast, Midwest, South, West, and Puerto Rico), body mass index (BMI: underweight, recommended, overweight, obesity), and menopausal status (premenopausal, postmenopausal).

#### By Other Potential Modifiers

Associations between frequently vs. infrequently perceiving ‘too hot’ SDITE and sleep duration varied by region of residence; however, there was no evidence of multiplicative effect modification or additive interaction by menopausal status for any sleep dimensions (Tables 2-3 and Supplemental Tables 9-10). Specifically, associations between frequently perceiving SDITE that were ‘too hot’ with both short and long sleep duration were strongest (both Wald p-values <0.01) among participants who resided in Puerto Rico.

**Table 2.**
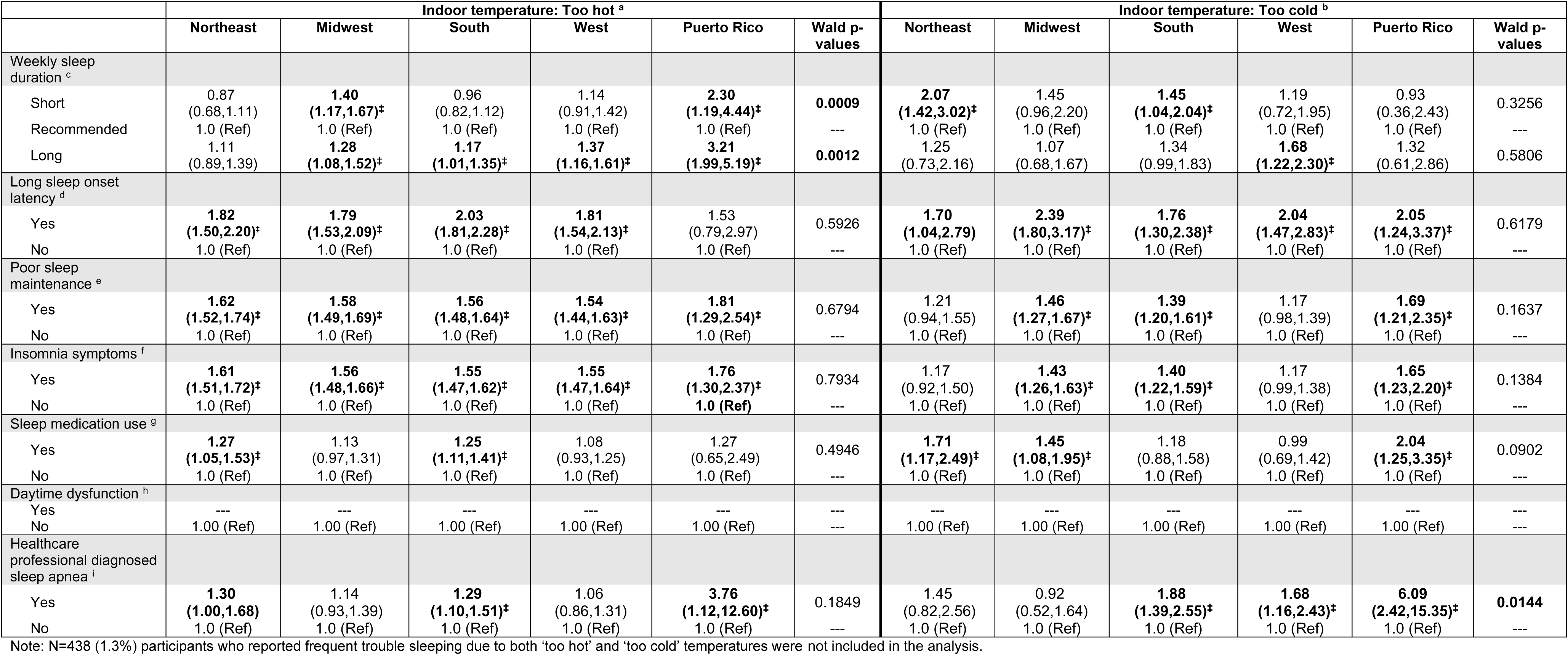
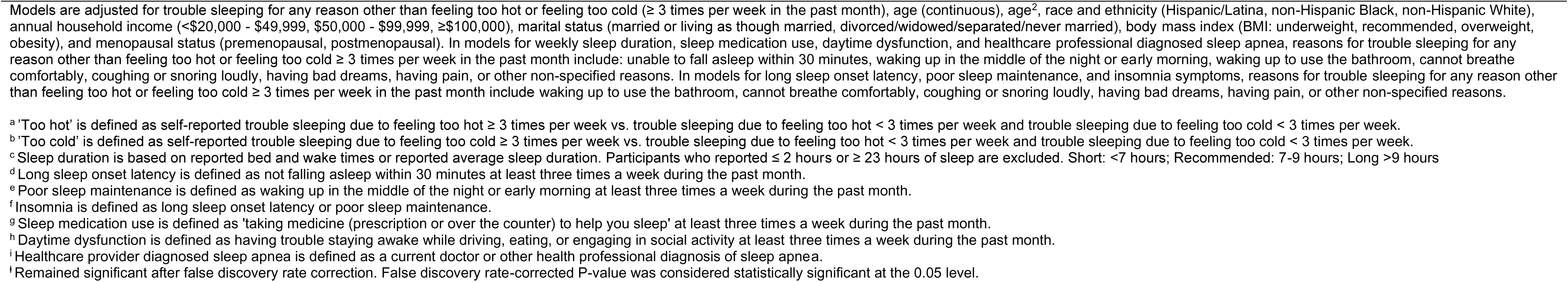
Prevalence ratios (95% confidence intervals) for associations between perceived indoor temperature extremes and sleep health dimensions by region of residence, Sister Study, 2017-2019, N=33,545.

|  | Indoor temperature: Too hot <sup>a</sup> |  |  |  |  |  | Indoor temperature: Too cold <sup>b</sup> |  |  |  |  |  |
| --- | --- | --- | --- | --- | --- | --- | --- | --- | --- | --- | --- | --- |
|  | Northeast | Midwest | South | West | Puerto Rico | Wald p-values | Northeast | Midwest | South | West | Puerto Rico | Wald p-values |
| Weekly sleep duration <sup>c</sup> |  |  |  |  |  |  |  |  |  |  |  |  |
| Short | 0.87<br>(0.68,1.11) | **1.40**<br>**(1.17,1.67)\*** | 0.96<br>(0.82,1.12) | 1.14<br>(0.91,1.42) | **2.30**<br>**(1.19,4.44)\*** | **0.0009** | **2.07**<br>**(1.42,3.02)\*** | 1.45<br>(0.96,2.20) | **1.45**<br>**(1.04,2.04)\*** | 1.19<br>(0.72,1.95) | 0.93<br>(0.36,2.43) | 0.3256 |
| Recommended | 1.0 (Ref) | 1.0 (Ref) | 1.0 (Ref) | 1.0 (Ref) | 1.0 (Ref) | --- | 1.0 (Ref) | 1.0 (Ref) | 1.0 (Ref) | 1.0 (Ref) | 1.0 (Ref) | --- |
| Long | 1.11<br>(0.89,1.39) | **1.28**<br>**(1.08,1.52)\*** | **1.17**<br>**(1.01,1.35)\*** | **1.37**<br>**(1.16,1.61)\*** | **3.21**<br>**(1.99,5.19)\*** | **0.0012** | 1.25<br>(0.73,2.16) | 1.07<br>(0.68,1.67) | 1.34<br>(0.99,1.83) | **1.68**<br>**(1.22,2.30)\*** | 1.32<br>(0.61,2.86) | 0.5806 |
| Long sleep onset latency <sup>d</sup> |  |  |  |  |  |  |  |  |  |  |  |  |
| Yes | **1.82**<br>**(1.50,2.20)\*** | **1.79**<br>**(1.53,2.09)\*** | **2.03**<br>**(1.81,2.28)\*** | **1.81**<br>**(1.54,2.13)\*** | 1.53<br>(0.79,2.97) | 0.5926 | **1.70**<br>**(1.04,2.79)** | **2.39**<br>**(1.80,3.17)\*** | **1.76**<br>**(1.30,2.38)\*** | **2.04**<br>**(1.47,2.83)\*** | **2.05**<br>**(1.24,3.37)\*** | 0.6179 |
| No | 1.0 (Ref) | 1.0 (Ref) | 1.0 (Ref) | 1.0 (Ref) | 1.0 (Ref) | --- | 1.0 (Ref) | 1.0 (Ref) | 1.0 (Ref) | 1.0 (Ref) | 1.0 (Ref) | --- |
| Poor sleep maintenance <sup>e</sup> |  |  |  |  |  |  |  |  |  |  |  |  |
| Yes | **1.62**<br>**(1.52,1.74)\*** | **1.58**<br>**(1.49,1.69)\*** | **1.56**<br>**(1.48,1.64)\*** | **1.54**<br>**(1.44,1.63)\*** | **1.81**<br>**(1.29,2.54)\*** | 0.6794 | 1.21<br>(0.94,1.55) | **1.46**<br>**(1.27,1.67)\*** | **1.39**<br>**(1.20,1.61)\*** | 1.17<br>(0.98,1.39) | **1.69**<br>**(1.21,2.35)\*** | 0.1637 |
| No | 1.0 (Ref) | 1.0 (Ref) | 1.0 (Ref) | 1.0 (Ref) | 1.0 (Ref) | --- | 1.0 (Ref) | 1.0 (Ref) | 1.0 (Ref) | 1.0 (Ref) | 1.0 (Ref) | --- |
| Insomnia symptoms <sup>f</sup> |  |  |  |  |  |  |  |  |  |  |  |  |
| Yes | **1.61**<br>**(1.51,1.72)\*** | **1.56**<br>**(1.48,1.66)\*** | **1.55**<br>**(1.47,1.62)\*** | **1.55**<br>**(1.47,1.64)\*** | **1.76**<br>**(1.30,2.37)\*** | 0.7934 | 1.17<br>(0.92,1.50) | **1.43**<br>**(1.26,1.63)\*** | **1.40**<br>**(1.22,1.59)\*** | 1.17<br>(0.99,1.38) | **1.65**<br>**(1.23,2.20)\*** | 0.1384 |
| No | 1.0 (Ref) | 1.0 (Ref) | 1.0 (Ref) | 1.0 (Ref) | **1.0 (Ref)** | --- | 1.0 (Ref) | 1.0 (Ref) | 1.0 (Ref) | 1.0 (Ref) | 1.0 (Ref) | --- |
| Sleep medication use <sup>g</sup> |  |  |  |  |  |  |  |  |  |  |  |  |
| Yes | **1.27**<br>**(1.05,1.53)\*** | 1.13<br>(0.97,1.31) | **1.25**<br>**(1.11,1.41)\*** | 1.08<br>(0.93,1.25) | 1.27<br>(0.65,2.49) | 0.4946 | **1.71**<br>**(1.17,2.49)\*** | **1.45**<br>**(1.08,1.95)\*** | 1.18<br>(0.88,1.58) | 0.99<br>(0.69,1.42) | **2.04**<br>**(1.25,3.35)\*** | 0.0902 |
| No | 1.0 (Ref) | 1.0 (Ref) | 1.0 (Ref) | 1.0 (Ref) | 1.0 (Ref) | --- | 1.0 (Ref) | 1.0 (Ref) | 1.0 (Ref) | 1.0 (Ref) | 1.0 (Ref) | --- |
| Daytime dysfunction <sup>h</sup> |  |  |  |  |  |  |  |  |  |  |  |  |
| Yes | --- | --- | --- | --- | --- | --- | --- | --- | --- | --- | --- | --- |
| No | 1.00 (Ref) | 1.00 (Ref) | 1.00 (Ref) | 1.00 (Ref) | 1.00 (Ref) | --- | 1.00 (Ref) | 1.00 (Ref) | 1.00 (Ref) | 1.00 (Ref) | 1.00 (Ref) | --- |
| Healthcare professional diagnosed sleep apnea <sup>i</sup> |  |  |  |  |  |  |  |  |  |  |  |  |
| Yes | **1.30**<br>**(1.00,1.68)** | 1.14<br>(0.93,1.39) | **1.29**<br>**(1.10,1.51)\*** | 1.06<br>(0.86,1.31) | **3.76**<br>**(1.12,12.60)\*** | 0.1849 | 1.45<br>(0.82,2.56) | 0.92<br>(0.52,1.64) | **1.88**<br>**(1.39,2.55)\*** | **1.68**<br>**(1.16,2.43)\*** | **6.09**<br>**(2.42,15.35)\*** | **0.0144** |
| No | 1.0 (Ref) | 1.0 (Ref) | 1.0 (Ref) | 1.0 (Ref) | 1.0 (Ref) | --- | 1.0 (Ref) | 1.0 (Ref) | 1.0 (Ref) | 1.0 (Ref) | 1.0 (Ref) | --- |

Note: N=438 (1.3%) participants who reported frequent trouble sleeping due to both ‘too hot’ and ‘too cold’ temperatures were not included in the analysis.
Models are adjusted for trouble sleeping for any reason other than feeling too hot or feeling too cold ( $\geq 3$ times per week in the past month), age (continuous), age<sup>2</sup>, race and ethnicity (Hispanic/Latina, non-Hispanic Black, non-Hispanic White), annual household income (<\$20,000 - \$49,999, \$50,000 - \$99,999, ≥ \$100,000), marital status (married or living as though married, divorced/widowed/separated/never married), body mass index (BMI: underweight, recommended, overweight, obesity), and menopausal status (premenopausal, postmenopausal). In models for weekly sleep duration, sleep medication use, daytime dysfunction, and healthcare professional diagnosed sleep apnea, reasons for trouble sleeping for any reason other than feeling too hot or feeling too cold $\geq 3$ times per week in the past month include: unable to fall asleep within 30 minutes, waking up in the middle of the night or early morning, waking up to use the bathroom, cannot breathe comfortably, coughing or snoring loudly, having bad dreams, having pain, or other non-specified reasons. In models for long sleep onset latency, poor sleep maintenance, and insomnia symptoms, reasons for trouble sleeping for any reason other than feeling too hot or feeling too cold $\geq 3$ times per week in the past month include waking up to use the bathroom, cannot breathe comfortably, coughing or snoring loudly, having bad dreams, having pain, or other non-specified reasons.
<sup>a</sup> 'Too hot' is defined as self-reported trouble sleeping due to feeling too hot $\geq 3$ times per week vs. trouble sleeping due to feeling too hot < 3 times per week and trouble sleeping due to feeling too cold < 3 times per week.
<sup>b</sup> 'Too cold' is defined as self-reported trouble sleeping due to feeling too cold $\geq 3$ times per week vs. trouble sleeping due to feeling too hot < 3 times per week and trouble sleeping due to feeling too cold < 3 times per week.
<sup>c</sup> Sleep duration is based on reported bed and wake times or reported average sleep duration. Participants who reported $\leq 2$ hours or $\geq 23$ hours of sleep are excluded. Short: <7 hours; Recommended: 7-9 hours; Long >9 hours
<sup>d</sup> Long sleep onset latency is defined as not falling asleep within 30 minutes at least three times a week during the past month.
<sup>e</sup> Poor sleep maintenance is defined as waking up in the middle of the night or early morning at least three times a week during the past month.
<sup>f</sup> Insomnia is defined as long sleep onset latency or poor sleep maintenance.
<sup>g</sup> Sleep medication use is defined as 'taking medicine (prescription or over the counter) to help you sleep' at least three times a week during the past month.
<sup>h</sup> Daytime dysfunction is defined as having trouble staying awake while driving, eating, or engaging in social activity at least three times a week during the past month.

**Table 3.**
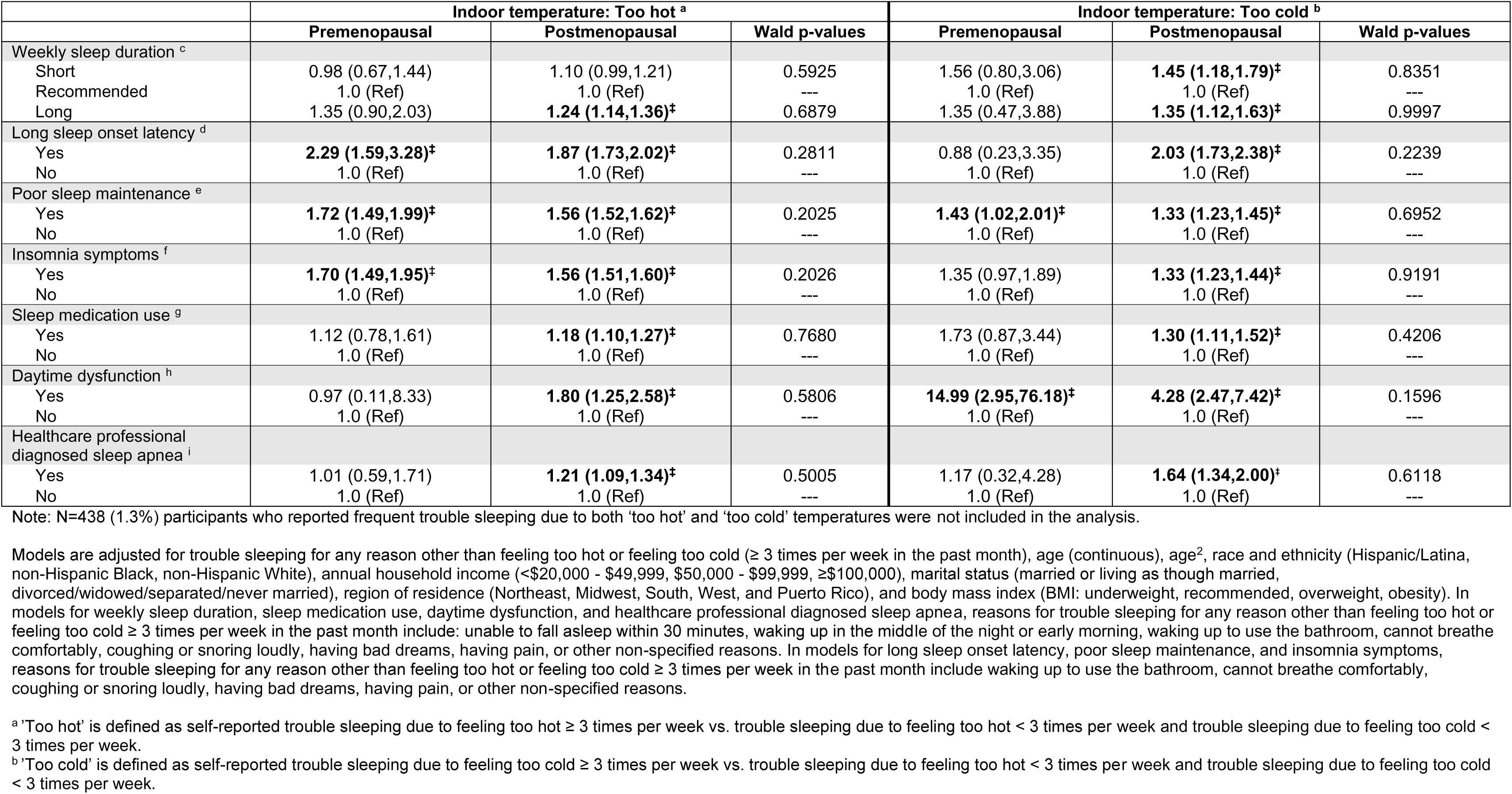
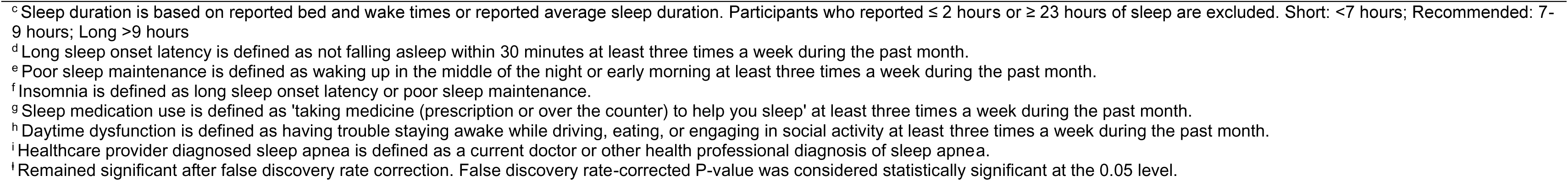
Prevalence ratios (95% confidence intervals) for associations between indoor temperature extremes and sleep health dimensions by menopausal status, Sister Study, 2017 – 2019, N = 33,454.

|  | Indoor temperature: Too hot <sup>a</sup> |  |  | Indoor temperature: Too cold <sup>b</sup> |  |  |
| --- | --- | --- | --- | --- | --- | --- |
|  | Premenopausal | Postmenopausal | Wald p-values | Premenopausal | Postmenopausal | Wald p-values |
| Weekly sleep duration <sup>c</sup> |  |  |  |  |  |  |
| Short | 0.98 (0.67,1.44) | 1.10 (0.99,1.21) | 0.5925 | 1.56 (0.80,3.06) | **1.45 (1.18,1.79)<sup>‡</sup>** | 0.8351 |
| Recommended | 1.0 (Ref) | 1.0 (Ref) | --- | 1.0 (Ref) | 1.0 (Ref) | --- |
| Long | 1.35 (0.90,2.03) | **1.24 (1.14,1.36)<sup>‡</sup>** | 0.6879 | 1.35 (0.47,3.88) | **1.35 (1.12,1.63)<sup>‡</sup>** | 0.9997 |
| Long sleep onset latency <sup>d</sup> |  |  |  |  |  |  |
| Yes | **2.29 (1.59,3.28)<sup>‡</sup>** | **1.87 (1.73,2.02)<sup>‡</sup>** | 0.2811 | 0.88 (0.23,3.35) | **2.03 (1.73,2.38)<sup>‡</sup>** | 0.2239 |
| No | 1.0 (Ref) | 1.0 (Ref) | --- | 1.0 (Ref) | 1.0 (Ref) | --- |
| Poor sleep maintenance <sup>e</sup> |  |  |  |  |  |  |
| Yes | **1.72 (1.49,1.99)<sup>‡</sup>** | **1.56 (1.52,1.62)<sup>‡</sup>** | 0.2025 | **1.43 (1.02,2.01)<sup>‡</sup>** | **1.33 (1.23,1.45)<sup>‡</sup>** | 0.6952 |
| No | 1.0 (Ref) | 1.0 (Ref) | --- | 1.0 (Ref) | 1.0 (Ref) | --- |
| Insomnia symptoms <sup>f</sup> |  |  |  |  |  |  |
| Yes | **1.70 (1.49,1.95)<sup>‡</sup>** | **1.56 (1.51,1.60)<sup>‡</sup>** | 0.2026 | 1.35 (0.97,1.89) | **1.33 (1.23,1.44)<sup>‡</sup>** | 0.9191 |
| No | 1.0 (Ref) | 1.0 (Ref) | --- | 1.0 (Ref) | 1.0 (Ref) | --- |
| Sleep medication use <sup>g</sup> |  |  |  |  |  |  |
| Yes | 1.12 (0.78,1.61) | **1.18 (1.10,1.27)<sup>‡</sup>** | 0.7680 | 1.73 (0.87,3.44) | **1.30 (1.11,1.52)<sup>‡</sup>** | 0.4206 |
| No | 1.0 (Ref) | 1.0 (Ref) | --- | 1.0 (Ref) | 1.0 (Ref) | --- |
| Daytime dysfunction <sup>h</sup> |  |  |  |  |  |  |
| Yes | 0.97 (0.11,8.33) | **1.80 (1.25,2.58)<sup>‡</sup>** | 0.5806 | **14.99 (2.95,76.18)<sup>‡</sup>** | **4.28 (2.47,7.42)<sup>‡</sup>** | 0.1596 |
| No | 1.0 (Ref) | 1.0 (Ref) | --- | 1.0 (Ref) | 1.0 (Ref) | --- |
| Healthcare professional diagnosed sleep apnea <sup>i</sup> |  |  |  |  |  |  |
| Yes | 1.01 (0.59,1.71) | **1.21 (1.09,1.34)<sup>‡</sup>** | 0.5005 | 1.17 (0.32,4.28) | **1.64 (1.34,2.00)<sup>‡</sup>** | 0.6118 |
| No | 1.0 (Ref) | 1.0 (Ref) | --- | 1.0 (Ref) | 1.0 (Ref) | --- |

Note: N=438 (1.3%) participants who reported frequent trouble sleeping due to both ‘too hot’ and ‘too cold’ temperatures were not included in the analysis.
Models are adjusted for trouble sleeping for any reason other than feeling too hot or feeling too cold (≥ 3 times per week in the past month), age (continuous), age<sup>2</sup>, race and ethnicity (Hispanic/Latina, non-Hispanic Black, non-Hispanic White), annual household income (<\$20,000 - \$49,999, \$50,000 - \$99,999, ≥\$100,000), marital status (married or living as though married, divorced/widowed/separated/never married), region of residence (Northeast, Midwest, South, West, and Puerto Rico), and body mass index (BMI: underweight, recommended, overweight, obesity). In models for weekly sleep duration, sleep medication use, daytime dysfunction, and healthcare professional diagnosed sleep apnea, reasons for trouble sleeping for any reason other than feeling too hot or feeling too cold ≥ 3 times per week in the past month include: unable to fall asleep within 30 minutes, waking up in the middle of the night or early morning, waking up to use the bathroom, cannot breathe comfortably, coughing or snoring loudly, having bad dreams, having pain, or other non-specified reasons. In models for long sleep onset latency, poor sleep maintenance, and insomnia symptoms, reasons for trouble sleeping for any reason other than feeling too hot or feeling too cold ≥ 3 times per week in the past month include waking up to use the bathroom, cannot breathe comfortably, coughing or snoring loudly, having bad dreams, having pain, or other non-specified reasons.
<sup>a</sup> ‘Too hot’ is defined as self-reported trouble sleeping due to feeling too hot ≥ 3 times per week vs. trouble sleeping due to feeling too hot < 3 times per week and trouble sleeping due to feeling too cold < 3 times per week.
<sup>b</sup> ‘Too cold’ is defined as self-reported trouble sleeping due to feeling too cold ≥ 3 times per week vs. trouble sleeping due to feeling too hot < 3 times per week and trouble sleeping due to feeling too cold < 3 times per week.
<sup>c</sup> Sleep duration is based on reported bed and wake times or reported average sleep duration. Participants who reported $\leq 2$ hours or $\geq 23$ hours of sleep are excluded. Short: $<7$ hours; Recommended: 7-9 hours; Long $>9$ hours
<sup>d</sup> Long sleep onset latency is defined as not falling asleep within 30 minutes at least three times a week during the past month.
<sup>e</sup> Poor sleep maintenance is defined as waking up in the middle of the night or early morning at least three times a week during the past month.
<sup>f</sup> Insomnia is defined as long sleep onset latency or poor sleep maintenance.
<sup>g</sup> Sleep medication use is defined as 'taking medicine (prescription or over the counter) to help you sleep' at least three times a week during the past month.
<sup>h</sup> Daytime dysfunction is defined as having trouble staying awake while driving, eating, or engaging in social activity at least three times a week during the past month.

Among women in Puerto Rico, women who frequently versus infrequently perceived ‘too hot’ SDITE had over two times the prevalence of short sleep (PR_Puerto Rico_=2.30 [1.19-4.44] vs. PR range= 0.87 [0.68-1.11] (Northeast) to 1.40 [1.17-1.67] (Midwest)) and over three times the prevalence of long sleep (PR_Puerto Rico_=3.21 [1.99-5.19] vs. PR range=1.11 [0.89-1.39] (Northeast) to 1.37 [1.16-1.61] (West)). For long sleep, there was also synergism between ‘too hot’ SDITE and region of residence in Puerto Rico vs. the South (RERI=1.45 [0.48, 2.42]). In the Midwest compared to the South, there was also synergism in associations with short sleep (RERI_Midwest_=0.41 [0.15,0.67]) and antagonism in associations with LSOL (RERI_Midwest_=-0.32 [- 0.63,-0.01).

### Frequently perceiving ‘too cold’ SDITE and multiple sleep dimensions

#### Overall

Compared to participants who reported infrequent SDITE, participants who reported frequent SDITE that were ‘too cold’ had a higher prevalence of all poor sleep dimensions after adjustment for trouble sleeping due to other reasons and FDR correction (Supplemental Table 6, Model 1). After full adjustment and FDR correction, frequently vs. infrequently perceiving ‘too cold’ SDITE remained associated with a higher prevalence of all poor sleep characteristics (Figure 1). The largest magnitudes of associations were for LSOL (PR=1.99 [1.70-2.33]), daytime dysfunction (PR=4.69 [2.83-7.77]), and HPD sleep apnea (PR=1.62 [1.33-1.97]).

#### By Race and Ethnicity

Although PRs varied by racial and ethnic group, CIs were wide, and there was little evidence of effect modification by race and ethnicity (most Wald p- value>0.05; Figure 1, Supplemental Table 7A). However, magnitudes of associations between frequently vs. infrequently perceiving ‘too cold’ SDITE and HPD sleep apnea were largest among Latina participants (PR=3.12 [1.68-5.79] vs. PR_NH Black_=2.39 [1.40-4.08] and PR_NH White_=1.45 [1.15-1.82, Wald p-value=0.03). Yet, there was no evidence of additive interaction across associations with any sleep dimensions (Supplemental Table 7B).

#### By Annual Household Income

Despite some dissimilarities in PRs by AHI, we did not observe effect modification by AHI for multiplicative associations with any sleep dimensions (Figure 2, Supplemental Table 8A). However, the RERI for associations between perceiving frequent vs. infrequent SDITE that were ‘too cold’ and sleep medication use was negative (-0.59 [-1.16, -0.02]) for participants with AHI of $50,000 - $99,000 vs. ≥$100,000 (Supplemental Table 8B). This suggests that the association between frequently perceiving ‘too cold’ SDITE coupled with middle income and sleep medication use was weaker than the sum of associations if each exposure was reported alone and summed.

#### By Other Potential Modifiers

There was effect modification by US region for multiplicative associations as well as additive interaction between frequently vs. infrequently perceiving ‘too cold’ SDITE and HPD sleep apnea (Table 2 and Supplemental Table 9). The magnitude of association was highest - albeit imprecise - among participants residing in Puerto Rico (PR=6.09 [2.42-15.35]) compared to null associations among participants residing in the Northeast and Midwest as well as compared to the associations observed among participants residing in the South (PR=1.88 [1.39-2.55]) and West (PR=1.68 [1.16-2.43], Wald p- value=0.01). Additionally, for HPD sleep apnea, there was antagonism (RERI_Midwest vs South_ =- 0.96 [-1.73, -0.19]). While there was no observed effect modification by menopausal status for any relative associations (all Wald p-values>0.05; Table 3), there was evidence of synergism among postmenopausal compared to premenopausal women in associations with LSOL (RERI=1.69 [0.43,2.95]; Supplemental Table 10).

#### Sensitivity analysis

In analyses restricted to participants who reported trouble sleeping for other reasons than temperature extremes, results were generally consistent with the main analysis although attenuated (Supplemental Tables 11-20B); exceptions were newly observed negative additive interaction with middle income (AHI $50,000 - $99,0000) in ‘too cold’ – long sleep duration associations and positive additive interaction with postmenopausal status in ‘too cold’ – HPD sleep apnea associations. Frequently perceiving SDITE remained more often reported by NH Black and Latina compared to NH White women. Like the main analysis, relative associations did not vary by race and ethnicity, yet differing from the main analysis, RERIs by race and ethnicity for associations between frequently perceiving ‘too hot’ SDITE with LSOL, poor sleep maintenance, and insomnia symptoms were attenuated and no longer significant. Consistent with the main analysis, PRs were largest among Latina women for associations between SDITE that were ‘too cold’ and HPD sleep apnea, although CIs overlapped with estimates among the other racial and ethnic groups and Wald p-value=0.20.

Women with AHI ≥$100,000 remained most likely to report SDITE that were ‘too hot’ while women with AHI ≤$49,999 remained most likely to report SDITE that were ‘too cold’. Also consistent with the main analysis there was synergism between frequently perceiving ‘too hot’ SDITE and AHI on LSOL in the middle AHI ($50,000 - $99,000) vs. high AHI (≥$100,000) categories. Similarly consistent with the main analysis, there was no observation of effect modification on the multiplicative scale, and the antagonism between middle vs. high AHI and ‘too cold’ SDITE on the sleep medication use association, although attenuated, remained.

Results for the exploratory analyses considering region of residence and menopausal status as potential effect modifiers were also consistent with the main analysis. For instance, relative associations between frequently vs. infrequently perceiving SDITE that were ‘too hot’ with both short and long sleep duration as well as frequently vs. infrequently perceiving SDITE that were ‘too cold’ and HPD sleep apnea remained strongest among women residing in Puerto Rico.

## DISCUSSION

Among a cohort of women of older age living in the US, we estimated the prevalence of perceived indoor temperature extremes that disrupt sleep. We also characterized sleep disturbances among women who frequently vs. infrequently reported sleep-disrupting indoor temperature extremes (SDITE) and determined whether observed relationships differed across race and ethnicity as well as annual household income (AHI) categories. Overall, we found that reports of trouble sleeping due to ‘too hot’ temperatures were prevalent (9.0%) while reported trouble sleeping due to ‘too cold’ temperatures was less prevalent (1.4%). Consistent with our hypotheses, NH Black women were most likely to frequently report SDITE that were ‘too hot’; prevalence was six percentage points higher compared to NH White women. Inconsistent with our hypothesis, perceiving ‘too hot’ SDITE was most reported by women within the highest income category. Nonetheless, perceiving ‘too hot’ SDITE was associated with all poor sleep dimensions measured in this study except short sleep duration even after adjustment for trouble sleeping due to other factors (e.g., pain, inability to breathe comfortably), other relevant confounders, and multiple comparisons; associations were strongest with LSOL and daytime dysfunction – defined as trouble staying awake while driving, eating meals, or engaging in social activity. Moreover, the apparent additive influences of ‘too hot’ SDITE and self-identifying as Latina as well as of ‘too hot’ SDITE and low income (≤$49,999) on associations with LSOL were larger than the sum of each. Also consistent with our hypothesis, Latina women and women within the lowest AHI category most often reported SDITE that were ‘too cold’; Latina women had three times the prevalence observed among NH White women, and women with AHI ≤$49,999 versus ≥$100,000 had two times the prevalence. These SDITE perceived as ‘too cold’ were associated with poorer sleep dimensions, especially with a higher prevalence of LSOL, daytime dysfunction, and HPD sleep apnea – an outcome in which associations were strongest among Latina women. Notably, groups with either the highest burdens or strongest relationships (e.g., Latina, low-income) are populations more likely to experience energy insecurity or difficulty meeting household energy needs.^43^

We also explored whether relationships varied by two potentially important effect modifiers, geographic region of residence and menopausal status. Aligned with observations of stronger additive associations with LSOL among Latina women, frequently vs. infrequently perceiving SDITE was more strongly associated with short sleep duration (‘too hot’), long sleep duration (‘too hot’), and HPD sleep apnea (‘too cold’) among women in Puerto Rico, the majority of which are Latina. Menopausal status did not act as an effect modifier; however, additive associations between perceiving SDITE that were ‘too cold’ with LSOL appear stronger among postmenopausal compared to menopausal women.

Explicitly, after adjustment and multiple comparison correction, women who reported frequent versus infrequent ‘too hot’ SDITE also reported longer sleep duration (>9 hours), LSOL, poor sleep maintenance, sleep medication use, daytime dysfunction, and HPD sleep apnea. Furthermore, after adjustment and multiple comparison correction, women who reported frequently versus infrequently perceiving ‘too cold’ SDITE had a higher prevalence of all poor sleep dimensions. For each temperature extreme, the highest magnitudes of association were with LSOL and daytime dysfunction. Overall, the findings suggest that women with perceived temperature extremes – whether too hot or too cold – are especially more likely to report delayed sleep onset and sleep that is generally more disturbed and non-restorative, given observed associations with daytime dysfunction.

The general relationships observed between perceived SDITE and sleep are mostly consistent with prior literature. Except for a few prior studies,^29,36^ studies generally suggest higher indoor temperatures as associated with LSOL, poor sleep quality, and sleep disturbances;^26–28,32,37,41,60–63^ thus prior literature related to hot indoor temperatures and poorer sleep health mostly agree with our findings. In fact, our findings related to ‘too hot’ temperatures align with a recent systematic review that showed higher indoor temperatures as persistently associated with degraded sleep quality, whether measured subjectively or objectively.^22^ However, our results suggesting no associations between perceived ‘too hot’ temperatures and short sleep duration add to the mixed evidence to date, with some studies suggesting an association and other suggesting no association.^22,29,32,61,64–70^ Lastly, our findings related to perceiving ‘too cold’ SDITE are consistent with prior reviews and original research in which the majority reported cold indoor temperatures as associated with LSOL, insomnia symptoms, poor sleep quality, and lower time in slow wave sleep, which correspond with our observed associations.^23,28,33–35,38,41^

Regarding differences by race and ethnicity as well as SES, our hypotheses were supported for certain groups and sleep dimensions. The most consistent differences in additive relationships were between perceived ‘too hot’ SDITE and LSOL in which both Latina ethnicity and ‘lower’ compared to ‘high’ AHI acted synergistically, suggesting potentially larger public health impacts on these groups. Moreover, associations between frequently versus infrequently perceiving ‘too hot’ SDITE and both short and long sleep duration were much stronger in Puerto Rico, a region which largely comprised Latina women, more so than in other regions.

Additionally, associations between frequently perceiving ‘too cold’ temperature extremes and a higher prevalence of HPD sleep apnea were stronger among Latina women compared to NH White women as well as in Puerto Rico. In combination, results suggest that sleep health among Latina women, particularly Latina women residing in Puerto Rico, may be particularly vulnerable to temperature extremes. Regarding synergism between ‘too hot’ SDITE and AHI, results were plausible even though women with ‘high’ AHI had higher unadjusted prevalence of perceiving ‘too hot’ SDITE. Factors like poorer sleep health even without the exposure among women with ‘low’ AHI and other SES-related differences may have amplified worse sleep outcomes. Lastly, for perceived sleep-disrupting ‘too cold’ temperatures in relation to sleep medication use, there was antagonistic interaction among women with middle versus high AHI; it is possible that higher income individuals may more frequently use sleep medication to assuage sleep disturbances related to sleep environments perceived as ‘too cold’.

Our results may be explained by synthesizing and connecting results from prior studies, including studies of objective sleep that demonstrated how ambient temperatures impact sleep architecture. Prior literature suggests intensified temperature extremes indoors compared to outdoors^25^ and demonstrates strong relationships between perceived and actual temperatures.^32,41^ Perceptions of temperature extremes that disrupt sleep may accurately reflect reality. Additionally, studies demonstrate that women are particularly susceptible to temperature extremes compared to men,^44^ and that socially-relegated populations may encounter more temperature extremes than other populations due to factors such as energy insecurity,^42^ a social determinant of health related to low socioeconomic status that reflects residents’ limited ability to control indoor environments. The thermal environment is recognized as one of the primary causes of sleep disturbances, hypothesized as acting through biologic pathways, such as impacting receptors, neurotransmitters, neurons, and hormones that are involved in sleep and circadian rhythm activity.^40,71^ Cold temperatures have been found to increase the number and duration of wakefulness periods and lighter sleep stages, and hot or high temperatures can reduce duration of deep sleep and increase sleep onset latency as well as wakefulness.^13,71^ Therefore, the observed linkage between perceived temperature extremes and poor sleep – namely LSOL and daytime dysfunction may be explained by this accumulation of findings about temperatures and sleep architecture from prior literature.

While our findings related to effect modification and additive interaction extend the prior literature given limited prior assessments, they are generally congruent with prior literature. To our knowledge, only one prior study investigated both hot and cold indoor temperature extremes (versus one category alone) in a multiethnic US population while considering socioeconomic status – albeit using objective temperature measurements in a pediatric sample.^30^ Consistent with our current study, there were U-shaped relationships between indoor temperature and actigraphy-assessed poor sleep dimensions. Partially in contrast to our study, indoor temperature extremes were more prevalent among children in families with lower SES while we observed only colder perceived temperatures to be more prevalent among women with low AHI. In another study using an unsupervised machine learning approach, women and low SES respondents were mostly likely to report uncomfortable temperatures, and uncomfortable temperatures were associated with self-reported sleep problems.^31^ Additionally, similar to our observations of effect modification, reviews of the literature on outdoor temperatures and sleep suggest stronger impacts on women, marginalized and socially-relegated populations, and individuals residing in the warmest regions (i.e., Puerto Rico in our study).^16,17,22^ Additional studies of social characteristics as potential effect modifiers of the indoor temperature and sleep relationship are warranted.

There are limitations and strengths worthy of consideration. The cross-sectional design limits causal interpretations. Additionally, exposure misclassification is possible. We investigated perceived temperature extremes considered to cause the participants trouble sleeping.

Therefore, sleep disturbances are expected to be higher in the exposed compared to the unexposed group. However, all models were adjusted for trouble sleeping due to other reasons, which was endorsed by most participants. Thus, the modeling approach allowed for the potential isolation of sleep disturbances due to temperature. Additionally, our use of perception better approximates thermal comfort, which is an important yet understudied factor in temperature-sleep associations.^13,37,72,73^ Nonetheless, additional studies that include both objective and subjective indoor temperatures are necessary.^22^ Further studies inclusive of indoor environmental factors like humidity, air flow, and bed microenvironment (e.g., bedding, air conditioning), which impact thermal comfort,^37,40^ are also necessary. Our study included subjective sleep assessments, which are moderately correlated with objective sleep measurements;^74^ therefore, additional studies using objectively-measured sleep are also warranted. There were also small samples sizes who reported some exposures (e.g., participants who reported ‘too cold’) and outcomes (e.g., daytime dysfunction), which resulted in imprecise estimates. Unmeasured confounding related to factors like seasonality is also possible. Additionally, ineligible Sister Study participants were more likely to have characteristics associated with both perceived SDITE and poor sleep; therefore, current results may be underestimations. Moreover, the Sister Study sample consisted of older women with higher SES than the general US population, thus limiting generalizability. However, we expect the mechanisms linking temperature to sleep to be applicable across other populations.

Nevertheless, additional studies with various racial and ethnic, sex, age, and SES groups are warranted. Despite the limitations, strengths include the sample heterogeneity in racial, ethnic, and geography of US women along with the large sample size, which allowed for robust investigation of important potential modifiers. Additionally, we captured multiple sleep dimensions with certain dimensions assessed using a validated instrument (i.e., PSQI).

Furthermore, by investigating perceived temperature extremes and sleep disturbances in the context of factors like SES and menopausal status, our findings contribute to the literature by considering physical and psychosocial characteristics of people sleeping in bedrooms, which has been identified as needed to guide bedroom design interventions to improve sleep.^75^

In conclusion, we characterized sleep dimensions among an older-aged population of Latina, NH Black, and NH White US women who reported perceived indoor temperatures.

Women who reported frequent trouble sleeping due to being ‘too hot’ were more likely to identify as NH Black while women who reported frequent trouble sleeping due to being ‘too cold’ were more likely to identify as Latina; these SDITE were consistently strongly associated with LSOL and daytime dysfunction. Additionally, certain associations were generally stronger, and burdens were higher among Latina women, including Latina women in Puerto Rico. Moreover, we assessed perceived indoor temperature using the established PSQI, which can be used in many existing studies to capture thermal comfort coupled with the collection of objectively measured temperature and other features of the indoor/bedroom microenvironment to build upon our approach. Although more studies are warranted, optimizing indoor temperatures can plausibly improve sleep health and serve as a target for interventions to address racial and ethnic sleep disparities among older-aged US women.

## Data Availability

All data produced in the present study are available upon reasonable request to the authors

## ACKNOWLEDGEMENTS

This research was supported, in part, by the Intramural Research Program of the National Institutes of Health (NIH) (Z1AES103325 (CLJ) and Z01ES044005 (DPS)). The contributions of the NIH authors were made as part of their official duties as NIH federal employees, are in compliance with agency policy requirements, and are considered Works of the United States Government. However, the findings and conclusions presented in this paper are those of the authors and do not necessarily reflect the views of the NIH or the U.S. Department of Health and Human Services.

The authors wish to thank the Sister Study participants. The authors also wish to thank the NIEHS library staff, Stacey Mantooth, MSLS and Erin Knight, MLS, for performing the literature search for this manuscript as well as Elizabeth Teka for assistance with the review of the literature.

The results of this study were presented, in part, at World Sleep (Rio de Janeiro, Brazil) on October 24, 2023.

## Author Contributions

Conceptualization: CL Jackson

Formal Analysis: DT Neo

Funding Acquisition: CL Jackson, DP Sandler

Methodology: SA Gaston, DT Neo, CL Jackson

Project administration: WB Jackson, DP Sandler, CL Jackson

Resources: CL Jackson, DP Sandler

Supervision: CL Jackson, WB Jackson II

Writing- original draft: SA Gaston

Writing- reviewing and editing: SA Gaston, DT Neo, WB Jackson II, DP Sandler, CL Jackson

## ABBREVIATIONS

AHI: annual household income
BMI: body mass index
FDR: false discovery rate
HPD: healthcare professional diagnosed
LSOL: long sleep onset latency
NH: non-Hispanic
PSQI: Pittsburgh Sleep Quality Index
RERI: relative excess risk due to interaction
SES: socioeconomic status
SDITE: sleep-disrupting indoor temperature extremes

