## Supplemental Material for "Perceived indoor temperature extremes are associated with sleep health among women in the United States"

Symielle A. Gaston, PhD, MPH [1]

Dayna T. Neo, PhD, MPH [2]

W. Braxton Jackson II, MPH [2]

Dale P. Sandler, PhD [1]

Chandra L. Jackson, PhD, MS [1,3]

1 Epidemiology Branch, National Institute of Environmental Health Sciences, National Institutes of Health, Research Triangle Park, NC, USA

2 DLH LLC, Bethesda, MD, USA

3 Division of Intramural Research, National Institute on Minority Health and Health Disparities, National Institutes of Health, Bethesda, MD, USA

Please direct correspondence to: Dr. Chandra L. Jackson, PhD, MS, 111 TW Alexander Drive, Research Triangle Park, North Carolina, 27709, Phone: 984-287-3701, Fax: 301-480-3290,

**Supplemental Figure 1. Study population flow chart**

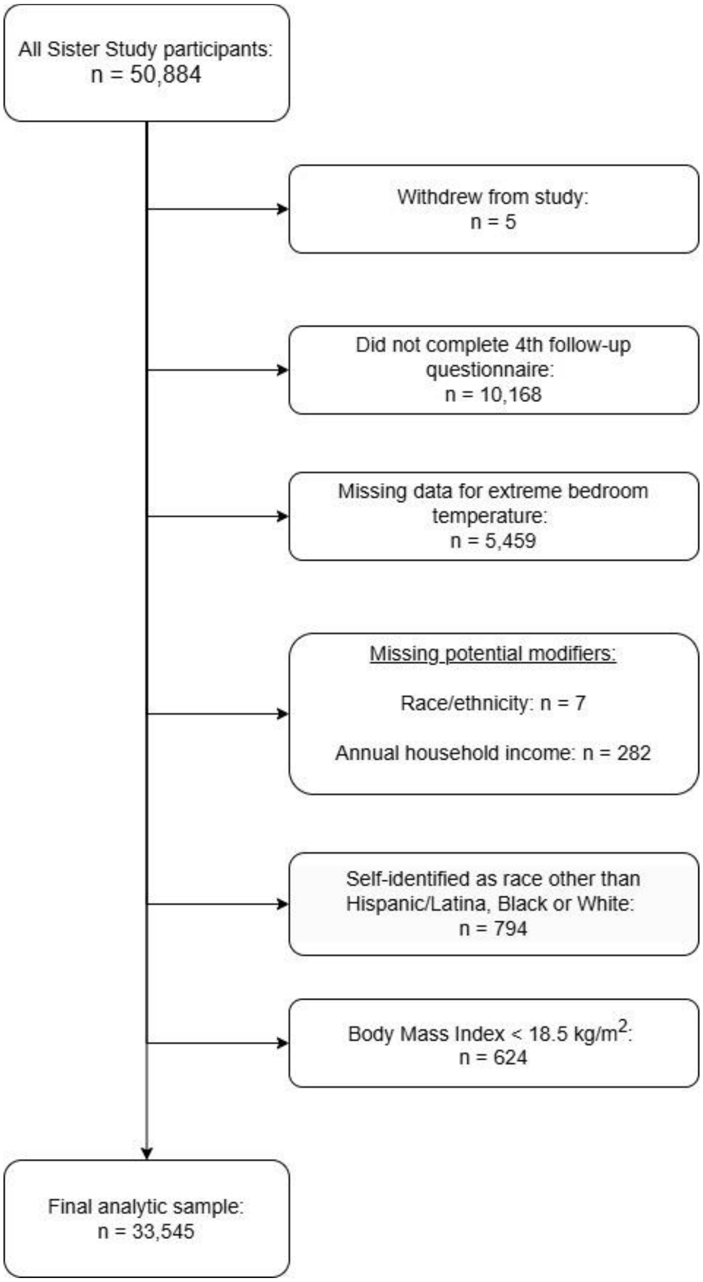

**Supplemental Table 1. Comparison of sociodemographic and clinical characteristics at enrollment (2003-2009) between eligible (n = 33,545) and ineligible (n=17,339) Sister Study participants (N=50,884)**

| | **Eligible n = 33,545 (65.9%)** | **Ineligible n = 17,339 (34.1%)** | **P-values** |
|---|---|---|---|
| **Sociodemographic characteristics** | | | |
| Age, years (mean ± SD) | 55.5 ± 8.6 | 56.0 ± 9.6 | <0.001 |
| Race and ethnicity | | | |
| Hispanic/Latina | 1,225 (3.7) | 1,290 (7.4) | <0.0001 |
| Non-Hispanic Black | 2,200 (6.6) | 2,262 (13.1) | |
| Non-Hispanic White | 30,120 (89.8) | 12,438 (71.7) | |
| Other | 0 (0.0) | 1,334 (7.7) | |
| *Missing* | *0 (0.0)* | *15 (0.1)* | |
| Educational attainment | | | |
| ≤High school or GED | 4,462 (13.3) | 3,342 (19.3) | <0.0001 |
| Some college/technical | 10,590 (31.6) | 6,590 (38.0) | |
| ≥ Bachelor's degree or higher | 18,489 (55.1) | 7,394 (42.6) | |
| *Missing* | *4 (0.01)* | *13 (0.1)* | |
| Annual household income | | | |
| <$20,000 - $49,999 | 7,095 (21.2) | 5,485 (31.6) | <0.0001 |
| $50,000-$99,999 | 13,601 (40.5) | 6,303 (36.4) | |
| ≥$100,000 | 11,913 (35.5) | 4,487 (25.9) | |
| *Missing* | 936 (2.8) | 1,064 (6.1) | |
| Marital status | | | |
| Married or living as though married | 25,855 (77.1) | 12,137 (70.0) | <0.0001 |
| Never married, divorced, widowed, or separated | 7,686 (22.9) | 5,187 (29.9) | |
| *Missing* | 4 (0.0) | 15 (0.1) | |
| Region of residence | | | |
| Northeast | 5,894 (17.6) | 2,621 (15.1) | <0.0001 |
| Midwest | 9,309 (27.8) | 4,306 (24.8) | |
| South | 10,559 (31.5) | 6,284 (36.2) | |
| West | 7,398 (22.1) | 3,635 (21.0) | |
| Puerto Rico | 385 (1.2) | 488 (2.8) | |
| *Missing* | *0 (0.0)* | *5 (0.0)* | |

| Clinical characteristics | | | |
| --- | --- | --- | --- |
| Body mass index, $kg/m^2$ | | | |
| Recommended (18.5 - 24.9) | 13,718 (40.9) | 5,910 (34.1) | <0.0001 |
| Overweight (25.0 - 29.9) | 10,582 (51.6) | 5,311 (30.6) | |
| Obesity (≥30) | 8,602 (25.6) | 5,469 (31.5) | |
| *Missing* | *643 (1.9)* | *649 (3.7)* | |
| Menopausal status | | | |
| Premenopausal | 11,328 (33.8) | 5,700 (32.9) | 0.04 |
| Postmenopausal | 22,210 (66.2) | 11,633 (67.1) | |
| *Missing or never had period* | *7 (0.0)* | *6 (0.0)* | |

Data presented as mean ± standard deviation or n (%)
Chi-square tests were used to compare categorical data.
A t-test was used to compare continuous age.

**Supplemental Table 2. Sociodemographic characteristics, clinical characteristics, and sleep health dimensions among eligible Sister Study participants by race and ethnicity as well as perceived indoor temperature extremes, Sister Study, 2017-2019, N=33,545**

| **Race and ethnicity** | **Hispanic/Latina [a]** | | | | **Non-Hispanic Black [b]** | | | | **Non-Hispanic White [c]** | | | |
|---|---|---|---|---|---|---|---|---|---|---|---|---|
| | Total n = 1,225 (3.7%) | Infrequent temperature extremes [d] n = 1,042 (85.1%) | Too hot [e ¥] n = 112 (9.1%) | Too cold [f §] n = 46 (3.8%) | Total n = 2,200 (6.6%) | Infrequent temperature extremes [d] n = 1,802 (81.9%) | Too hot [e ¥] n = 324 (14.7%) | Too cold [f §] n = 34 (1.6%) | Total n = 30,120 (89.8%) | Infrequent temperature extremes [d] n = 26,783 (88.9%) | Too hot [e ¥] n = 2,582 (8.6%) | Too cold [f §] n = 382 (1.3%) |
| **Sociodemographic characteristics** | | | | | | | | | | | | |
| Age, years (mean ± SD) [ᵻ] | 63.9 ± 8.5 | 64.2 ± 8.5 | 60.6 ± 7.8 | 65.9 ± 8.8 | 65.3 ± 8.2 | 65.9 ± 8.1 | 62.4 ± 7.7 | 66.0 ± 10.6 | 67.4 ± 8.5 | 67.9 ± 8.5 | 62.7 ± 7.6 | 69.1 ± 9.0 |
| Age [ᵻ] | | | | | | | | | | | | |
| < 67.2 yrs | 795 (64.9) | 662 (63.5) | 90 (80.4) | 25 (54.4) | 1,283 (58.3) | 1,004 (55.7) | 232 (71.6) | 16 (47.1) | 14,701 (48.8) | 12,422 (46.4) | 1,858 (72.0) | 150 (39.3) |
| ≥ 67.2 yrs | 430 (35.1) | 380 (36.5) | 22 (19.6) | 21 (45.7) | 917 (41.7) | 798 (44.3) | 92 (28.4) | 18 (52.9) | 15,419 (51.2) | 14,361 (53.6) | 724 (28.0) | 232 (60.7) |
| Educational attainment [g ᵻ] | | | | | | | | | | | | |
| <High school / GED | 225 (18.4) | 179 (17.2) | 26 (23.2) | 13 (28.3) | 180 (8.2) | 140 (7.8) | 31 (9.6) | 8 (23.5) | 4,057 (13.5) | 3,561 (13.3) | 371 (14.4) | 72 (18.9) |
| Some college / technical | 394 (32.2) | 329 (31.6) | 38 (33.9) | 15 (32.6) | 686 (31.2) | 548 (30.4) | 110 (34.0) | 12 (35.3) | 9,510 (31.6) | 8,369 (31.2) | 870 (33.7) | 141 (36.9) |
| ≥ Bachelor's or higher | 606 (49.5) | 534 (51.3) | 48 (42.9) | 18 (39.1) | 1,333 (60.6) | 1,113 (61.8) | 183 (56.5) | 14 (41.2) | 16,550 (54.9) | 14,851 (55.5) | 1,341 (51.9) | 169 (44.2) |
| *Missing* | *0 (0.0)* | *0 (0.0)* | *0 (0.0)* | *0 (0.0)* | *1 (0.1)* | *1 (0.1)* | *0 (0.0)* | *0 (0.0)* | *3 (0.0)* | *2 (0.0)* | *0 (0.0)* | *0 (0.0)* |
| Annual household income [ᵻ] | | | | | | | | | | | | |
| <$20,000 - $49,999 | 580 (47.4) | 483 (46.4) | 44 (39.3) | 37 (80.4) | 756 (34.4) | 632 (35.1) | 91 (28.1) | 18 (52.9) | 7,562 (25.1) | 6,832 (25.5) | 504 (19.5) | 141 (36.9) |
| $50,000-$99,999 | 351 (28.7) | 304 (29.2) | 34 (30.4) | 6 (13.0) | 839 (38.1) | 675 (37.5) | 141 (43.5) | 7 (20.6) | 11,187 (37.1) | 10,031 (37.5) | 896 (34.7) | 121 (31.7) |
| ≥$100,000 | 294 (24.0) | 255 (24.5) | 34 (30.4) | 3 (6.5) | 605 (27.5) | 495 (27.5) | 92 (28.4) | 9 (26.5) | 11,371 (37.8) | 9,920 (37.0) | 1,182 (45.8) | 120 (31.4) |
| Marital status [ᵻ] | | | | | | | | | | | | |
| Married or living as though married | 771 (62.9) | 645 (61.9) | 81 (72.3) | 26 (56.5) | 1,005 (45.7) | 790 (43.8) | 188 (58.0) | 10 (29.4) | 21,027 (69.8) | 18,434 (68.8) | 2,056 (79.6) | 260 (68.1) |
| Never married, divorced, widowed, or separated | 454 (37.1) | 397 (38.1) | 31 (27.7) | 20 (43.5) | 1,195 (54.3) | 1,012 (56.2) | 136 (42.0) | 24 (70.6) | 9,093 (30.2) | 8,349 (31.2) | 526 (20.4) | 122 (31.9) |
| Region of residence [ᵻ] | | | | | | | | | | | | |
| Northeast | 98 (8.0) | 79 (7.6) | 15 (13.4) | 1 (2.2) | 193 (8.8) | 159 (8.8) | 29 (9.0) | 2 (5.9) | 5,260 (17.5) | 4,697 (17.5) | 448 (17.4) | 61 (16.0) |
| Midwest | 77 (6.3) | 70 (6.7) | 7 (6.3) | 0 (0.0) | 451 (20.5) | 377 (20.9) | 61 (18.8) | 7 (20.6) | 8,457 (28.1) | 7,520 (28.1) | 724 (28.0) | 101 (26.4) |
| South | 349 (28.5) | 306 (29.4) | 30 (26.8) | 8 (17.4) | 1,379 (62.7) | 1,113 (61.8) | 216 (66.7) | 20 (58.8) | 9,370 (31.1) | 8,354 (31.2) | 782 (30.3) | 119 (31.2) |
| West | 335 (27.4) | 290 (27.8) | 33 (29.5) | 6 (13.0) | 177 (8.1) | 153 (8.5) | 18 (5.6) | 5 (14.7) | 7,029 (23.3) | 6,208 (23.2) | 628 (24.3) | 101 (26.4) |
| Puerto Rico | 366 (29.9) | 297 (28.5) | 27 (24.1) | 31 (67.4) | 0 (0.0) | 0 (0.0) | 0 (0.0) | 0 (0.0) | 4 (0.0) | 4 (0.0) | 0 (0.0) | 0 (0.0) |
| **Clinical characteristics** | | | | | | | | | | | | |
| Body Mass Index, $kg/m^2$ [ᵻ] | | | | | | | | | | | | |
| 18.5 - 24.9 | 386 (31.5) | 338 (32.4) | 31 (27.7) | 12 (26.1) | 373 (17.0) | 316 (17.5) | 46 (14.2) | 6 (17.7) | 12,369 (41.1) | 11,132 (41.6) | 911 (35.3) | 172 (45.0) |
| 25.0 - 29.9 | 443 (36.2) | 375 (36.0) | 37 (33.0) | 20 (43.5) | 741 (33.7) | 616 (34.2) | 99 (30.6) | 10 (29.4) | 9,583 (31.8) | 8,503 (31.8) | 877 (34.0) | 93 (24.4) |
| ≥30 | 383 (31.3) | 318 (30.5) | 43 (38.4) | 14 (30.4) | 1,063 (48.3) | 851 (47.2) | 178 (54.9) | 17 (50.0) | 7,774 (25.8) | 6,799 (25.4) | 760 (29.4) | 112 (29.3) |
| *Missing* | *13 (1.1)* | *11 (1.1)* | *1 (0.9)* | *0 (0.0)* | *23 (1.1)* | *19 (1.1)* | *1 (0.3)* | *1 (2.9)* | *394 (1.3)* | *349 (1.3)* | *34 (1.3)* | *5 (1.3)* |
| Menopausal status [ᵻ] | | | | | | | | | | | | |
| Premenopausal | 87 (7.1) | 79 (7.6) | 7 (6.3) | 1 (2.2) | 143 (6.5) | 104 (5.8) | 29 (9.0) | 5 (14.7) | 1,116 (3.7) | 927 (3.5) | 147 (5.7) | 19 (5.0) |
| Postmenopausal | 1,138 (92.9) | 963 (92.4) | 105 (93.8) | 45 (97.8) | 2,055 (93.4) | 1,696 (94.1) | 295 (91.1) | 29 (85.3) | 29,000 (96.3) | 25,852 (96.5) | 2,435 (94.3) | 363 (95.0) |

| | | | | | | | | | | | | |
|---|---|---|---|---|---|---|---|---|---|---|---|---|
| Missing or never had a period | 0 (0.0) | 0 (0.0) | 0 (0.0) | 0 (0.0) | 2 (0.1) | 2 (0.1) | 0 (0.0) | 0 (0.0) | 4 (0.0) | 4 (0.0) | 0 (0.0) | 0 (0.0) |
| Trouble sleeping for any other reason [h ǂ] | | | | | | | | | | | | |
| Yes | 783 (63.9) | 622 (59.7) | 100 (89.3) | 36 (78.3) | 1,494 (67.9) | 1,150 (63.8) | 276 (85.2) | 31 (91.2) | 21,576 (71.6) | 18,575 (69.4) | 2,306 (89.3) | 339 (88.7) |
| No | 442 (36.1) | 420 (40.3) | 12 (10.7) | 10 (21.7) | 706 (32.1) | 652 (36.2) | 48 (14.8) | 3 (8.8) | 8,543 (28.4) | 8,207 (30.6) | 276 (10.7) | 43 (11.3) |
| *Missing* | *0 (0.0)* | *0 (0.0)* | *0 (0.0)* | *0 (0.0)* | *0 (0.0)* | *0 (0.0)* | *0 (0.0)* | *0 (0.0)* | *1 (0.0)* | *1 (0.0)* | *0 (0.0)* | *0 (0.0)* |
| **Sleep health dimensions** | | | | | | | | | | | | |
| Average weekly sleep duration [i ǂ] | | | | | | | | | | | | |
| Short | 191 (15.6) | 160 (15.4) | 20 (17.9) | 6 (13.0) | 628 (28.6) | 504 (28.0) | 92 (28.4) | 14 (41.2) | 3,004 (10.0) | 2,618 (9.8) | 289 (11.2) | 56 (14.7) |
| Recommended | 799 (65.2) | 695 (66.7) | 65 (58.0) | 27 (58.7) | 1,274 (57.9) | 1,055 (58.6) | 188 (58.0) | 15 (44.1) | 22,322 (74.1) | 20,008 (74.7) | 1,829 (70.8) | 236 (61.8) |
| Long | 196 (16.0) | 157 (15.1) | 24 (21.4) | 8 (17.4) | 257 (11.7) | 212 (11.8) | 36 (11.1) | 4 (11.8) | 4,470 (14.8) | 3,865 (14.4) | 448 (17.4) | 79 (20.7) |
| *Missing* | *39 (3.2)* | *30 (2.9)* | *3 (2.7)* | *5 (10.9)* | *41 (1.9)* | *31 (1.7)* | *8 (2.5)* | *1 (2.9)* | *324 (1.1)* | *292 (1.1)* | *16 (0.6)* | *11 (2.9)* |
| Long sleep onset latency [j ǂ] | | | | | | | | | | | | |
| Yes | 216 (17.6) | 146 (14) | 41 (36.6) | 14 (30.4) | 327 (14.9) | 210 (11.7) | 90 (27.8) | 7 (20.6) | 3,640 (12.1) | 2,841 (10.6) | 558 (21.6) | 99 (25.9) |
| No | 968 (79.0) | 864 (82.9) | 67 (59.8) | 28 (60.9) | 1,834 (83.4) | 1,559 (86.5) | 229 (70.7) | 26 (76.5) | 26,150 (86.8) | 23,642 (88.3) | 2,006 (77.7) | 277 (72.5) |
| *Missing* | *41 (3.4)* | *32 (3.1)* | *4 (3.6)* | *4 (8.7)* | *39 (1.8)* | *33 (1.8)* | *5 (1.5)* | *1 (2.9)* | *330 (1.1)* | *300 (1.1)* | *18 (0.7)* | *6 (1.6)* |
| Poor sleep maintenance [k ǂ] | | | | | | | | | | | | |
| Yes | 410 (33.5) | 300 (28.8) | 68 (60.7) | 23 (50.0) | 793 (36.1) | 568 (31.5) | 179 (55.3) | 19 (55.9) | 12,057 (40.0) | 9,844 (36.8) | 1,717 (66.5) | 220 (57.6) |
| No | 777 (63.4) | 709 (68.0) | 43 (38.4) | 20 (43.5) | 1,372 (62.4) | 1,206 (66.9) | 139 (42.9) | 14 (41.2) | 17,728 (58.9) | 16,625 (62.1) | 850 (32.9) | 160 (41.9) |
| *Missing* | *38 (3.1)* | *33 (3.2)* | *1 (0.9)* | *3 (6.5)* | *35 (1.6)* | *28 (1.6)* | *6 (1.9)* | *1 (2.9)* | *335 (1.1)* | *314 (1.2)* | *15 (0.6)* | *2 (0.5)* |
| Insomnia symptoms [l ǂ] | | | | | | | | | | | | |
| Yes | 465 (38.0) | 342 (32.8) | 79 (70.5) | 25 (54.4) | 864 (39.3) | 616 (34.2) | 195 (60.2) | 21 (61.8) | 13,026 (43.3) | 10,663 (39.8) | 1,828 (70.8) | 233 (61.0) |
| No | 733 (59.8) | 678 (65.1) | 32 (28.6) | 18 (39.1) | 1,316 (59.8) | 1,169 (64.9) | 126 (38.9) | 13 (38.2) | 16,905 (56.1) | 15,940 (59.5) | 749 (29.0) | 147 (38.5) |
| *Missing* | *27 (2.2)* | *22 (2.1)* | *1 (0.9)* | *3 (6.5)* | *20 (0.9)* | *17 (0.9)* | *3 (0.9)* | *0 (0.0)* | *189 (0.6)* | *180 (0.7)* | *5 (0.2)* | *2 (0.5)* |
| Sleep medication use [m ǂ] | | | | | | | | | | | | |
| Yes | 180 (14.7) | 139 (13.3) | 18 (16.1) | 12 (26.1) | 242 (11.0) | 184 (10.2) | 44 (13.6) | 5 (14.7) | 5,868 (19.5) | 5,061 (18.9) | 599 (23.2) | 104 (27.2) |
| No | 1,023 (83.5) | 887 (85.1) | 94 (83.9) | 31 (67.4) | 1,939 (88.1) | 1,604 (89.0) | 276 (85.2) | 29 (85.3) | 24,131 (80.1) | 21,613 (80.7) | 1,973 (76.4) | 277 (72.5) |
| *Missing* | *22 (1.8)* | *16 (1.5)* | *0 (0.0)* | *3 (6.5)* | *19 (0.9)* | *14 (0.8)* | *4 (1.2)* | *0 (0.0)* | *121 (0.4)* | *109 (0.4)* | *10 (0.4)* | *1 (0.3)* |
| Daytime dysfunction [n] | | | | | | | | | | | | |
| Yes | 14 (1.1) | 9 (0.9) | 1 (0.9) | 2 (4.4) | 19 (0.9) | 11 (0.6) | 5 (1.5) | 3 (8.8) | 219 (0.7) | 166 (0.6) | 32 (1.2) | 11 (2.9) |
| No | 1,190 (97.1) | 1,015 (97.4) | 110 (98.2) | 43 (93.5) | 2,157 (98.1) | 1,771 (98.3) | 315 (97.2) | 31 (91.2) | 29,539 (98.1) | 26,288 (98.2) | 2,524 (97.8) | 368 (96.3) |
| *Missing* | *21 (1.7)* | *18 (1.7)* | *1 (0.9)* | *1 (2.2)* | *24 (1.1)* | *20 (1.1)* | *4 (1.2)* | *0 (0.0)* | *362 (1.2)* | *329 (1.2)* | *26 (1.0)* | *3 (0.8)* |
| Healthcare provider diagnosed sleep apnea [o ǂ] | | | | | | | | | | | | |
| Yes | 100 (8.2) | 76 (7.3) | 13 (11.6) | 8 (17.4) | 302 (13.7) | 229 (12.7) | 54 (16.7) | 10 (29.4) | 3,051 (10.1) | 2,649 (9.9) | 301 (11.7) | 58 (15.2) |
| No | 1,107 (90.4) | 953 (91.5) | 99 (88.4) | 36 (78.3) | 1,857 (84.4) | 1,539 (85.4) | 264 (81.5) | 23 (67.7) | 26,767 (88.9) | 23,863 (89.1) | 2,265 (87.7) | 314 (82.2) |
| *Missing* | *18 (1.5)* | *13 (1.3)* | *0 (0.0)* | *2 (4.4)* | *41 (1.9)* | *34 (1.9)* | *6 (1.9)* | *1 (2.9)* | *302 (1.0)* | *271 (1.0)* | *16 (0.6)* | *10 (2.6)* |

Data presented as mean ± standard deviation or n (%)

Note: N=438 (1.3%) participants who reported frequent trouble sleeping due to both ‘too hot’ and ‘too cold’ temperatures were not included in the analysis.

Abbreviations: SD, Standard Deviation; GED, General Educational Development

[¥]Chi-square test p-values indicate significant differences between the overall prevalence of ‘too hot’ by race and ethnicity.

[§]Chi-square test p-values indicate significant differences between the overall prevalence of ‘too cold’ by race and ethnicity.

[ǂ]Chi-square or ANOVA test p-values indicate significant differences in characteristics between the total populations of each race and ethnicity.

[a] Excluded for reporting both ‘Too hot’ and ‘Too cold’, Hispanic/Latina: n = 25 (2.0%)
[b] Excluded for reporting both ‘Too hot’ and ‘Too cold’, Non-Hispanic Black: n = 40 (1.8%)
[c] Excluded for reporting both ‘Too hot’ and ‘Too cold’, Non-Hispanic White: n = 373 (1.2%)
[d] ‘Infrequent temperature extremes’ is defined as self-reported trouble sleeping due to feeling too hot < 3 times per week and trouble sleeping due to feeling too cold < 3 times per week in the past month.
[e] ’Too hot’ is defined as self-reported trouble sleeping due to feeling too hot ≥ 3 times per week in the past month.
[f] ’Too cold’ is defined as self-reported trouble sleeping due to feeling too cold ≥ 3 times per week in the past month.
[g] Educational attainment was assessed at baseline.
[h] Reasons for trouble sleeping ≥ 3 times per week in the past month include: unable to fall asleep within 30 minutes, waking up in the middle of the night or early morning, waking up to use the bathroom, cannot breathe comfortably, coughing or snoring loudly, having bad dreams, having pain, or other non-specified reasons.
[i] Sleep duration is based on reported bed and wake times or reported average sleep duration. Participants who reported ≤ 2 hours or ≥ 23 hours of sleep are excluded. Short: <7 hours; Recommended: 7-9 hours; Long >9 hours.
[j] Long sleep onset latency is defined as not falling asleep within 30 minutes at least three times a week during the past month.
[k] Poor sleep maintenance is defined as waking up in the middle of the night or early morning at least three times a week during the past month.
[l] Insomnia symptoms is defined as long sleep onset latency or poor sleep maintenance.
[m] Sleep medication use is defined as 'taking medicine (prescription or over the counter) to help you sleep' at least three times a week during the past month.
[n] Daytime dysfunction is defined as having trouble staying awake while driving, eating, or engaging in social activity at least three times a week during the past month.
[o] Healthcare provider diagnosed sleep apnea is defined as a current doctor or other health professional diagnosis of sleep apnea.

**Supplemental Table 3. Sociodemographic characteristics, clinical characteristics, and sleep health dimensions among eligible Sister Study participants by annual household income as well as perceived indoor temperature extremes, Sister Study, 2017-2019, N = 33,545**

| Annual household income | <$20,000 - $49,999 [a] | | | | $50,000 - $99,999 [b] | | | | ≥ $100,000[c] | | | |
|---|---|---|---|---|---|---|---|---|---|---|---|---|
| | Total n = 8,898 (26.5%) | Infrequent temperature extremes [d] n = 7,947 (89.3%) | Too hot [e ¥] n = 639 (7.2%) | Too cold [f §] n = 196 (2.2%) | Total n = 12,377 (36.9%) | Infrequent temperature extremes [d] n = 11,010 (89.0%) | Too hot [e ¥] n = 1,071 (8.7%) | Too cold [f §] n = 134 (1.1 %) | Total n = 12,270 (36.6%) | Infrequent temperature extremes [d] n = 10, 670 (87.0%) | Too hot [e ¥] n = 1,308 (10.7%) | Too cold [f §] n = 132 (1.1%) |
| **Sociodemographic characteristics** | | | | | | | | | | | | |
| Age, years (mean ± SD) [ᵻ] | 70.5 ± 8.2 | 71.0 ± 8.1 | 65.2 ± 7.9 | 71.0 ± 9.0 | 67.8 ± 8.0 | 68.2 ± 8.0 | 63.7 ± 7.3 | 67.9 ± 8.4 | 64.0 ± 8.2 | 64.5 ±8.2 | 60.4 ± 7.1 | 65.5 ± 9.1 |
| Age [ᵻ] | | | | | | | | | | | | |
| < 67.2 yrs | 3,011 (33.8) | 2,494 (31.4) | 382 (59.8) | 64 (32.7) | 5,756 (46.5) | 4,863 (44.2) | 720 (67.2) | 58 (43.3) | 8,012 (65.3) | 6,731 (63.1) | 1,078 (82.4) | 69 (52.3) |
| ≥ 67.2 yrs | 5,887 (66.2) | 5,453 (68.6) | 257 (40.2) | 132 (67.4) | 6,621 (53.5) | 6,147 (55.8) | 351 (32.8) | 76 (56.7) | 4,258 (34.7) | 3,939 (36.9) | 230 (17.6) | 63 (47.7) |
| Race and ethnicity [ᵻ] | | | | | | | | | | | | |
| Hispanic/Latina | 580 (6.5) | 483 (6.1) | 44 (6.9) | 37 (18.9) | 351 (2.8) | 304 (2.8) | 34 (3.2) | 6 (4.5) | 294 (2.4) | 255 (2.4) | 34 (2.6) | 3 (2.3) |
| Non-Hispanic Black | 756 (8.5) | 632 (8.0) | 91 (14.2) | 18 (9.2) | 839 (6.8) | 675 (6.1) | 141 (13.2) | 7 (5.2) | 605 (4.9) | 495 (4.6) | 92 (7.0) | 9 (6.8) |
| Non-Hispanic White | 7,562 (85.0) | 6,832 (86.0) | 504 (78.9) | 141 (71.9) | 11,187 (90.4) | 10,031 (91.1) | 896 (83.7) | 121 (90.3) | 11,371 (92.7) | 9,920 (93.0) | 1,182 (90.4) | 120 (90.9) |
| *Missing* | *0 (0.0)* | *0 (0.0)* | *0 (0.0)* | *0 (0.0)* | *0 (0.0)* | *0 (0.0)* | *0 (0.0)* | *0 (0.0)* | *0 (0.0)* | *0 (0.0)* | *0 (0.0)* | *0 (0.0)* |
| Educational attainment [g ᵻ] | | | | | | | | | | | | |
| <High school / GED | 2,160 (24.3) | 1,893 (23.8) | 176 (27.5) | 61 (31.1) | 1,615 (13.1) | 1,412 (12.8) | 159 (14.9) | 21 (15.7) | 687 (5.6) | 575 (5.4) | 93 (7.1) | 11 (8.3) |
| Some college / technical | 3,625 (40.7) | 3,240 (40.8) | 257 (40.2) | 79 (40.3) | 4,189 (33.9) | 3,648 (33.1) | 421 (39.3) | 53 (39.6) | 2,776 (22.6) | 2,358 (22.1) | 340 (26.0) | 36 (27.3) |
| ≥ Bachelor's or higher | 3,111 (35.0) | 2,812 (35.4) | 206 (32.2) | 56 (28.6) | 6,572 (53.1) | 5,949 (54.0) | 491 (45.9) | 60 (44.8) | 8,806 (71.8) | 7,737 (72.5) | 875 (66.9) | 85 (64.4) |
| *Missing* | *2 (0.0)* | *2 (0.0)* | *0 (0.0)* | *0 (0.0)* | *1 (0.0)* | *1 (0.0)* | *0 (0.0)* | *0 (0.0)* | *1 (0.0)* | *0 (0.0)* | *0 (0.0)* | *0 (0.0)* |
| Marital status [ᵻ] | | | | | | | | | | | | |
| Married or living as though married | 3,393 (38.1) | 2,966 (37.3) | 303 (47.4) | 78 (39.8) | 8,645 (69.8) | 7,585 (68.9) | 834 (77.9) | 101 (75.4) | 10,765 (87.7) | 9,318 (87.3) | 1,188 (90.8) | 117 (88.6) |
| Never married, divorced, widowed, or separated | 5,505 (61.9) | 4,981 (62.7) | 336 (52.6) | 118 (60.2) | 3,732 (30.2) | 3,425 (31.1) | 237 (22.1) | 33 (24.6) | 1,505 (12.3) | 1,352 (12.7) | 120 (9.2) | 15 (11.4) |
| Region of residence [ᵻ] | | | | | | | | | | | | |
| Northeast | 1,246 (14.0) | 1,127 (14.2) | 89 (13.9) | 20 (10.2) | 1,935 (15.6) | 1,761 (16.0) | 134 (12.5) | 22 (16.4) | 2,370 (19.3) | 2,047 (19.2) | 269 (20.6) | 22 (16.7) |
| Midwest | 2,525 (28.4) | 2,283 (28.7) | 162 (25.4) | 52 (26.5) | 3,565 (28.8) | 3,171 (28.8) | 313 (29.2) | 30 (22.4) | 2,895 (23.6) | 2,513 (23.6) | 317 (24.2) | 26 (19.7) |
| South | 2,946 (33.1) | 2,619 (33.0) | 231 (36.2) | 52 (26.5) | 4,089 (33.0) | 3,609 (32.8) | 376 (35.1) | 46 (34.3) | 4,063 (33.1) | 3,545 (33.2) | 421 (32.2) | 49 (37.1) |
| West | 1,884 (21.2) | 1,681 (21.2) | 135 (21.1) | 44 (22.5) | 2,731 (22.1) | 2,419 (22.0) | 245 (22.9) | 33 (24.6) | 2,926 (23.9) | 2,551 (23.9) | 299 (22.9) | 35 (26.5) |
| Puerto Rico | 297 (3.3) | 237 (3.0) | 22 (3.4) | 28 (14.3) | 57 (0.5) | 50 (0.5) | 3 (0.3) | 3 (2.2) | 16 (0.1) | 14 (0.1) | 2 (0.2) | 0 (0.0) |
| **Clinical characteristics** | | | | | | | | | | | | |
| Body Mass Index, kg/$m^2$ [ᵻ] | | | | | | | | | | | | |
| 18.5 - 24.9 | 2,854 (32.1) | 2,605 (32.8) | 158 (24.7) | 65 (33.2) | 4,679 (37.8) | 4,220 (38.3) | 338 (31.6) | 62 (46.3) | 5,595 (45.6) | 4,961 (46.5) | 492 (37.6) | 63 (47.7) |
| 25.0 - 29.9 | 2,852 (32.1) | 2,546 (32.0) | 211 (33.0) | 57 (29.1) | 4,064 (32.8) | 3,633 (33.0) | 354 (33.1) | 30 (22.4) | 3,851 (31.4) | 3,315 (31.1) | 448 (34.3) | 36 (27.3) |
| ≥30 | 3,067 (34.5) | 2,681 (33.7) | 265 (41.5) | 71 (36.2) | 3,469 (28.0) | 3,014 (27.4) | 363 (33.9) | 41 (30.6) | 2,684 (21.9) | 2,273 (21.3) | 353 (27.0) | 31 (23.5) |
| *Missing* | *125 (1.4)* | *115 (1.5)* | *5 (0.8)* | *3 (1.5)* | *165 (1.3)* | *143 (1.3)* | *16 (1.5)* | *1 (0.8)* | *140 (1.1)* | *121 (1.1)* | *15 (1.2)* | *2 (1.5)* |
| Menopausal status [ᵻ] | | | | | | | | | | | | |

| | | | | | | | | | | | | |
|---|---|---|---|---|---|---|---|---|---|---|---|---|
| Premenopausal | 144 (1.6) | 116 (1.5) | 22 (3.4) | 1 (0.5) | 345 (2.8) | 272 (2.5) | 53 (5.0) | 9 (6.7) | 857 (7.0) | 722 (6.8) | 108 (8.3) | 15 (11.4) |
| Postmenopausal | 8,751 (98.4) | 7,828 (98.5) | 617 (96.6) | 195 (99.5) | 12,031 (97.2) | 10,737 (97.5) | 1,018 (95.1) | 125 (93.3) | 11,411 (93.0) | 9,946 (93.2) | 1,200 (91.7) | 117 (88.6) |
| *Missing or never had a period* | *3 (0.0)* | *3 (0.0)* | *0 (0.0)* | *0 (0.0)* | *1 (0.0)* | *1 (0.0)* | *0 (0.0)* | *0 (0.0)* | *2 (0.0)* | *2 (0.0)* | *0 (0.0)* | *0 (0.0)* |
| Trouble sleeping for any other reason [h ǂ] | | | | | | | | | | | | |
| Yes | 6,614 (74.3) | 5,741 (72.2) | 585 (91.6) | 176 (89.8) | 8,820 (71.3) | 7,605 (69.1) | 943 (88.1) | 119 (88.8) | 8,419 (68.6) | 7,001 (65.6) | 1,154 (88.2) | 111 (84.1) |
| No | 2,284 (25.7) | 2,206 (27.8) | 54 (8.5) | 20 (10.2) | 3,556 (28.7) | 3,404 (30.9) | 128 (12.0) | 15 (11.2) | 3,851 (31.4) | 3,669 (34.4) | 154 (11.8) | 21 (15.9) |
| *Missing* | *0 (0.0)* | *0 (0.0)* | *0 (0.0)* | *0 (0.0)* | *1 (0.0)* | *1 (0.0)* | *0 (0.0)* | *0 (0.0)* | *0 (0.0)* | *0 (0.0)* | *0 (0.0)* | *0 (0.0)* |
| **Sleep health dimensions** | | | | | | | | | | | | |
| Average weekly sleep duration [i ǂ] | | | | | | | | | | | | |
| Short | 1,186 (13.3) | 1,035 (13.0) | 93 (14.6) | 36 (18.4) | 1,390 (11.2) | 1,184 (10.8) | 161 (15.0) | 22 (16.4) | 1,247 (10.2) | 1,063 (10.0) | 147 (11.2) | 18 (13.6) |
| Recommended | 5,902 (66.3) | 5,345 (67.3) | 399 (62.4) | 104 (53.1) | 9,070 (73.3) | 8,144 (74.0) | 732 (68.4) | 91 (67.9) | 9,423 (76.8) | 8,269 (77.5) | 951 (72.7) | 83 (62.9) |
| Long | 1,615 (18.2) | 1,403 (17.7) | 134 (21.0) | 44 (22.5) | 1,801 (14.6) | 1,576 (14.3) | 171 (16.0) | 19 (14.2) | 1,507 (12.3) | 1,255 (11.8) | 203 (15.5) | 28 (21.2) |
| *Missing* | *195 (2.2)* | *164 (2.1)* | *13 (2.0)* | *12 (6.1)* | *116 (0.9)* | *106 (1.0)* | *7 (0.7)* | *2 (1.5)* | *93 (0.8)* | *83 (0.8)* | *7 (0.5)* | *3 (2.3)* |
| Long sleep onset latency [j ǂ] | | | | | | | | | | | | |
| Yes | 1,420 (16.0) | 1,106 (13.9) | 192 (30.1) | 64 (32.7) | 1,561 (12.6) | 1,186 (10.8) | 274 (25.6) | 31 (23.1) | 1,202 (9.8) | 905 (8.5) | 223 (17.1) | 25 (18.9) |
| No | 7,313 (82.2) | 6,697 (84.3) | 436 (68.2) | 125 (63.8) | 10,685 (86.3) | 9,702 (88.1) | 791 (73.9) | 102 (76.1) | 10,954 (89.3) | 9,666 (90.6) | 1,075 (82.2) | 104 (78.8) |
| *Missing* | *165 (1.9)* | *144 (1.8)* | *11 (1.7)* | *7 (3.6)* | *131 (1.1)* | *122 (1.1)* | *6 (0.6)* | *1 (0.8)* | *114 (0.9)* | *99 (0.9)* | *10 (0.8)* | *3 (2.3)* |
| Poor sleep maintenance [k ǂ] | | | | | | | | | | | | |
| Yes | 3,634 (40.8) | 3,011 (37.9) | 426 (66.7) | 112 (57.1) | 4,832 (39.0) | 3,957 (35.9) | 677 (63.2) | 82 (61.2) | 4,794 (39.1) | 3,744 (35.1) | 861 (65.8) | 68 (51.5) |
| No | 5,109 (57.4) | 4,791 (60.3) | 211 (33.0) | 79 (40.3) | 7,406 (59.8) | 6,925 (62.9) | 386 (36.0) | 51 (38.1) | 7,362 (60.0) | 6,824 (64.0) | 435 (33.3) | 64 (48.5) |
| *Missing* | *155 (1.7)* | *145 (1.8)* | *2 (0.3)* | *5 (2.6)* | *139 (1.1)* | *128 (1.2)* | *8 (0.8)* | *1 (0.8)* | *114 (0.9)* | *102 (1)* | *12 (0.9)* | *0 (0.0)* |
| Insomnia symptoms [l ǂ] | | | | | | | | | | | | |
| Yes | 3,961 (44.5) | 3,288 (41.4) | 458 (71.7) | 120 (61.2) | 5,263 (42.5) | 4,316 (39.2) | 731 (68.3) | 85 (63.4) | 5,131 (41.8) | 4,017 (37.7) | 913 (69.8) | 74 (56.1) |
| No | 4,850 (54.5) | 4,580 (57.6) | 180 (28.2) | 72 (36.7) | 7,031 (56.8) | 6,615 (60.1) | 337 (31.5) | 48 (35.8) | 7,073 (57.6) | 6,592 (61.8) | 390 (29.8) | 58 (43.9) |
| *Missing* | *87 (1.0)* | *79 (1.0)* | *1 (0.2)* | *4 (2.0)* | *83 (0.7)* | *79 (0.7)* | *3 (0.3)* | *1 (0.8)* | *66 (0.5)* | *61 (0.6)* | *5 (0.4)* | *0 (0.0)* |
| Sleep medication use [m ǂ] | | | | | | | | | | | | |
| Yes | 1,839 (20.7) | 1,571 (19.8) | 170 (26.6) | 57 (29.1) | 2,370 (19.2) | 2,053 (18.7) | 242 (22.6) | 27 (20.2) | 2,081 (17.0) | 1,760 (16.5) | 249 (19.0) | 37 (28.0) |
| No | 6,984 (78.5) | 6,312 (79.4) | 465 (72.8) | 136 (69.4) | 9,955 (80.4) | 8,913 (81.0) | 822 (76.8) | 106 (79.1) | 10,154 (82.8) | 8,879 (83.2) | 1,056 (80.7) | 95 (72.0) |
| *Missing* | *75 (0.8)* | *64 (0.8)* | *4 (0.6)* | *3 (1.5)* | *52 (0.4)* | *44 (0.4)* | *7 (0.7)* | *1 (0.8)* | *35 (0.3)* | *31 (0.3)* | *3 (0.2)* | *0 (0.0)* |
| Daytime dysfunction [n ǂ] | | | | | | | | | | | | |
| Yes | 89 (1.0) | 69 (0.9) | 11 (1.7) | 5 (2.6) | 87 (0.7) | 63 (0.6) | 14 (1.3) | 7 (5.2) | 76 (0.6) | 54 (0.5) | 13 (1.0) | 4 (3.0) |
| No | 8,677 (97.5) | 7,755 (97.6) | 623 (97.5) | 189 (96.4) | 12,136 (98.1) | 10,810 (98.2) | 1,042 (97.3) | 126 (94.0) | 12,073 (98.4) | 10,509 (98.5) | 1,284 (98.2) | 127 (96.2) |
| *Missing* | *132 (1.5)* | *123 (1.6)* | *5 (0.8)* | *2 (1.0)* | *154 (1.2)* | *137 (1.2)* | *15 (1.4)* | *1 (0.8)* | *121 (1.0)* | *107 (1.0)* | *11 (0.8)* | *1 (0.8)* |
| Healthcare provider diagnosed sleep apnea [o ǂ] | | | | | | | | | | | | |
| Yes | 1,156 (13.0) | 998 (12.6) | 106 (16.6) | 36 (18.4) | 1,300 (10.5) | 1,114 (10.1) | 136 (12.7) | 22 (16.4) | 997 (8.1) | 842 (7.9) | 126 (9.6) | 18 (13.6) |
| No | 7,595 (85.4) | 6,824 (85.9) | 523 (81.9) | 153 (78.1) | 10,947 (88.5) | 9,778 (88.8) | 931 (86.9) | 106 (79.1) | 11,189 (91.2) | 9,753 (91.4) | 1,174 (89.8) | 114 (86.4) |
| *Missing* | *147 (1.7)* | *125 (1.6)* | *10 (1.6)* | *7 (3.6)* | *130 (1.1)* | *118 (1.1)* | *4 (0.4)* | *6 (4.5)* | *84 (0.7)* | *75 (0.7)* | *8 (0.6)* | *0 (0.0)* |

Data presented as mean ± standard deviation or n (%)
Note: N=438 (1.3%) participants who reported both 'too hot' and 'too cold' temperatures were not included in the analysis.
Abbreviations: SD, Standard Deviation; GED, General Educational Development

[¥] Chi-square test p-values indicate significant differences in the overall prevalence of ‘too hot’ by annual household income category.
[§] Chi-square test p-values indicate significant differences in the overall prevalence of ‘too cold’ by annual household income category.
[ǂ] Chi-square or ANOVA test p-values indicate significant differences in characteristics between the total populations of each annual household income category.
[a] Excluded for reporting both ‘Too hot’ and ‘Too cold’, < $20,000 - $49,999: n = 116 (1.3%)
[b] Excluded for reporting both ‘Too hot’ and ‘Too cold’, $50,000 - $99,999: n = 162 (1.3%)
[c] Excluded for reporting both ‘Too hot’ and ‘Too cold’, ≥ $100,000: n = 160 (1.3%)
[d] Infrequent temperature extremes’ is defined as self-reported trouble sleeping due to feeling too hot < 3 times per week and trouble sleeping due to feeling too cold < 3 times per week in the past month.
[e] ’Too hot’ is defined as self-reported trouble sleeping due to feeling too hot ≥ 3 times per week in the past month.
[f] ’Too cold’ is defined as self-reported trouble sleeping due to feeling too cold ≥ 3 times per week in the past month.
[g] Educational attainment was assessed at baseline.
[h] Reasons for trouble sleeping ≥ 3 times per week in the past month include: unable to fall asleep within 30 minutes, waking up in the middle of the night or early morning, waking up to use the bathroom, cannot breathe comfortably, coughing or snoring loudly, having bad dreams, having pain, or other non-specified reasons.
[i] Sleep duration is based on reported bed and wake times or reported average sleep duration. Participants who reported ≤ 2 hours or ≥ 23 hours of sleep are excluded. Short: <7 hours; Recommended: 7-9 hours; Long >9 hours
[j] Long sleep onset latency is defined as not falling asleep within 30 minutes at least three times a week during the past month.
[k] Poor sleep maintenance is defined as waking up in the middle of the night or early morning at least three times a week during the past month.
[l] Insomnia symptoms is defined as having long sleep onset latency or poor sleep maintenance.
[m] Sleep medication use is defined as 'taking medicine (prescription or over the counter) to help you sleep' at least three times a week during the past month.
[n] Daytime dysfunction is defined as having trouble staying awake while driving, eating, or engaging in social activity at least three times a week during the past month.
[o] Healthcare provider diagnosed sleep apnea is defined as a current doctor or other health professional diagnosis of sleep apnea.

**Supplemental Table 4. Sociodemographic characteristics, clinical characteristics, and sleep health dimensions among eligible Sister Study participants by region of residence as well as perceived indoor temperature extremes, Sister Study, 2017-2019, N = 33,545**

| Region | Northeast [a] | | | | Midwest [b] | | | | South [c] | | | | West [d] | | | | Puerto Rico [e] | | | |
|---|---|---|---|---|---|---|---|---|---|---|---|---|---|---|---|---|---|---|---|---|
| | Total n = 5,551 (16.6%) | Infrequent temperature extremes [f] n = 4,935 (88.9%) | Too hot[g¥] n = 492 (8.9%) | Too cold[h§] n = 64 (1.2%) | Total n = 8,985 (26.8%) | Infrequent temperature extremes [f] n = 7,967 (88.7%) | Too hot[g¥] n = 792 (8.8%) | Too cold[h§] n = 108 (1.2%) | Total n = 11,098 (33.1%) | Infrequent temperature extremes [f] n = 9,773 (88.1%) | Too hot[g¥] n = 1,028 (9.3%) | Too cold[h§] n = 147 (1.3%) | Total n = 7,541 (22.5%) | Infrequent temperature extremes [f] n = 6,651 (88.2%) | Too hot[g¥] n = 679 (9.0%) | Too cold[h§] n = 112 (1.5%) | Total n = 370 (1.1%) | Infrequent temperature extremes [f] n = 301 (81.4%) | Too hot[g¥] n = 27 (7.3%) | Too cold [h§] n = 31 (8.4%) |
| **Sociodemographic characteristics** | | | | | | | | | | | | | | | | | | | | |
| Age, years (mean ± SD) [ǂ] | 66.7 ± 8.7 | 67.3 ± 8.6 | 61.5 ± 7.3 | 67.9 ± 8.6 | 66.8 ± 8.5 | 67.3 ± 8.5 | 62.1 ± 7.4 | 68.0 ± 9.3 | 67.2 ± 8.5 | 67.7 ± 8.4 | 62.9 ± 7.7 | 68.4 ± 9.7 | 67.7 ± 8.5 | 68.2 ± 8.4 | 63.5 ± 7.8 | 70.0 ± 8.7 | 65.8 ± 8.2 | 65.9 ± 8.2 | 64.3 ± 7.5 | 66.8 ± 8.3 |
| Age [ǂ] | | | | | | | | | | | | | | | | | | | | |
| < 67.2 yrs | 2,898 (52.2) | 2,434 (49.3) | 389 (79.1) | 26 (40.6) | 4,630 (51.5) | 3,896 (48.9) | 597 (75.4) | 48 (44.4) | 5,480 (49.4) | 4,590 (47.0) | 717 (69.8) | 63 (42.9) | 3,567 (47.3) | 3,002 (45.1) | 460 (67.8) | 38 (33.9) | 204 (55.1) | 166 (55.2) | 17 (63.0) | 16 (51.6) |
| ≥ 67.2 yrs | 2,653 (47.8) | 2,501 (50.7) | 103 (20.9) | 38 (59.4) | 4,355 (48.5) | 4,071 (51.1) | 195 (24.6) | 60 (55.6) | 5,618 (50.6) | 5,183 (53.0) | 311 (30.3) | 84 (57.1) | 3,974 (52.7) | 3,649 (54.9) | 219 (32.3) | 74 (66.1) | 166 (44.9) | 135 (44.9) | 10 (37.0) | 15 (48.4) |
| Race and ethnicity [ǂ] | | | | | | | | | | | | | | | | | | | | |
| Hispanic/Latina | 98 (1.8) | 79 (1.6) | 15 (3.1) | 1 (1.6) | 77 (0.9) | 70 (0.9) | 7 (0.9) | 0 (0.0) | 349 (3.1) | 306 (3.1) | 30 (2.9) | 8 (5.4) | 335 (4.4) | 290 (4.4) | 33 (4.9) | 6 (5.4) | 366 (98.9) | 297 (98.7) | 27 (100.0) | 31 (100.0) |
| Non-Hispanic Black | 193 (3.5) | 159 (3.2) | 29 (5.9) | 2 (3.1) | 451 (5.0) | 377 (4.7) | 61 (7.7) | 7 (6.5) | 1,379 (12.4) | 1,113 (11.4) | 216 (21.0) | 20 (13.6) | 177 (2.4) | 153 (2.3) | 18 (2.7) | 5 (4.5) | 4 (1.1) | 0 (0.0) | 0 (0.0) | 0 (0.0) |
| Non-Hispanic White | 5,260 (94.8) | 4,697 (95.2) | 448 (91.1) | 61 (95.3) | 8,457 (94.1) | 7,520 (94.4) | 724 (91.4) | 101 (93.5) | 9,370 (84.4) | 8,354 (85.5) | 782 (76.1) | 119 (81.0) | 7,029 (93.2) | 6,208 (93.3) | 628 (92.5) | 101 (90.2) | 4 (1.1) | 4 (1.3) | 0 (0.0) | 0 (0.0) |
| Educational attainment [i ǂ] | | | | | | | | | | | | | | | | | | | | |
| <High school / GED | 750 (13.5) | 656 (13.3) | 71 (14.4) | 15 (23.4) | 1,459 (16.2) | 1,277 (16.0) | 143 (18.1) | 24 (22.2) | 1,399 (12.6) | 1,229 (12.6) | 123 (12.0) | 30 (20.4) | 771 (10.2) | 658 (9.9) | 80 (11.8) | 16 (14.3) | 83 (22.4) | 60 (19.9) | 11 (40.7) | 8 (25.8) |
| Some college / technical | 1,541 (27.8) | 1,349 (27.3) | 149 (30.3) | 22 (34.4) | 2,997 (33.4) | 2,643 (33.2) | 262 (33.1) | 46 (42.6) | 3,507 (31.6) | 3,047 (31.2) | 354 (34.4) | 52 (35.4) | 2,446 (32.4) | 2,130 (32.0) | 246 (36.2) | 38 (33.9) | 99 (26.8) | 77 (25.6) | 7 (25.9) | 10 (32.3) |
| ≥ Bachelor's or higher | 3,258 (58.7) | 2,929 (59.4) | 272 (55.3) | 27 (42.2) | 4,529 (50.4) | 4,047 (50.8) | 387 (48.9) | 38 (35.2) | 6,190 (55.8) | 5,495 (56.2) | 551 (53.6) | 65 (44.2) | 4,324 (57.3) | 3,863 (58.1) | 353 (52.0) | 58 (51.8) | 188 (50.8) | 164 (54.5) | 9 (33.3) | 13 (41.9) |
| *Missing* | *2 (0.0)* | *1 (0.0)* | *0 (0.0)* | *0 (0.0)* | *0 (0.0)* | *0 (0.0)* | *0 (0.0)* | *0 (0.0)* | *2 (0.0)* | *2 (0.0)* | *0 (0.0)* | *0 (0.0)* | *0 (0.0)* | *0 (0.0)* | *0 (0.0)* | *0 (0.0)* | *0 (0.0)* | *0 (0.0)* | *0 (0.0)* | *0 (0.0)* |
| Annual household income [ǂ] | | | | | | | | | | | | | | | | | | | | |
| < $20,000 - $49,999 | 1,246 (22.5) | 1,127 (22.8) | 89 (18.1) | 20 (31.3) | 2,525 (28.1) | 2,283 (28.7) | 162 (20.5) | 52 (48.2) | 2,946 (26.6) | 2,619 (26.8) | 231 (22.5) | 52 (35.4) | 1,884 (25.0) | 1,681 (25.3) | 135 (19.9) | 44 (39.3) | 297 (80.3) | 237 (78.7) | 22 (81.5) | 28 (90.3) |
| $50,000 – $99,999 | 1,935 (34.9) | 1,761 (35.7) | 134 (27.2) | 22 (34.4) | 3,565 (39.7) | 3,171 (39.8) | 313 (39.5) | 30 (27.8) | 4,089 (36.8) | 3,609 (36.9) | 376 (36.6) | 46 (31.3) | 2,731 (36.2) | 2,419 (36.4) | 245 (36.1) | 33 (29.5) | 57 (15.4) | 50 (16.6) | 3 (11.1) | 3 (9.7) |
| ≥ $100,000 | 2,370 (42.7) | 2,047 (41.5) | 269 (54.7) | 22 (34.4) | 2,895 (32.2) | 2,513 (31.5) | 317 (40.0) | 26 (24.1) | 4,063 (36.6) | 3,545 (36.3) | 421 (41.0) | 49 (33.3) | 2,926 (38.8) | 2,551 (38.4) | 299 (44.0) | 35 (31.3) | 16 (4.3) | 14 (4.7) | 2 (7.4) | 0 (0.0) |
| Marital status [ǂ] | | | | | | | | | | | | | | | | | | | | |
| Married/living as though married | 3,805 (68.5) | 3,327 (67.4) | 387 (78.7) | 47 (73.4) | 6,277 (69.9) | 5,497 (69.0) | 623 (78.7) | 70 (64.8) | 7,426 (66.9) | 6,459 (66.1) | 774 (75.3) | 94 (63.9) | 5,071 (67.2) | 4,405 (66.2) | 523 (77.0) | 68 (60.7) | 224 (60.5) | 181 (60.1) | 18 (66.7) | 17 (54.8) |
| Never married, divorced, widowed, or separated | 1,746 (31.5) | 1,608 (32.6) | 105 (21.3) | 17 (26.6) | 2,708 (30.1) | 2,470 (31.0) | 169 (21.3) | 38 (35.2) | 3,672 (33.1) | 3,314 (33.9) | 254 (24.7) | 53 (36.1) | 2,470 (32.8) | 2,246 (33.8) | 156 (23.0) | 44 (39.3) | 146 (39.5) | 120 (39.9) | 9 (33.3) | 14 (45.2) |
| **Clinical characteristics** | | | | | | | | | | | | | | | | | | | | |
| Body Mass Index, kg/m$^2$ [ǂ] | | | | | | | | | | | | | | | | | | | | |
| 18.5 - 24.9 | 2,300 (41.4) | 2,080 (42.2) | 168 (34.2) | 23 (35.9) | 3,224 (35.9) | 2,889 (36.3) | 251 (31.7) | 44 (40.7) | 4,159 (37.5) | 3,767 (38.5) | 290 (28.2) | 55 (37.4) | 3,333 (44.2) | 2,961 (44.5) | 269 (39.6) | 59 (52.7) | 112 (30.3) | 89 (29.6) | 10 (37.0) | 9 (29.0) |
| 25.0 - 29.9 | 1,745 (31.4) | 1,556 (31.5) | 154 (31.3) | 17 (26.6) | 2,915 (32.4) | 2,572 (32.3) | 272 (34.3) | 30 (27.8) | 3,619 (32.6) | 3,182 (32.6) | 349 (34.0) | 38 (25.9) | 2,339 (31.0) | 2,061 (31.0) | 229 (33.7) | 25 (22.3) | 149 (40.3) | 123 (40.9) | 9 (33.3) | 13 (41.9) |

| | | | | | | | | | | | | | | | | | | | | |
|---|---|---|---|---|---|---|---|---|---|---|---|---|---|---|---|---|---|---|---|---|
| ≥30 | 1,434 (25.8) | 1,235 (25.0) | 164 (33.3) | 23 (35.9) | 2,738 (30.5) | 2,410 (30.3) | 257 (32.5) | 34 (31.5) | 3,163 (28.5) | 2,686 (27.5) | 378 (36.8) | 51 (34.7) | 1,780 (23.6) | 1,551 (23.3) | 174 (25.6) | 26 (23.2) | 105 (28.4) | 86 (28.6) | 8 (29.6) | 9 (29.0) |
| *Missing* | *72 (1.3)* | *64 (1.3)* | *6 (1.2)* | *1 (1.6)* | *108 (1.2)* | *96 (1.2)* | *12 (1.5)* | *0 (0.0)* | *157 (1.4)* | *138 (1.4)* | *11 (1.1)* | *3 (2.0)* | *89 (1.2)* | *78 (1.2)* | *7 (1.0)* | *2 (1.8)* | *4 (1.1)* | *3 (1.0)* | *0 (0.0)* | *0 (0.0)* |
| Menopausal status | | | | | | | | | | | | | | | | | | | | |
| Premenopausal | 230 (4.1) | 192 (3.9) | 31 (6.3) | 2 (3.1) | 376 (4.2) | 314 (3.9) | 50 (6.3) | 7 (6.5) | 437 (3.9) | 350 (3.6) | 65 (6.3) | 10 (6.8) | 282 (3.7) | 235 (3.5) | 36 (5.3) | 5 (4.5) | 21 (5.7) | 19 (6.3) | 1 (3.7) | 1 (3.2) |
| Postmenopausal | 5,321 (95.9) | 4,743 (96.1) | 461 (93.7) | 62 (96.9) | 8,608 (95.8) | 7,652 (96.1) | 742 (93.7) | 101 (93.5) | 10,657 (96.0) | 9,419 (96.4) | 963 (93.7) | 137 (93.2) | 7,258 (96.3) | 6,415 (96.5) | 643 (94.7) | 107 (95.5) | 349 (94.3) | 282 (93.7) | 26 (96.3) | 30 (96.8) |
| *Missing or never had a period* | *0 (0.0)* | *0 (0.0)* | *0 (0.0)* | *0 (0.0)* | *1 (0.0)* | *1 (0.0)* | *0 (0.0)* | *0 (0.0)* | *4 (0.0)* | *4 (0.0)* | *0 (0.0)* | *0 (0.0)* | *1 (0.0)* | *1 (0.0)* | *0 (0.0)* | *0 (0.0)* | *0 (0.0)* | *0 (0.0)* | *0 (0.0)* | *0 (0.0)* |
| Trouble sleeping for any other reason [j ǂ] | | | | | | | | | | | | | | | | | | | | |
| Yes | 3,876 (69.8) | 3,334 (67.6) | 430 (87.4) | 55 (85.9) | 6,290 (70.0) | 5,375 (67.5) | 702 (88.6) | 101 (93.5) | 7,916 (71.3) | 6,722 (68.8) | 921 (89.6) | 131 (89.1) | 5,522 (73.2) | 4,725 (71.0) | 605 (89.1) | 96 (85.7) | 249 (67.3) | 191 (63.5) | 24 (88.9) | 23 (74.2) |
| No | 1,675 (30.2) | 1,601 (32.4) | 62 (12.6) | 9 (14.1) | 2,695 (30.0) | 2,592 (32.5) | 90 (11.4) | 7 (6.5) | 3,181 (28.7) | 3,050 (31.2) | 107 (10.4) | 16 (10.9) | 2,019 (26.8) | 1,926 (29.0) | 74 (10.9) | 16 (14.3) | 121 (32.7) | 110 (36.5) | 3 (11.1) | 8 (25.8) |
| *Missing* | *0 (0.0)* | *0 (0.0)* | *0 (0.0)* | *0 (0.0)* | *0 (0.0)* | *0 (0.0)* | *0 (0.0)* | *0 (0.0)* | *1 (0.0)* | *1 (0.0)* | *0 (0.0)* | *0 (0.0)* | *0 (0.0)* | *0 (0.0)* | *0 (0.0)* | *0 (0.0)* | *0 (0.0)* | *0 (0.0)* | *0 (0.0)* | *0 (0.0)* |
| **Sleep health dimensions** | | | | | | | | | | | | | | | | | | | | |
| Average weekly sleep duration [k ǂ] | | | | | | | | | | | | | | | | | | | | |
| Short | 709 (12.8) | 624 (12.6) | 61 (12.4) | 17 (26.6) | 944 (10.5) | 792 (9.9) | 119 (15.0) | 17 (15.7) | 1,369 (12.3) | 1,172 (12.0) | 140 (13.6) | 26 (17.7) | 743 (9.9) | 648 (9.7) | 75 (11.1) | 12 (10.7) | 58 (15.7) | 46 (15.3) | 6 (22.2) | 4 (12.9) |
| Recommended | 4,005 (72.2) | 3,571 (72.4) | 357 (72.6) | 35 (54.7) | 6,657 (74.1) | 5,974 (75.0) | 542 (68.4) | 71 (65.7) | 7,912 (71.3) | 7,033 (72.0) | 703 (68.4) | 87 (59.2) | 5,590 (74.1) | 4,980 (74.9) | 472 (69.5) | 67 (59.8) | 231 (62.4) | 200 (66.5) | 8 (29.6) | 18 (58.1) |
| Long | 765 (13.8) | 673 (13.6) | 71 (14.4) | 10 (15.6) | 1,273 (14.2) | 1,100 (13.8) | 126 (15.9) | 16 (14.8) | 1,693 (15.3) | 1,463 (15.0) | 174 (16.9) | 29 (19.7) | 1,131 (15.0) | 957 (14.4) | 127 (18.7) | 30 (26.8) | 61 (16.5) | 41 (13.6) | 10 (37.0) | 6 (19.4) |
| *Missing* | *72 (1.3)* | *67 (1.4)* | *3 (0.6)* | *2 (3.1)* | *111 (1.2)* | *101 (1.3)* | *5 (0.6)* | *4 (3.7)* | *124 (1.1)* | *105 (1.1)* | *11 (1.1)* | *5 (3.4)* | *77 (1.0)* | *66 (1.0)* | *5 (0.7)* | *3 (2.7)* | *20 (5.4)* | *14 (4.7)* | *3 (11.1)* | *3 (9.7)* |
| Long sleep onset latency [l ǂ] | | | | | | | | | | | | | | | | | | | | |
| Yes | 634 (11.4) | 499 (10.1) | 102 (20.7) | 13 (20.3) | 1,064 (11.8) | 822 (10.3) | 161 (20.3) | 34 (31.5) | 1,501 (13.5) | 1,126 (11.5) | 278 (27) | 34 (23.1) | 910 (12.1) | 700 (10.5) | 141 (20.8) | 28 (25.0) | 74 (20.0) | 50 (16.6) | 7 (25.9) | 11 (35.5) |
| No | 4,846 (87.3) | 4,373 (88.6) | 384 (78.1) | 49 (76.6) | 7,832 (87.2) | 7,063 (88.7) | 627 (79.2) | 73 (67.6) | 9,457 (85.2) | 8,521 (87.2) | 741 (72.1) | 111 (75.5) | 6,545 (86.8) | 5,872 (88.3) | 534 (78.7) | 82 (73.2) | 272 (73.5) | 236 (78.4) | 16 (59.3) | 16 (51.6) |
| *Missing* | *71 (1.3)* | *63 (1.3)* | *6 (1.2)* | *2 (3.1)* | *89 (1.0)* | *82 (1.0)* | *4 (0.5)* | *1 (0.9)* | *140 (1.3)* | *126 (1.3)* | *9 (0.9)* | *2 (1.4)* | *86 (1.1)* | *79 (1.2)* | *4 (0.6)* | *2 (1.8)* | *24 (6.5)* | *15 (5.0)* | *4 (14.8)* | *4 (12.9)* |
| Poor sleep maintenance [m ǂ] | | | | | | | | | | | | | | | | | | | | |
| Yes | 2,228 (40.1) | 1,808 (36.6) | 339 (68.9) | 33 (51.6) | 3,437 (38.3) | 2,781 (34.9) | 501 (63.3) | 70 (64.8) | 4,336 (39.1) | 3,477 (35.6) | 661 (64.3) | 85 (57.8) | 3,141 (41.7) | 2,565 (38.6) | 447 (65.8) | 59 (52.7) | 118 (31.9) | 81 (26.9) | 16 (59.3) | 15 (48.4) |
| No | 3,259 (58.7) | 3,066 (62.1) | 151 (30.7) | 30 (46.9) | 5,454 (60.7) | 5,102 (64.0) | 284 (35.9) | 37 (34.3) | 6,619 (59.6) | 6,162 (63.1) | 359 (34.9) | 62 (42.2) | 4,315 (57.2) | 4,007 (60.3) | 228 (33.6) | 52 (46.4) | 230 (62.2) | 203 (67.4) | 10 (37.0) | 13 (41.9) |
| *Missing* | *64 (1.2)* | *61 (1.2)* | *2 (0.4)* | *1 (1.6)* | *94 (1.1)* | *84 (1.1)* | *7 (0.9)* | *1 (0.9)* | *143 (1.3)* | *134 (1.4)* | *8 (0.8)* | *0 (0.0)* | *85 (1.1)* | *79 (1.2)* | *4 (0.6)* | *1 (0.9)* | *22 (6.0)* | *17 (5.7)* | *1 (3.7)* | *3 (9.7)* |
| Insomnia symptoms [n ǂ] | | | | | | | | | | | | | | | | | | | | |
| Yes | 2,380 (42.9) | 1,939 (39.3) | 356 (72.4) | 34 (53.1) | 3,739 (41.6) | 3,035 (38.1) | 536 (67.7) | 74 (68.5) | 4,742 (42.7) | 3,814 (39.0) | 712 (69.3) | 92 (62.6) | 3,357 (44.5) | 2,737 (41.2) | 480 (70.7) | 62 (55.4) | 137 (37.0) | 96 (31.9) | 18 (66.7) | 17 (54.8) |
| No | 3,133 (56.4) | 2,960 (60.0) | 135 (27.4) | 29 (45.3) | 5,195 (57.8) | 4,884 (61.3) | 254 (32.1) | 34 (31.5) | 6,275 (56.5) | 5,883 (60.2) | 312 (30.4) | 55 (37.4) | 4,136 (54.9) | 3,868 (58.2) | 198 (29.2) | 49 (43.8) | 215 (58.1) | 192 (63.8) | 8 (29.6) | 11 (35.5) |
| *Missing* | *38 (0.7)* | *36 (0.7)* | *1 (0.2)* | *1 (1.6)* | *51 (0.6)* | *48 (0.6)* | *2 (0.3)* | *0 (0.0)* | *81 (0.7)* | *76 (0.8)* | *4 (0.4)* | *0 (0.0)* | *48 (0.6)* | *46 (0.7)* | *1 (0.2)* | *1 (0.9)* | *18 (4.9)* | *13 (4.3)* | *1 (3.7)* | *3 (9.7)* |
| Sleep medication use [o ǂ] | | | | | | | | | | | | | | | | | | | | |
| Yes | 914 (16.5) | 782 (15.9) | 101 (20.5) | 19 (29.7) | 1,617 (18.0) | 1,392 (17.5) | 160 (20.2) | 31 (28.7) | 2,163 (19.5) | 1,839 (18.8) | 242 (23.5) | 35 (23.8) | 1,519 (20.1) | 1,317 (19.8) | 151 (22.2) | 24 (21.4) | 77 (20.8) | 54 (17.9) | 7 (25.9) | 12 (38.7) |
| No | 4,611 (83.1) | 4,132 (83.7) | 387 (78.7) | 45 (70.3) | 7,320 (81.5) | 6,531 (82.0) | 628 (79.3) | 77 (71.3) | 8,888 (80.1) | 7,895 (80.8) | 781 (76.0) | 110 (74.8) | 5,991 (79.5) | 5,306 (79.8) | 527 (77.6) | 87 (77.7) | 283 (76.5) | 240 (79.7) | 20 (74.1) | 18 (58.1) |
| Missing | 26 (0.5) | 21 (0.4) | 4 (0.8) | 0 (0.0) | 48 (0.5) | 44 (0.6) | 4 (0.5) | 0 (0.0) | 47 (0.4) | 39 (0.4) | 5 (0.5) | 2 (1.4) | 31 (0.4) | 28 (0.4) | 1 (0.2) | 1 (0.9) | 10 (2.7) | 7 (2.3) | 0 (0.0) | 1 (3.2) |
| Daytime dysfunction [p] | | | | | | | | | | | | | | | | | | | | |

| | | | | | | | | | | | | | | | | | | | | |
|---|---|---|---|---|---|---|---|---|---|---|---|---|---|---|---|---|---|---|---|---|
| Yes | 36 (0.7) | 29 (0.6) | 5 (1.0) | 2 (3.1) | 63 (0.7) | 53 (0.7) | 7 (0.9) | 1 (0.9) | 101 (0.9) | 68 (0.7) | 17 (1.7) | 8 (5.4) | 50 (0.7) | 36 (0.5) | 8 (1.2) | 4 (3.6) | 2 (0.5) | 0 (0.0) | 1 (3.7) | 1 (3.2) |
| No | 5,452 (98.2) | 4,847 (98.2) | 483 (98.2) | 62 (96.9) | 8,811 (98.1) | 7,816 (98.1) | 774 (97.7) | 106 (98.2) | 10,867 (97.9) | 9,589 (98.1) | 999 (97.2) | 138 (93.9) | 7,400 (98.1) | 6,531 (98.2) | 668 (98.4) | 106 (94.6) | 356 (96.2) | 291 (96.7) | 25 (92.6) | 30 (96.8) |
| *Missing* | *63 (1.1)* | *59 (1.2)* | *4 (0.8)* | *0 (0.0)* | *111 (1.2)* | *98 (1.2)* | *11 (1.4)* | *1 (0.9)* | *130 (1.2)* | *116 (1.2)* | *12 (1.2)* | *1 (0.7)* | *91 (1.2)* | *84 (1.3)* | *3 (0.4)* | *2 (1.8)* | *12 (3.2)* | *10 (3.3)* | *1 (3.7)* | *0 (0.0)* |
| Healthcare provider diagnosed sleep apnea [q ł] | | | | | | | | | | | | | | | | | | | | |
| Yes | 470 (8.5) | 403 (8.2) | 54 (11.0) | 9 (14.1) | 936 (10.4) | 820 (10.3) | 89 (11.2) | 10 (9.3) | 1,177 (10.6) | 981 (10.0) | 144 (14.0) | 31 (21.1) | 852 (11.3) | 741 (11.1) | 78 (11.5) | 20 (17.9) | 18 (4.9) | 9 (3.0) | 3 (11.1) | 6 (19.4) |
| No | 5,024 (90.5) | 4,481 (90.8) | 433 (88.0) | 54 (84.4) | 7,944 (88.4) | 7,058 (88.6) | 696 (87.9) | 92 (85.2) | 9,804 (88.3) | 8,684 (88.9) | 880 (85.6) | 112 (76.2) | 6,615 (87.7) | 5,845 (87.9) | 595 (87.6) | 91 (81.3) | 344 (93.0) | 287 (95.4) | 24 (88.9) | 24 (77.4) |
| *Missing* | *57 (1.0)* | *51 (1.0)* | *5 (1.0)* | *1 (1.6)* | *105 (1.2)* | *89 (1.1)* | *7 (0.9)* | *6 (5.6)* | *117 (1.1)* | *108 (1.1)* | *4 (0.4)* | *4 (2.7)* | *74 (1.0)* | *65 (1.0)* | *6 (0.9)* | *1 (0.9)* | *8 (2.2)* | *5 (1.7)* | *0 (0.0)* | *1 (3.2)* |

Data presented as mean ± standard deviation or n (%)
Note: N=438 (1.3%) participants who reported both 'too hot' and 'too cold' temperatures were not included in the analysis.
Abbreviations: SD, Standard Deviation; GED, General Educational Development
[¥]Chi-square test p-values indicate significant differences between in the overall prevalence of 'too hot' by region of residence.
[§]Chi-square test p-values indicate significant differences between the overall prevalence of 'too cold' by region or residence.
[ł]Chi-square or ANOVA test p-values indicate significant differences in characteristics between the total populations of each region of residence.

[a] Excluded for reporting both 'Too hot' and 'Too cold', Northeast: n = 60 (1.1%)
[b] Excluded for reporting both 'Too hot' and 'Too cold', Midwest: n = 118 (1.3%)
[c] Excluded for reporting both 'Too hot' and 'Too cold', South: n = 150 (1.4%)
[d] Excluded for reporting both 'Too hot' and 'Too cold', West: n = 99 (1.3%)
[e] Excluded for reporting both 'Too hot' and 'Too cold', Puerto Rico: n = 11 (3.0%)
[f] 'Infrequent temperature extremes' is defined as self-reported trouble sleeping due to feeling too hot < 3 times per week and trouble sleeping due to feeling too cold < 3 times per week in the past month.
[g] 'Too hot' is defined as self-reported trouble sleeping due to feeling too hot ≥ 3 times per week in the past month.
[h] 'Too cold' is defined as self-reported trouble sleeping due to feeling too cold ≥ 3 times per week in the past month.
[i] Educational attainment was assessed at baseline.
[j] Reasons for trouble sleeping ≥ 3 times per week in the past month include: unable to fall asleep within 30 minutes, waking up in the middle of the night or early morning, waking up to use the bathroom, cannot breathe comfortably, coughing or snoring loudly, having bad dreams, having pain, or other non-specified reasons.
[k] Sleep duration is based on reported bed and wake times or reported average sleep duration. Participants who reported ≤ 2 hours or ≥ 23 hours of sleep are excluded. Short: <7 hours; Recommended: 7-9 hours; Long >9 hours
[l] Long sleep onset latency is defined as not falling asleep within 30 minutes at least three times a week during the past month.
[m] Poor sleep maintenance is defined as waking up in the middle of the night or early morning at least three times a week during the past month.
[n] Insomnia symptoms is defined as having long sleep onset latency or poor sleep maintenance.
[o] Sleep medication use is defined as 'taking medicine (prescription or over the counter) to help you sleep' at least three times a week during the past month.
[p] Daytime dysfunction is defined as having trouble staying awake while driving, eating, or engaging in social activity at least three times a week during the past month.
[q] Healthcare provider diagnosed sleep apnea is defined as a current doctor or other health professional diagnosis of sleep apnea.

**Supplemental Table 5. Sociodemographic characteristics, clinical characteristics, and sleep health dimensions among eligible Sister Study participants by menopausal status at 4th follow-up as well as extreme indoor temperature, Sister Study, 2017 – 2019, N = 33,545**

| Menopausal status | Premenopausal [a] | | | | Postmenopausal [b] | | | |
|---|---|---|---|---|---|---|---|---|
| | Total n = 1,346 (4.0%) | Infrequent temperature extremes [c] n = 1,110 (82.5%) | Too hot [d ¥] n = 183 (13.6%) | Too cold [e §] n = 25 (1.9%) | Total n = 32,193 (96.0%) | Infrequent temperature extremes [c] n = 28,511 (88.6%) | Too hot [d ¥] n = 2,835 (8.8%) | Too cold [e §] n = 437 (1.4%) |
| **Sociodemographic characteristics** | | | | | | | | |
| Age, years (mean ± SD) [ł] | 51.2 ± 2.6 | 51.1 ± 2.6 | 51.7 ± 2.6 | 50.9 ± 2.7 | 67.8 ± 8.0 | 68.3 ± 8.0 | 63.3 ± 7.3 | 69.5 ± 8.3 |
| Age [ł] | | | | | | | | |
| < 67.2 yrs | 1,346 (100.0) | 1,110 (100.0) | 183 (100.0) | 25 (100.0) | 15,432 (47.9) | 12,977 (45.5) | 1,997 (70.4) | 166 (38.0) |
| ≥ 67.2 yrs | 0 (0.0) | 0 (0.0) | 0 (0.0) | 0 (0.0) | 16,761 (52.1) | 15,534 (54.5) | 838 (29.6) | 271 (62.0) |
| Race and ethnicity [ł] | | | | | | | | |
| Hispanic/Latina | 87 (6.5) | 79 (7.1) | 7 (3.8) | 1 (4.0) | 1,138 (3.5) | 963 (3.4) | 105 (3.7) | 45 (10.3) |
| Non-Hispanic Black | 143 (10.6) | 104 (9.4) | 29 (15.9) | 5 (20.0) | 2,055 (6.4) | 1,696 (6.0) | 295 (10.4) | 29 (6.6) |
| Non-Hispanic White | 1,116 (82.9) | 927 (83.5) | 147 (80.3) | 19 (76.0) | 29,000 (90.1) | 25,852 (90.7) | 2,435 (85.9) | 363 (83.1) |
| Educational attainment [f ł] | | | | | | | | |
| <High school / GED | 112 (8.3) | 87 (7.8) | 18 (9.8) | 5 (20.0) | 4,349 (13.5) | 3,792 (13.3) | 410 (14.5) | 88 (20.1) |
| Some college / technical | 352 (26.2) | 276 (24.9) | 55 (30.1) | 8 (32.0) | 10,236 (31.8) | 8,968 (31.5) | 963 (34.0) | 160 (36.6) |
| ≥ Bachelor's or higher | 882 (65.5) | 747 (67.3) | 110 (60.1) | 12 (48.0) | 17,604 (54.7) | 15,748 (55.2) | 1,462 (51.6) | 189 (43.3) |
| *Missing* | *0 (0.0)* | *0 (0.0)* | *0 (0.0)* | *0 (0.0)* | *4 (0.0)* | *3 (0.0)* | *0 (0.0)* | *0 (0.0)* |
| Annual household income [ł] | | | | | | | | |
| < $20,000 - $49,999 | 144 (10.7) | 116 (10.5) | 22 (12.0) | 1 (4.0) | 8,751 (27.2) | 7,828 (27.5) | 617 (21.8) | 195 (44.6) |
| $50,000 – $99,999 | 345 (25.6) | 272 (24.5) | 53 (29.0) | 9 (36.0) | 12,031 (37.4) | 10,737 (37.7) | 1,018 (35.9) | 125 (28.6) |
| ≥ $100,000 | 857 (63.7) | 722 (65.1) | 108 (59.0) | 15 (60.0) | 11,411 (35.5) | 9,946 (34.9) | 1,200 (42.3) | 117 (26.8) |
| Marital status [ł] | | | | | | | | |
| Married or living as though married | 1,025(76.2) | 852 (76.8) | 137 (74.9) | 21 (84.0) | 21,775 (67.6) | 19,014 (66.7) | 2,188(77.2) | 275 (62.9) |
| Never married, divorced, widowed, or separated | 321 (23.8) | 258 (23.2) | 46 (25.1) | 4 (16.0) | 10,418 (32.4) | 9,497 (33.3) | 647 (22.8) | 162 (37.1) |
| Region of residence | | | | | | | | |
| Northeast | 230 (17.1) | 192 (17.3) | 31 (16.9) | 2 (8.0) | 5,321 (16.5) | 4,743 (16.6) | 461 (16.3) | 62 (14.2) |
| Midwest | 376 (27.9) | 314 (28.3) | 50 (27.3) | 7 (28.0) | 8,608 (26.7) | 7,652 (26.8) | 742 (26.2) | 101 (23.1) |
| South | 437 (32.5) | 350 (31.5) | 65 (35.5) | 10 (40.0) | 10,657 (33.1) | 9,419 (33.0) | 963 (34.0) | 137 (31.4) |
| West | 282 (21.0) | 235 (21.2) | 36 (19.7) | 5 (20.0) | 7,258 (22.6) | 6,415 (22.5) | 643 (22.7) | 107 (24.5) |
| Puerto Rico | 21 (1.6) | 19 (1.7) | 1 (0.6) | 1 (4.0) | 349 (1.1) | 282 (1.0) | 26 (0.9) | 30 (6.9) |
| **Clinical characteristics** | | | | | | | | |
| Body Mass Index, $kg/m^2$ [ł] | | | | | | | | |
| 18.5 - 24.9 | 496 (36.9) | 426 (38.4) | 47 (25.7) | 10 (40.0) | 12,630 (39.2) | 11,358 (39.8) | 941 (33.2) | 180 (41.2) |
| 25.0 - 29.9 | 390 (29.0) | 320 (28.8) | 57 (31.2) | 5 (20.0) | 10,375 (32.2) | 9,172 (32.2) | 956 (33.7) | 118 (27.0) |
| ≥30 | 446 (33.1) | 354 (31.9) | 75 (41.0) | 10 (40.0) | 8,772 (27.3) | 7,612 (26.7) | 906 (32.0) | 133 (30.4) |
| *Missing* | *14 (1.0)* | *10 (0.9)* | *4 (2.2)* | *0 (0.0)* | *416 (1.3)* | *369 (1.3)* | *32 (1.1)* | *6 (1.4)* |

| | | | | | | | | |
|---|---|---|---|---|---|---|---|---|
| Trouble sleeping for any other reason [g ƚ] | | | | | | | | |
| Yes | 791 (58.8) | 588 (53) | 158 (86.3) | 20 (80.0) | 23,057 (71.6) | 19,754 (69.3) | 2,524 (89.0) | 386 (88.3) |
| No | 555 (41.2) | 522 (47) | 25 (13.7) | 5 (20.0) | 9,135 (28.4) | 8,756 (30.7) | 311 (11.0) | 51 (11.7) |
| *Missing* | *0 (0.0)* | *0 (0.0)* | *0 (0.0)* | *0 (0.0)* | *1 (0.0)* | *1 (0.0)* | *0 (0.0)* | *0 (0.0)* |
| **Sleep health characteristics** | | | | | | | | |
| Average weekly sleep duration [h ƚ] | | | | | | | | |
| Short | 183 (13.6) | 146 (13.2) | 27 (14.8) | 6 (24.0) | 3,640 (11.3) | 3,136 (11.0) | 374 (13.2) | 70 (16.0) |
| Recommended | 1,023 (76) | 858 (77.3) | 130 (71) | 16 (64.0) | 23,367 (72.6) | 20,895 (73.3) | 1,952 (68.9) | 262 (60.0) |
| Long | 134 (10.0) | 102 (9.2) | 25 (13.7) | 3 (12.0) | 4,788 (14.9) | 4,131 (14.5) | 483 (17.0) | 88 (20.1) |
| *Missing* | *6 (0.5)* | *4 (0.4)* | *1 (0.6)* | *0 (0.0)* | *398 (1.2)* | *349 (1.2)* | *26 (0.9)* | *17 (3.9)* |
| Long sleep onset latency [i ƚ] | | | | | | | | |
| Yes | 125 (9.3) | 76 (6.9) | 36 (19.7) | 2 (8.0) | 4,057 (12.6) | 3,120 (10.9) | 653 (23.0) | 118 (27.0) |
| No | 1,207 (89.7) | 1,024 (92.3) | 144 (78.7) | 23 (92.0) | 27,740 (86.2) | 25,036 (87.8) | 2,158 (76.1) | 308 (70.5) |
| *Missing* | *14 (1.0)* | *10 (0.9)* | *3 (1.6)* | *0 (0.0)* | *396 (1.2)* | *355 (1.3)* | *24 (0.9)* | *11 (2.5)* |
| Poor sleep maintenance [j ƚ] | | | | | | | | |
| Yes | 478 (35.5) | 330 (29.7) | 117 (63.9) | 14 (56.0) | 12,779 (39.7) | 10,379 (36.4) | 1,847 (65.2) | 248 (56.8) |
| No | 856 (63.6) | 773 (69.6) | 61 (33.3) | 11 (44.0) | 19,018 (59.1) | 17,764 (62.3) | 971 (34.3) | 183 (41.9) |
| *Missing* | *12 (0.9)* | *7 (0.6)* | *5 (2.7)* | *0 (0.0)* | *396 (1.2)* | *368 (1.3)* | *17 (0.6)* | *6 (1.4)* |
| Insomnia symptoms [k ƚ] | | | | | | | | |
| Yes | 512 (38.0) | 354 (31.9) | 125 (68.3) | 14 (56.0) | 13,840 (43.0) | 11,264 (39.5) | 1,977 (69.7) | 265 (60.6) |
| No | 827 (61.4) | 750 (67.6) | 57 (31.2) | 11 (44.0) | 18,124 (56.3) | 17,034 (59.8) | 850 (30.0) | 167 (38.2) |
| *Missing* | *7 (0.5)* | *6 (0.5)* | *1 (0.6)* | *0 (0.0)* | *229 (0.7)* | *213 (0.8)* | *8 (0.3)* | *5 (1.1)* |
| Sleep medication use [l ƚ] | | | | | | | | |
| Yes | 181 (13.5) | 139 (12.5) | 30 (16.4) | 6 (24.0) | 6,109 (19.0) | 5,245 (18.4) | 631 (22.3) | 115 (26.3) |
| No | 1,159 (86.1) | 965 (86.9) | 153 (83.6) | 19 (76.0) | 25,928 (80.5) | 23,133 (81.1) | 2,190 (77.3) | 318 (72.8) |
| *Missing* | *6 (0.5)* | *6 (0.5)* | *0 (0.0)* | *0 (0.0)* | *156 (0.5)* | *133 (0.5)* | *14 (0.5)* | *4 (0.9)* |
| Daytime dysfunction [m] | | | | | | | | |
| Yes | 8 (0.6) | 5 (0.5) | 1 (0.6) | 2 (8.0) | 243 (0.8) | 180 (0.6) | 37 (1.3) | 14 (3.2) |
| No | 1,326 (98.5) | 1,095 (98.7) | 180 (98.4) | 23 (92.0) | 31,555 (98.0) | 27,974 (98.1) | 2,769 (97.7) | 419 (95.9) |
| *Missing* | *12 (0.9)* | *10 (0.9)* | *2 (1.1)* | *0 (0.0)* | *395 (1.2)* | *357 (1.3)* | *29 (1.0)* | *4 (0.9)* |
| Healthcare provider diagnosed sleep apnea [n ƚ] | | | | | | | | |
| Yes | 93 (6.9) | 73 (6.6) | 14 (7.7) | 2 (8.0) | 3,359 (10.4) | 2,880 (10.1) | 354 (12.5) | 74 (16.9) |
| No | 1,247 (92.6) | 1,031 (92.9) | 169 (92.4) | 23 (92.0) | 28,479 (88.5) | 25,319 (88.8) | 2,459 (86.7) | 350 (80.1) |
| *Missing* | *6 (0.5)* | *6 (0.5)* | *0 (0.0)* | *0 (0.0)* | *355 (1.1)* | *312 (1.1)* | *22 (0.8)* | *13 (3.0)* |

Data presented as mean ± standard deviation or n (%)

Note: N=438 (1.3%) participants who reported both ‘too hot’ and ‘too cold’ temperatures were not included in the analysis.

Abbreviations: SD, Standard Deviation; GED, General Educational Development

[¥]Chi-square test p-values indicate significant differences between the overall prevalence of ‘too hot’ by menopausal status.

[§]Chi-square test p-values indicate significant differences between the overall prevalence of ‘too cold’ by menopausal status.

[ƚ]Chi-square or ANOVA test p-values indicate significant differences in characteristics between the total populations of each menopausal status category.

[a] Excluded for reporting both ‘Too hot’ and ‘Too cold’, Premenopausal: n = 28 (2.1%)
[b] Excluded for reporting both ‘Too hot’ and ‘Too cold’, Postmenopausal: n = 410 (1.3%)
[c] ‘Infrequent temperature extremes’ is defined as self-reported trouble sleeping due to feeling too hot < 3 times per week and trouble sleeping due to feeling too cold < 3 times per week in the past month.
[d] ’Too hot’ is defined as self-reported trouble sleeping due to feeling too hot ≥ 3 times per week in the past month.
[e] ’Too cold’ is defined as self-reported trouble sleeping due to feeling too cold ≥ 3 times per week in the past month.
[f] Educational attainment was assessed at baseline.
[g] Reasons for trouble sleeping ≥ 3 times per week in the past month include: unable to fall asleep within 30 minutes, waking up in the middle of the night or early morning, waking up to use the bathroom, cannot breathe comfortably, coughing or snoring loudly, having bad dreams, having pain, or other non-specified reasons.
[h] Sleep duration is based on reported bed and wake times or reported average sleep duration. Participants who reported ≤ 2 hours or ≥ 23 hours of sleep are excluded. Short: <7 hours; Recommended: 7-9 hours; Long >9 hours
[i] Long sleep onset latency is defined as not falling asleep within 30 minutes at least three times a week during the past month.
[j] Poor sleep maintenance is defined as waking up in the middle of the night or early morning at least three times a week during the past month.
[k] Insomnia symptoms is defined as long sleep onset latency or poor sleep maintenance.
[l] Sleep medication use is defined as 'taking medicine (prescription or over the counter) to help you sleep' at least three times a week during the past month.
[m] Daytime dysfunction is defined as having trouble staying awake while driving, eating, or engaging in social activity at least three times a week during the past month.
[n] Healthcare provider diagnosed sleep apnea is defined as a current doctor or other health professional diagnosis of sleep apnea

**Supplemental Table 6. Prevalence ratios (95% confidence intervals) for associations between perceived indoor temperature extremes and sleep health dimensions, Sister Study, 2017-2019, N=33,545**

| | **Indoor temperature: Too hot [a]** | | | | **Indoor temperature: Too cold [b]** | | | |
|---|---|---|---|---|---|---|---|---|
| | Crude | Model 1 [c] | Model 2 | Model 3 | Crude | Model 1 [c] | Model 2 | Model 3 |
| Weekly sleep duration [d] | | | | | | | | |
| Short | **1.23 (1.12,1.36)ǂ** | **1.19 (1.08,1.31)‡** | 1.09 (0.99,1.21) | 1.09 (0.99,1.20) | **1.66 (1.35,2.03)‡** | **1.60 (1.31,1.96)‡** | **1.46 (1.20,1.78)‡** | **1.46 (1.20,1.78)‡** |
| Recommended | 1.00 (Ref) | 1.00 (Ref) | 1.00 (Ref) | 1.00 (Ref) | 1.00 (Ref) | 1.00 (Ref) | 1.00 (Ref) | 1.00 (Ref) |
| Long | **1.20 (1.10,1.30)‡** | **1.13 (1.03,1.22)‡** | **1.26 (1.15,1.37)‡** | **1.25 (1.15,1.36)‡** | **1.51 (1.25,1.81)‡** | **1.42 (1.18,1.70)‡** | **1.35 (1.12,1.62)‡** | **1.35 (1.13,1.62)‡** |
| Long sleep onset latency [e] | | | | | | | | |
| Yes | **2.11 (1.96,2.27)‡** | **1.89 (1.76,2.04)‡** | **1.91 (1.77,2.06)‡** | **1.88 (1.74,2.03)‡** | **2.44 (2.09,2.86)‡** | **2.14 (1.82,2.50)‡** | **1.99 (1.70,2.33)‡** | **1.99 (1.70,2.33)‡** |
| No | 1.00 (Ref) | 1.00 (Ref) | 1.00 (Ref) | 1.00 (Ref) | 1.00 (Ref) | 1.00 (Ref) | 1.00 (Ref) | 1.00 (Ref) |
| Poor sleep maintenance [f] | | | | | | | | |
| Yes | **1.79 (1.74,1.85)‡** | **1.58 (1.53,1.63)‡** | **1.57 (1.52,1.62)‡** | **1.57 (1.52,1.62)‡** | **1.56 (1.44,1.69)‡** | **1.33 (1.22,1.44)‡** | **1.34 (1.24,1.45)‡** | **1.34 (1.23,1.45)‡** |
| No | 1.00 (Ref) | 1.00 (Ref) | 1.00 (Ref) | 1.00 (Ref) | 1.00 (Ref) | 1.00 (Ref) | 1.00 (Ref) | 1.00 (Ref) |
| Insomnia symptoms [g] | | | | | | | | |
| Yes | **1.77 (1.72,1.82)‡** | **1.57 (1.53,1.62)‡** | **1.57 (1.52,1.61)‡** | **1.56 (1.52,1.61)‡** | **1.54 (1.43,1.66)‡** | **1.32 (1.23,1.43)‡** | **1.33 (1.24,1.44)‡** | **1.33 (1.23,1.43)‡** |
| No | 1.00 (Ref) | 1.00 (Ref) | 1.00 (Ref) | 1.00 (Ref) | 1.00 (Ref) | 1.00 (Ref) | 1.00 (Ref) | 1.00 (Ref) |
| Sleep medication use [h] | | | | | | | | |
| Yes | **1.20 (1.12,1.29)‡** | **1.10 (1.02,1.18)‡** | **1.18 (1.10,1.27)‡** | **1.18 (1.09,1.27)‡** | **1.44 (1.23,1.68)‡** | **1.33 (1.13,1.55)‡** | **1.31 (1.12,1.53)‡** | **1.31 (1.12,1.53)‡** |
| No | 1.00 (Ref) | 1.00 (Ref) | 1.00 (Ref) | 1.00 (Ref) | 1.00 (Ref) | 1.00 (Ref) | 1.00 (Ref) | 1.00 (Ref) |
| Daytime dysfunction [i] | | | | | | | | |
| Yes | **2.03 (1.44,2.88)‡** | **1.79 (1.26,2.54)‡** | **1.79 (1.25,2.55)‡** | **1.76 (1.23,2.52)‡** | **5.59 (3.38,9.23)‡** | **4.94 (2.99,8.16)‡** | **4.81 (2.91,7.95)‡** | **4.69 (2.83,7.77)‡** |
| No | 1.00 (Ref) | 1.00 (Ref) | 1.00 (Ref) | 1.00 (Ref) | 1.00 (Ref) | 1.00 (Ref) | 1.00 (Ref) | 1.00 (Ref) |
| Healthcare professional diagnosed sleep apnea [j] | | | | | | | | |
| Yes | **1.22 (1.10,1.35)‡** | **1.19 (1.07,1.32)‡** | **1.26 (1.14,1.40)‡** | **1.20 (1.09,1.33)‡** | **1.67 (1.36,2.06)‡** | **1.63 (1.32,2.01)‡** | **1.64 (1.33,2.01)‡** | **1.62 (1.33,1.97)‡** |
| No | 1.00 (Ref) | 1.00 (Ref) | 1.00 (Ref) | 1.00 (Ref) | 1.00 (Ref) | 1.00 (Ref) | 1.00 (Ref) | 1.00 (Ref) |

Note: N=438 (1.3%) participants who reported both ‘too hot’ and ‘too cold’ temperatures were not included in the analysis.

Model 1: Adjusted for frequent trouble sleeping for any reason other than feeling too hot or feeling too cold.
Model 2: Adjusted for frequent trouble sleeping for any reason other than feeling too hot or feeling too cold, age (continuous), age$^2$, race and ethnicity (Hispanic/Latina, non-Hispanic Black, non-Hispanic White), annual household income (<$20,000 - $49,999, $50,000 - $99,999, ≥$100,000), marital status (married or living as though married, divorced/widowed/separated/never married), and region of residence (Northeast, Midwest, South, West, and Puerto Rico).
Model 3: Adjusted for trouble sleeping for any reason other than feeling too hot or feeling too cold, age (continuous), age$^2$, race and ethnicity (Hispanic/Latina, non-Hispanic Black, non-Hispanic White), annual household income (<$20,000 - $49,999, $50,000 - $99,999, ≥$100,000), marital status (married or living as though married, divorced/widowed/separated/never married), region of residence (Northeast, Midwest, South, West, and Puerto Rico), body mass index (BMI: underweight, recommended, overweight, obesity), and menopausal status (premenopausal, postmenopausal).

[a] Too hot’ is defined as self-reported trouble sleeping due to feeling too hot ≥ 3 times per week vs. trouble sleeping due to feeling too hot < 3 times per week and trouble sleeping due to feeling too cold < 3 times per week.
[b] ’Too cold’ is defined as self-reported trouble sleeping due to feeling too cold ≥ 3 times per week vs. trouble sleeping due to feeling too hot < 3 times per week and trouble sleeping due to feeling too cold < 3 times per week.

[c] In models for weekly sleep duration, sleep medication use, daytime dysfunction, and healthcare professional diagnosed sleep apnea, reasons for trouble sleeping for any reason other than feeling too hot or feeling too cold ≥ 3 times per week in the past month include: unable to fall asleep within 30 minutes, waking up in the middle of the night or early morning, waking up to use the bathroom, cannot breathe comfortably, coughing or snoring loudly, having bad dreams, having pain, or other non-specified reasons. In models for long sleep onset latency, poor sleep maintenance, and insomnia symptoms, reasons for trouble sleeping for any reason other than feeling too hot or feeling too cold ≥ 3 times per week in the past month include waking up to use the bathroom, cannot breathe comfortably, coughing or snoring loudly, having bad dreams, having pain, or other non-specified reasons.
[d] Sleep duration is based on reported bed and wake times or reported average sleep duration. Participants who reported ≤ 2 hours or ≥ 23 hours of sleep are excluded. Short: <7 hours; Recommended: 7-9 hours; Long >9 hours
[e] Long sleep onset latency is defined as not falling asleep within 30 minutes at least three times a week during the past month.
[f] Poor sleep maintenance is defined as waking up in the middle of the night or early morning at least three times a week during the past month.
[g] Insomnia symptoms is defined as long sleep onset latency or poor sleep maintenance.
[h] Sleep medication use is defined as 'taking medicine (prescription or over the counter) to help you sleep' at least three times a week during the past month.
[i] Daytime dysfunction is defined as having trouble staying awake while driving, eating, or engaging in social activity at least three times a week during the past month.
[j] Healthcare provider diagnosed sleep apnea is defined as a current doctor or other health professional diagnosis of sleep apnea.
[ƚ] Remained significant after false discovery rate correction. False discovery rate-corrected P-value was considered statistically significant at the 0.05 level.

**Supplemental Table 7A. Prevalence ratios (95% confidence intervals) for associations between perceived indoor temperature extremes and sleep health dimensions by race and ethnicity, Sister Study, 2017-2019, N=33,545**

| | **Indoor temperature: Too hot [a]** | | | | **Indoor temperature: Too cold [b]** | | | |
|---|---|---|---|---|---|---|---|---|
| | Hispanic/Latina | Non-Hispanic Black | Non-Hispanic White | Wald p-values | Hispanic/Latina | Non-Hispanic Black | Non-Hispanic White | Wald p-values |
| Weekly sleep duration [c] | | | | | | | | |
| Short | 1.19 (0.79,1.80) | 0.97 (0.81,1.17) | **1.12 (1.00,1.26)**[‡] | 0.3834 | 0.92 (0.43,1.98) | 1.38 (0.96,2.00) | **1.57 (1.25,1.99)**[‡] | 0.3929 |
| Recommended | 1.00 (ref) | 1.00 (ref) | 1.00 (ref) | --- | 1.00 (ref) | 1.00 (ref) | 1.00 (ref) | --- |
| Long | **1.47 (1.02,2.13)**[‡] | 0.95 (0.69,1.32) | **1.27 (1.16,1.39)**[‡] | 0.1690 | 1.15 (0.61,2.14) | 1.27 (0.53,3.02) | **1.38 (1.14,1.68)**[‡] | 0.8460 |
| Long sleep onset latency [d] | | | | | | | | |
| Yes | **2.24 (1.70,2.96)**[‡] | **2.04 (1.65,2.52)**[‡] | **1.84 (1.69,2.00)**[‡] | 0.3079 | **1.70 (1.08,2.67)**[‡] | 1.52 (0.77,2.99) | **2.08 (1.75,2.47)**[‡] | 0.5131 |
| No | 1.00 (ref) | 1.00 (ref) | 1.00 (ref) | --- | 1.00 (ref) | 1.00 (ref) | 1.00 (ref) | --- |
| Poor sleep maintenance [e] | | | | | | | | |
| Yes | **1.68 (1.41,1.99)**[‡] | **1.47 (1.31,1.65)**[‡] | **1.58 (1.53,1.63)**[‡] | 0.4053 | **1.58 (1.18,2.11)**[‡] | **1.39 (1.05,1.85)**[‡] | **1.31 (1.20,1.43)**[‡] | 0.4680 |
| No | 1.00 (ref) | 1.00 (ref) | 1.00 (ref) | --- | 1.00 (ref) | 1.00 (ref) | 1.00 (ref) | --- |
| Insomnia symptoms [f] | | | | | | | | |
| Yes | **1.75 (1.51,2.02)**[‡] | **1.49 (1.34,1.65)**[‡] | **1.57 (1.52,1.61)**[‡] | 0.2065 | **1.50 (1.16,1.95)**[‡] | **1.45 (1.12,1.88)**[‡] | **1.30 (1.20,1.42)**[‡] | 0.4825 |
| No | 1.00 (ref) | 1.00 (ref) | 1.00 (ref) | --- | 1.00 (ref) | 1.00 (ref) | 1.00 (ref) | --- |
| Sleep medication use [g] | | | | | | | | |
| Yes | 1.13 (0.72,1.76) | 1.27 (0.93,1.72) | **1.17 (1.09,1.27)**[‡] | 0.8807 | 1.53 (0.94,2.48) | 1.08 (0.43,2.73) | **1.30 (1.10,1.54)**[‡] | 0.7548 |
| No | 1.00 (ref) | 1.00 (ref) | 1.00 (ref) | --- | 1.00 (ref) | 1.00 (ref) | 1.00 (ref) | --- |
| Daytime dysfunction [h] | | | | | | | | |
| Yes | 0.76 (0.09,6.37) | 2.32 (0.79,6.82) | **1.77 (1.21,2.60)**[‡] | 0.6548 | **7.15 (1.79,28.56)**[‡] | **12.52 (3.51,44.59)**[‡] | **3.84 (2.09,7.08)**[‡] | 0.2220 |
| No | 1.00 (ref) | 1.00 (ref) | 1.00 (ref) | --- | 1.00 (ref) | 1.00 (ref) | 1.00 (ref) | --- |
| Healthcare professional diagnosed sleep apnea [i] | | | | | | | | |
| Yes | 1.52 (0.90,2.58) | 1.29 (0.99,1.68) | **1.18 (1.05,1.32)**[‡] | 0.5567 | **3.12 (1.68,5.79)**[‡] | **2.39 (1.40,4.08)**[‡] | **1.45 (1.15,1.82)**[‡] | **0.0276** |
| No | 1.00 (ref) | 1.00 (ref) | 1.00 (ref) | --- | 1.00 (ref) | 1.00 (ref) | 1.00 (ref) | --- |

Note: N=438 (1.3%) participants who reported both ‘too hot’ and ‘too cold’ temperatures were not included in the analysis.

Models are adjusted for frequent trouble sleeping for any reason other than feeling too hot or feeling too cold, age (continuous), $age^2$, annual household income (<$20,000 - $49,999, $50,000 - $99,999, ≥$100,000), marital status (married or living as though married, divorced/widowed/separated/never married), region of residence (Northeast, Midwest, South, West, and Puerto Rico), body mass index (BMI: underweight, recommended, overweight, obesity), and menopausal status (premenopausal, postmenopausal). In models for weekly sleep duration, sleep medication use, daytime dysfunction, and healthcare professional diagnosed sleep apnea, reasons for trouble sleeping for any reason other than feeling too hot or feeling too cold ≥ 3 times per week in the past month include: unable to fall asleep within 30 minutes, waking up in the middle of the night or early morning, waking up to use the bathroom, cannot breathe comfortably, coughing or snoring loudly, having bad dreams, having pain, or other non-specified reasons. In models for long sleep onset latency, poor sleep maintenance, and insomnia symptoms, reasons for trouble sleeping for any reason other than feeling too hot or feeling too cold ≥ 3 times per week in the past month include waking up to use the bathroom, cannot breathe comfortably, coughing or snoring loudly, having bad dreams, having pain, or other non-specified reasons.

[a] 'Too hot' is defined as self-reported trouble sleeping due to feeling too hot ≥ 3 times per week vs. trouble sleeping due to feeling too hot < 3 times per week and trouble sleeping due to feeling too cold < 3 times per week.

[b] 'Too cold' is defined as self-reported trouble sleeping due to feeling too cold ≥ 3 times per week vs. trouble sleeping due to feeling too hot < 3 times per week and trouble sleeping due to feeling too cold < 3 times per week.

[c] Sleep duration is based on reported bed and wake times or reported average sleep duration. Participants who reported ≤ 2 hours or ≥ 23 hours of sleep are excluded. Short: <7 hours; Recommended: 7-9 hours; Long >9 hours

[d] Long sleep onset latency is defined as not falling asleep within 30 minutes at least three times a week during the past month.

[e] Poor sleep maintenance is defined as waking up in the middle of the night or early morning at least three times a week during the past month.

[f] Insomnia is defined as long sleep onset latency or poor sleep maintenance.

[g] Sleep medication use is defined as 'taking medicine (prescription or over the counter) to help you sleep' at least three times a week during the past month.

[h] Daytime dysfunction is defined as having trouble staying awake while driving, eating, or engaging in social activity at least three times a week during the past month.

[i] Healthcare provider diagnosed sleep apnea is defined as a current doctor or other health professional diagnosis of sleep apnea.

[ƚ] Remained significant after false discovery rate correction. False discovery rate-corrected P-value was considered statistically significant at the 0.05 level.

**Supplemental Table 7B. Relative excess risk due to interaction (RERI) between perceived indoor temperature extremes and race and ethnicity, Sister Study, 2017-2019, N=33,545**

| | **Indoor temperature: Too hot [a]** | | | **Indoor temperature: Too cold [b]** | | |
|---|---|---|---|---|---|---|
| | Hispanic/Latina | Non-Hispanic Black | Non-Hispanic White | Hispanic/Latina | Non-Hispanic Black | Non-Hispanic White |
| Weekly sleep duration [c] | | | | | | |
| Short | 0.17 (-0.55, 0.89) | -0.19 (-0.65, 0.27) | 1.00 (Ref) | -0.70 (-1.83, 0.43) | 0.37 (-0.93, 1.67) | 1.00 (Ref) |
| Recommended | --- | --- | | --- | --- | |
| Long | 0.30 (-0.32, 0.92) | -0.31 (-0.64, 0.02) | | -0.20 (-1.11, 0.71) | -0.11 (-1.24, 1.02) | |
| Long sleep onset latency [d] | | | | | | |
| Yes | **0.79 (0.06, 1.52)** | 0.19 (-0.20, 0.58) | 1.00 (Ref) | -0.20 (-1.20, 0.80) | -0.56 (-1.65, 0.53) | 1.00 (Ref) |
| No | --- | --- | | --- | --- | |
| Poor sleep maintenance [e] | | | | | | |
| Yes | 0.02 (-0.21, 0.25) | **-0.15 (-0.29, -0.01)** | 1.00 (Ref) | 0.20 (-0.21, 0.61) | 0.04 (-0.33, 0.41) | 1.00 (Ref) |
| No | --- | --- | | --- | --- | |
| Insomnia symptoms [f] | | | | | | |
| Yes | 0.12 (-0.07, 0.31) | **-0.13 (-0.26, -0.00)** | 1.00 (Ref) | 0.15 (-0.21, 0.51) | 0.10 (-0.25, 0.45) | 1.00 (Ref) |
| No | --- | --- | | --- | --- | |
| Sleep medication use [g] | | | | | | |
| Yes | -0.09 (-0.41, 0.23) | -0.03 (-0.25, 0.19) | 1.00 (Ref) | 0.02 (-0.47, 0.51) | -0.26 (-0.84, 0.32) | 1.00 (Ref) |
| No | --- | --- | | --- | --- | |
| Daytime dysfunction [h] | | | | | | |
| Yes | -1.19 (-4.02, 1.64) | 0.24 (-1.52, 2.00) | 1.00 (Ref) | 8.61 (-8.14, 25.36) | 6.21 (-5.18, 17.60) | 1.00 (Ref) |
| No | --- | --- | | --- | --- | |
| Healthcare professional diagnosed sleep apnea [i] | | | | | | |
| Yes | 0.22 (-0.36, 0.80) | 0.08 (-0.24, 0.40) | 1.00 (Ref) | 1.14 (-0.27, 2.55) | 0.81 (-0.36, 1.98) | 1.00 (Ref) |
| No | --- | --- | | --- | --- | |

Relative Excess Risk due to Interaction (RERI) = 0: no additive interaction; RERI >0: positive additive interaction; RERI <0: negative additive interaction

Models are adjusted for frequent trouble sleeping for any reason other than feeling too hot or feeling too cold, age (continuous), age$^2$, annual household income (<$20,000 - $49,999, $50,000 - $99,999, ≥$100,000), marital status (married or living as though married, divorced/widowed/separated/never married), region of residence (Northeast, Midwest, South, West, and Puerto Rico), body mass index (BMI: underweight, recommended, overweight, obesity), and menopausal status (premenopausal, postmenopausal). In models for weekly sleep duration, sleep medication use, daytime dysfunction, and healthcare professional diagnosed sleep apnea, reasons for trouble sleeping for any reason other than feeling too hot or feeling too cold ≥ 3 times per week in the past month include: unable to fall asleep within 30 minutes, waking up in the middle of the night or early morning, waking up to use the bathroom, cannot breathe comfortably, coughing or snoring loudly, having bad dreams, having pain, or other non-specified reasons. In models for long sleep onset latency, poor sleep maintenance, and insomnia symptoms, reasons for trouble sleeping for any reason other than feeling too hot or feeling too cold ≥ 3 times per week in the past month include waking up to use the bathroom, cannot breathe comfortably, coughing or snoring loudly, having bad dreams, having pain, or other non-specified reasons.

[a] 'Too hot' is defined as self-reported trouble sleeping due to feeling too hot ≥ 3 times per week vs. trouble sleeping due to feeling too hot < 3 times per week and trouble sleeping due to feeling too cold < 3 times per week.
[b] 'Too cold' is defined as self-reported trouble sleeping due to feeling too cold ≥ 3 times per week vs. trouble sleeping due to feeling too hot < 3 times per week and trouble sleeping due to feeling too cold < 3 times per week.

[c] Sleep duration is based on reported bed and wake times or reported average sleep duration. Participants who reported ≤ 2 hours or ≥ 23 hours of sleep are excluded. Short: <7 hours; Recommended: 7-9 hours; Long >9 hours
[d] Long sleep onset latency is defined as not falling asleep within 30 minutes at least three times a week during the past month.
[e] Poor sleep maintenance is defined as waking up in the middle of the night or early morning at least three times a week during the past month.
[f] Insomnia is defined as long sleep onset latency or poor sleep maintenance.
[g] Sleep medication use is defined as 'taking medicine (prescription or over the counter) to help you sleep' at least three times a week during the past month.
[h] Daytime dysfunction is defined as having trouble staying awake while driving, eating, or engaging in social activity at least three times a week during the past month.
[i] Healthcare provider diagnosed sleep apnea is defined as a current doctor or other health professional diagnosis of sleep apnea.

**Supplemental Table 8A. Prevalence ratios (95% confidence intervals) for associations between perceived indoor temperature extremes and sleep health dimensions by annual household income, Sister Study, 2017-2019, N = 33,545**

| | Indoor temperature: Too hot [a] | | | | Indoor temperature: Too cold [b] | | | |
|---|---|---|---|---|---|---|---|---|
| | <$20,000 - $49,999 | $50,000 - $99,999 | ≥ $100,000 | Wald p-values | <$20,000 - $49,999 | $50,000 - $99,999 | ≥ $100,000 | Wald p-values |
| Weekly sleep duration [c] | | | | | | | | |
| Short | 1.02 (0.84,1.24) | **1.20 (1.03,1.40)‡** | 1.02 (0.87,1.20) | 0.2546 | **1.47 (1.10,1.96)‡** | **1.49 (1.03,2.16)‡** | 1.41 (0.95,2.09) | 0.9772 |
| Recommended | 1.00 (Ref) | 1.00 (Ref) | 1.00 (Ref) | --- | 1.00 (Ref) | 1.00 (Ref) | 1.00 (Ref) | --- |
| Long | **1.22 (1.05,1.43)‡** | **1.17 (1.01,1.35)‡** | **1.35 (1.17,1.54)‡** | 0.3616 | **1.36 (1.05,1.76)‡** | 1.02 (0.68,1.53) | **1.73 (1.25,2.40)‡** | 0.1325 |
| Long sleep onset latency [d] | | | | | | | | |
| Yes | **1.89 (1.65,2.15)‡** | **2.02 (1.80,2.28)‡** | **1.73 (1.51,1.98)‡** | 0.2201 | **2.00 (1.62,2.48)‡** | **1.91 (1.40,2.60)‡** | **2.06 (1.44,2.94)‡** | 0.9480 |
| No | 1.00 (Ref) | 1.00 (Ref) | 1.00 (Ref) | --- | 1.00 (Ref) | 1.00 (Ref) | 1.00 (Ref) | --- |
| Poor sleep maintenance [e] | | | | | | | | |
| Yes | **1.54 (1.45,1.64)‡** | **1.54 (1.46,1.62)‡** | **1.62 (1.54,1.70)‡** | 0.2592 | **1.33 (1.18,1.50)‡** | **1.46 (1.26,1.68)‡** | **1.22 (1.04,1.44)‡** | 0.2815 |
| No | 1.00 (Ref) | 1.00 (Ref) | 1.00 (Ref) | --- | 1.00 (Ref) | 1.00 (Ref) | 1.00 (Ref) | --- |
| Insomnia symptoms [f] | | | | | | | | |
| Yes | **1.53 (1.45,1.62)‡** | **1.53 (1.46,1.60)‡** | **1.61 (1.54,1.68)‡** | 0.2131 | **1.32 (1.18,1.48)‡** | **1.41 (1.23,1.61)‡** | **1.27 (1.09,1.47)‡** | 0.5785 |
| No | 1.00 (Ref) | 1.00 (Ref) | 1.00 (Ref) | --- | 1.00 (Ref) | 1.00 (Ref) | 1.00 (Ref) | --- |
| Sleep medication use [g] | | | | | | | | |
| Yes | **1.29 (1.12,1.48)‡** | **1.19 (1.05,1.34)‡** | 1.10 (0.98,1.24) | 0.2363 | **1.36 (1.08,1.70)‡** | 1.00 (0.72,1.40) | **1.59 (1.22,2.09)‡** | 0.1085 |
| No | 1.00 (Ref) | 1.00 (Ref) | 1.00 (Ref) | --- | 1.00 (Ref) | 1.00 (Ref) | 1.00 (Ref) | --- |
| Daytime dysfunction [h] | | | | | | | | |
| Yes | 1.68 (0.89,3.16) | **2.01 (1.12,3.60)‡** | 1.61 (0.87,2.97) | 0.8570 | **2.76 (1.14,6.66)‡** | **7.96 (3.72,17.03)‡** | **5.24 (1.96,14.04)‡** | 0.2042 |
| No | 1.00 (Ref) | 1.00 (Ref) | 1.00 (Ref) | --- | 1.00 (Ref) | 1.00 (Ref) | 1.00 (Ref) | --- |
| Healthcare professional diagnosed sleep apnea [i] | | | | | | | | |
| Yes | **1.25 (1.04,1.50)‡** | **1.20 (1.02,1.41)‡** | 1.16 (0.97,1.38) | 0.8342 | **1.58 (1.19,2.09)‡** | **1.70 (1.17,2.45)‡** | **1.62 (1.06,2.47)‡** | 0.9554 |
| No | 1.00 (Ref) | 1.00 (Ref) | 1.00 (Ref) | --- | 1.00 (Ref) | 1.00 (Ref) | 1.00 (Ref) | --- |

Note: N=438 (1.3%) participants who reported both ‘too hot’ and ‘too cold’ temperatures were not included in the analysis.

Models are adjusted for trouble sleeping for any reason other than feeling too hot or feeling too cold, age (continuous), age$^2$, race and ethnicity (Hispanic/Latina, non-Hispanic Black, non-Hispanic White), marital status (married or living as though married, divorced/widowed/separated/never married), region of residence (Northeast, Midwest, South, West, and Puerto Rico), body mass index (BMI: underweight, recommended, overweight, obesity), and menopausal status (premenopausal, postmenopausal). In models for weekly sleep duration, sleep medication use, daytime dysfunction, and healthcare professional diagnosed sleep apnea, reasons for trouble sleeping for any reason other than feeling too hot or feeling too cold ≥ 3 times per week in the past month include: unable to fall asleep within 30 minutes, waking up in the middle of the night or early morning, waking up to use the bathroom, cannot breathe comfortably, coughing or snoring loudly, having bad dreams, having pain, or other non-specified reasons. In models for long sleep onset latency, poor sleep maintenance, and insomnia symptoms, reasons for trouble sleeping for any reason other than feeling too hot or feeling too cold ≥ 3 times per week in the past month include waking up to use the bathroom, cannot breathe comfortably, coughing or snoring loudly, having bad dreams, having pain, or other non-specified reasons.

[a] ’Too hot’ is defined as self-reported trouble sleeping due to feeling too hot ≥ 3 times per week vs. trouble sleeping due to feeling too hot < 3 times per week and trouble sleeping due to feeling too cold < 3 times per week.

[b] 'Too cold' is defined as self-reported trouble sleeping due to feeling too cold ≥ 3 times per week vs. trouble sleeping due to feeling too hot < 3 times per week and trouble sleeping due to feeling too cold < 3 times per week.

[c] Sleep duration is based on reported bed and wake times or reported average sleep duration. Participants who reported ≤ 2 hours or ≥ 23 hours of sleep are excluded. Short: <7 hours; Recommended: 7-9 hours; Long >9 hours

[d] Long sleep onset latency is defined as not falling asleep within 30 minutes at least three times a week during the past month.

[e] Poor sleep maintenance is defined as waking up in the middle of the night or early morning at least three times a week during the past month.

[f] Insomnia is defined as long sleep onset latency or poor sleep maintenance.

[g] Sleep medication use is defined as 'taking medicine (prescription or over the counter) to help you sleep' at least three times a week during the past month.

[h] Daytime dysfunction is defined as having trouble staying awake while driving, eating, or engaging in social activity at least three times a week during the past month.

[i] Healthcare provider diagnosed sleep apnea is defined as a current doctor or other health professional diagnosis of sleep apnea.

[ƚ] Remained significant after false discovery rate correction. False discovery rate-corrected P-value was considered statistically significant at the 0.05 level.

**Supplemental Table 8B. Relative excess risk due to interaction (RERI) between perceived temperature extremes and annual household income, Sister Study, 2017-2019, N = 33,545**

| | Indoor temperature: Too hot [a] | | | Indoor temperature: Too cold [b] | | |
|---|---|---|---|---|---|---|
| | <$20,000 - $49,999 | $50,000 - $99,999 | ≥ $100,000 | <$20,000 - $49,999 | $50,000 - $99,999 | ≥ $100,000 |
| Weekly sleep duration [c] | | | | | | |
| Short | 0.00 (-0.28, 0.28) | 0.19 (-0.05, 0.43) | 1.00 (Ref) | 0.17 (-0.58, 0.92) | 0.11 (-0.69, 0.91) | 1.00 (Ref) |
| Recommended | --- | --- | | --- | --- | |
| Long | -0.02 (-0.33, 0.29) | -0.15 (-0.41, 0.11) | | -0.21 (-0.96, 0.54) | -0.71 (-1.45, 0.03) | |
| Long sleep onset latency [d] | --- | --- | | --- | --- | |
| Yes | **0.58 (0.19, 0.97)** | **0.51 (0.18, 0.84)** | 1.00 (Ref) | 0.48 (-0.48, 1.44) | 0.06 (-0.95, 1.07) | 1.00 (Ref) |
| No | --- | --- | | --- | --- | |
| Poor sleep maintenance [e] | | | | | | |
| Yes | -0.08 (-0.18, 0.02) | **-0.09 (-0.18, -0.00)** | 1.00 (Ref) | 0.11 (-0.14, 0.36) | 0.23 (-0.05, 0.51) | 1.00 (Ref) |
| No | --- | --- | | --- | --- | |
| Insomnia symptoms [f] | | | | | | |
| Yes | -0.07 (-0.16, 0.02) | **-0.08 (-0.16, -0.00)** | 1.00 (Ref) | 0.06 (-0.18, 0.30) | 0.14 (-0.13, 0.41) | 1.00 (Ref) |
| No | --- | --- | | --- | --- | |
| Sleep medication use [g] | | | | | | |
| Yes | **0.24 (0.01, 0.47)** | 0.10 (-0.09, 0.29) | 1.00 (Ref) | -0.17 (-0.72, 0.38) | **-0.59 (-1.16, -0.02)** | 1.00 (Ref) |
| No | --- | --- | | --- | --- | |
| Daytime dysfunction [h] | | | | | | |
| Yes | 0.47 (-1.38, 2.32) | 0.48 (-0.98, 1.94) | 1.00 (Ref) | -1.41 (-7.74, 4.92) | 3.29 (-4.77, 11.35) | 1.00 (Ref) |
| No | --- | --- | | --- | --- | |
| Healthcare professional diagnosed sleep apnea [i] | | | | | | |
| Yes | 0.12 (-0.19, 0.43) | 0.05 (-0.23, 0.33) | 1.00 (Ref) | 0.03 (-0.81, 0.87) | 0.12 (-0.82, 1.06) | 1.00 (Ref) |
| No | --- | --- | | --- | --- | |

Relative Excess Risk due to Interaction (RERI) = 0: no additive interaction; RERI >0: positive additive interaction; RERI <0: negative additive interaction

Models are adjusted for trouble sleeping for any reason other than feeling too hot or feeling too cold, age (continuous), age$^2$, race and ethnicity (Hispanic/Latina, non-Hispanic Black, non-Hispanic White), marital status (married or living as though married, divorced/widowed/separated/never married), region of residence (Northeast, Midwest, South, West, and Puerto Rico), body mass index (BMI: underweight, recommended, overweight, obesity), and menopausal status (premenopausal, postmenopausal). In models for weekly sleep duration, sleep medication use, daytime dysfunction, and healthcare professional diagnosed sleep apnea, reasons for trouble sleeping for any reason other than feeling too hot or feeling too cold ≥ 3 times per week in the past month include: unable to fall asleep within 30 minutes, waking up in the middle of the night or early morning, waking up to use the bathroom, cannot breathe comfortably, coughing or snoring loudly, having bad dreams, having pain, or other non-specified reasons. In models for long sleep onset latency, poor sleep maintenance, and insomnia symptoms, reasons for trouble sleeping for any reason other than feeling too hot or feeling too cold ≥ 3 times per week in the past month include waking up to use the bathroom, cannot breathe comfortably, coughing or snoring loudly, having bad dreams, having pain, or other non-specified reasons.

[a] 'Too hot' is defined as self-reported trouble sleeping due to feeling too hot ≥ 3 times per week vs. trouble sleeping due to feeling too hot < 3 times per week and trouble sleeping due to feeling too cold < 3 times per week.
[b] 'Too cold' is defined as self-reported trouble sleeping due to feeling too cold ≥ 3 times per week vs. trouble sleeping due to feeling too hot < 3 times per week and trouble sleeping due to feeling too cold < 3 times per week.
[c] Sleep duration is based on reported bed and wake times or reported average sleep duration. Participants who reported ≤ 2 hours or ≥ 23 hours of sleep are excluded. Short: <7 hours; Recommended: 7-9 hours; Long >9 hours
[d] Long sleep onset latency is defined as not falling asleep within 30 minutes at least three times a week during the past month.
[e] Poor sleep maintenance is defined as waking up in the middle of the night or early morning at least three times a week during the past month.
[f] Insomnia is defined as long sleep onset latency or poor sleep maintenance.
[g] Sleep medication use is defined as 'taking medicine (prescription or over the counter) to help you sleep' at least three times a week during the past month.
[h] Daytime dysfunction is defined as having trouble staying awake while driving, eating, or engaging in social activity at least three times a week during the past month.
[i] Healthcare provider diagnosed sleep apnea is defined as a current doctor or other health professional diagnosis of sleep apnea.

**Supplemental Table 9. Relative excess risk due to interaction (RERI) between perceived indoor temperature extremes and region of residence, Sister Study, 2017-2019, N=33,545**

| | Indoor temperature: Too hot [a] | | | | | Indoor temperature: Too cold [b] | | | | |
|---|---|---|---|---|---|---|---|---|---|---|
| | Northeast | Midwest | South | West | Puerto Rico | Northeast | Midwest | South | West | Puerto Rico |
| Weekly sleep duration [c] | | | | | | | | | | |
| Short | -0.11 (-0.41, 0.19) | **0.41 (0.15, 0.67)** | | 0.17 (-0.10, 0.44) | 1.27 (-0.08, 2.62) | 0.83 (-0.21, 1.87) | -0.04 (-0.77, 0.69) | | -0.28 (-1.01, 0.45) | -0.52 (-1.51, 0.47) |
| Recommended | --- | --- | 1.00 (Ref) | --- | --- | --- | --- | 1.00 (Ref) | --- | --- |
| Long | -0.06 (-0.34, 0.22) | 0.08 (-0.16, 0.32) | | 0.17 (-0.09, 0.43) | **1.45 (0.48, 2.42)** | -0.10 (-0.87, 0.67) | -0.28 (-0.87, 0.31) | | 0.29 (-0.35, 0.93) | -0.11 (-0.93, 0.71) |
| Long sleep onset latency [d] | | | | | | | | | | |
| Yes | -0.27 (-0.63, 0.09) | **-0.32 (-0.63, -0.01)** | 1.00 (Ref) | -0.28 (-0.61, 0.05) | -0.50 (-1.48, 0.48) | -0.12 (-1.04, 0.80) | 0.47 (-0.31, 1.25) | 1.00 (Ref) | 0.19 (-0.60, 0.98) | 0.37 (-0.75, 1.49) |
| No | --- | --- | | --- | --- | --- | --- | | --- | --- |
| Poor sleep maintenance [e] | | | | | | | | | | |
| Yes | 0.08 (-0.03, 0.19) | 0.01 (-0.09, 0.11) | 1.00 (Ref) | 0.00 (-0.11, 0.11) | 0.17 (-0.32, 0.66) | -0.18 (-0.55, 0.19) | 0.06 (-0.22, 0.34) | 1.00 (Ref) | -0.22 (-0.51, 0.07) | 0.23 (-0.25, 0.71) |
| No | --- | --- | | --- | --- | --- | --- | | --- | --- |
| Insomnia symptoms [f] | | | | | | | | | | |
| Yes | 0.07 (-0.03, 0.17) | 0.00 (-0.09, 0.09) | 1.00 (Ref) | 0.02 (-0.07, 0.11) | 0.16 (-0.27, 0.59) | -0.22 (-0.56, 0.12) | 0.03 (-0.22, 0.28) | 1.00 (Ref) | -0.22 (-0.49, 0.05) | 0.21 (-0.21, 0.63) |
| No | --- | --- | | --- | --- | --- | --- | | --- | --- |
| Sleep medication use [g] | | | | | | | | | | |
| Yes | -0.03 (-0.27, 0.21) | -0.13 (-0.33, 0.07) | 1.00 (Ref) | -0.17 (-0.38, 0.04) | 0.14 (-1.06, 1.34) | 0.40 (-0.23, 1.03) | 0.22 (-0.29, 0.73) | 1.00 (Ref) | -0.19 (-0.69, 0.31) | 1.33 (-0.06, 2.72) |
| No | --- | --- | | --- | --- | --- | --- | | --- | --- |
| Daytime dysfunction [h] | | | | | | | | | | |
| Yes | --- | --- | 1.00 (Ref) | --- | --- | --- | --- | 1.00 (Ref) | --- | --- |
| No | --- | --- | | --- | --- | --- | --- | | --- | --- |
| Healthcare professional diagnosed sleep apnea [i] | | | | | | | | | | |
| Yes | -0.03 (-0.37, 0.31) | -0.16 (-0.45, 0.13) | 1.00 (Ref) | -0.22 (-0.55, 0.11) | 0.67 (-0.71, 2.05) | -0.49 (-1.40, 0.42) | **-0.96 (-1.73, -0.19)** | 1.00 (Ref) | -0.06 (-0.99, 0.87) | 0.93 (-0.68, 2.54) |
| No | --- | --- | | --- | --- | --- | --- | | --- | --- |

Relative Excess Risk due to Interaction (RERI) = 0: no additive interaction; RERI >0: positive additive interaction; RERI <0: negative additive interaction

Models are adjusted for trouble sleeping for any reason other than feeling too hot or feeling too cold, age (continuous), age$^2$, race and ethnicity (Hispanic/Latina, non-Hispanic Black, non-Hispanic White), annual household income (<$20,000 - $49,999, $50,000 - $99,999, ≥$100,000), body mass index (BMI: underweight, recommended, overweight, obesity), marital status (married or living as though married, divorced/widowed/separated/never married), and menopausal status (premenopausal, postmenopausal). In models for weekly sleep duration, sleep medication use, daytime dysfunction, and healthcare professional diagnosed sleep apnea, reasons for trouble sleeping for any reason other than feeling too hot or feeling too cold ≥ 3 times per week in the past month include: unable to fall asleep within 30 minutes, waking up in the middle of the night or early morning, waking up to use the bathroom, cannot breathe comfortably, coughing or snoring loudly, having bad dreams, having pain, or other non-specified reasons. In models for long sleep onset latency, poor sleep maintenance, and insomnia symptoms, reasons for trouble sleeping for any reason other than feeling too hot or feeling too cold ≥ 3 times per week in the past month include waking up to use the bathroom, cannot breathe comfortably, coughing or snoring loudly, having bad dreams, having pain, or other non-specified reasons.

[a] 'Too hot' is defined as self-reported trouble sleeping due to feeling too hot ≥ 3 times per week vs. trouble sleeping due to feeling too hot < 3 times per week and trouble sleeping due to feeling too cold < 3 times per week.
[b] 'Too cold' is defined as self-reported trouble sleeping due to feeling too cold ≥ 3 times per week vs. trouble sleeping due to feeling too hot < 3 times per week and trouble sleeping due to feeling too cold < 3 times per week.
[c] Sleep duration is based on reported bed and wake times or reported average sleep duration. Participants who reported ≤ 2 hours or ≥ 23 hours of sleep are excluded. Short: <7 hours; Recommended: 7-9 hours; Long >9 hours
[d] Long sleep onset latency is defined as not falling asleep within 30 minutes at least three times a week during the past month.
[e] Poor sleep maintenance is defined as waking up in the middle of the night or early morning at least three times a week during the past month.
[f] Insomnia is defined as long sleep onset latency or poor sleep maintenance.
[g] Sleep medication use is defined as 'taking medicine (prescription or over the counter) to help you sleep' at least three times a week during the past month.
[h] Daytime dysfunction is defined as having trouble staying awake while driving, eating, or engaging in social activity at least three times a week during the past month.

[i] Healthcare provider diagnosed sleep apnea is defined as a current doctor or other health professional diagnosis of sleep apnea.

**Supplemental Table 10. Relative excess risk due to interaction (RERI) between perceived indoor temperature extremes and menopausal status, Sister Study, 2017 – 2019, N = 33,545**

| | Indoor temperature: Too hot [a] | | Indoor temperature: Too cold [b] | |
|---|---|---|---|---|
| | **Premenopausal** | **Postmenopausal** | **Premenopausal** | **Postmenopausal** |
| Weekly sleep duration [c] | | | | |
| Short | | 0.12 (-0.25, 0.49) | | -0.05 (-1.12, 1.02) |
| Recommended | 1.0 (Ref) | --- | 1.0 (Ref) | --- |
| Long | | -0.13 (-0.65, 0.39) | | -0.00 (-1.42, 1.42) |
| Long sleep onset latency [d] | | | | |
| Yes | 1.0 (Ref) | -0.10 (-0.77, 0.57) | 1.0 (Ref) | **1.69 (0.43, 2.95)** |
| No | | --- | | --- |
| Poor sleep maintenance [e] | | | | |
| Yes | 1.0 (Ref) | -0.11 (-0.24, 0.02) | 1.0 (Ref) | -0.07 (-0.54, 0.40) |
| No | | --- | | --- |
| Insomnia symptoms [f] | | | | |
| Yes | 1.0 (Ref) | -0.10 (-0.20, 0.00) | 1.0 (Ref) | 0.01 (-0.42, 0.44) |
| No | | --- | | --- |
| Sleep medication use [g] | | | | |
| Yes | 1.0 (Ref) | 0.08 (-0.30, 0.46) | 1.0 (Ref) | -0.44 (-1.67, 0.79) |
| No | | --- | | --- |
| Daytime dysfunction [h] | | | | |
| Yes | 1.0 (Ref) | 1.29 (-0.94, 3.52) | 1.0 (Ref) | -8.85 (-29.77, 12.87) |
| No | | --- | | --- |
| Healthcare professional diagnosed sleep apnea [i] | | | | |
| Yes | 1.0 (Ref) | 0.24 (-0.26, 0.74) | 1.0 (Ref) | 0.56 (-0.99, 2.11) |
| No | | --- | | --- |

Relative Excess Risk due to Interaction (RERI) = 0: no additive interaction; RERI >0: positive additive interaction; RERI <0: negative additive interaction

Models are adjusted for trouble sleeping for any reason other than feeling too hot or feeling too cold, age (continuous), age$^2$, race and ethnicity (Hispanic/Latina, non-Hispanic Black, non-Hispanic White), annual household income (<$20,000 - $49,999, $50,000 - $99,999, ≥$100,000), marital status (married or living as though married, divorced/widowed/separated/never married), region of residence (Northeast, Midwest, South, West, and Puerto Rico), and body mass index (BMI: underweight, recommended, overweight, obesity). In models for weekly sleep duration, sleep medication use, daytime dysfunction, and healthcare professional diagnosed sleep apnea, reasons for trouble sleeping for any reason other than feeling too hot or feeling too cold ≥ 3 times per week in the past month include: unable to fall asleep within 30 minutes, waking up in the middle of the night or early morning, waking up to use the bathroom, cannot breathe comfortably, coughing or snoring loudly, having bad dreams, having pain, or other non-specified reasons. In models for long sleep onset latency, poor sleep maintenance, and insomnia symptoms, reasons for trouble sleeping for any reason other than feeling too hot or feeling too cold ≥ 3 times per week in the past month include waking up to use the bathroom, cannot breathe comfortably, coughing or snoring loudly, having bad dreams, having pain, or other non-specified reasons.

[a] 'Too hot' is defined as self-reported trouble sleeping due to feeling too hot ≥ 3 times per week vs. trouble sleeping due to feeling too hot < 3 times per week and trouble sleeping due to feeling too cold < 3 times per week.

[b] 'Too cold' is defined as self-reported trouble sleeping due to feeling too cold ≥ 3 times per week vs. trouble sleeping due to feeling too hot < 3 times per week and trouble sleeping due to feeling too cold < 3 times per week.
[c] Sleep duration is based on reported bed and wake times or reported average sleep duration. Participants who reported ≤ 2 hours or ≥ 23 hours of sleep are excluded. Short: <7 hours; Recommended: 7-9 hours; Long >9 hours
[d] Long sleep onset latency is defined as not falling asleep within 30 minutes at least three times a week during the past month.
[e] Poor sleep maintenance is defined as waking up in the middle of the night or early morning at least three times a week during the past month.
[f] Insomnia is defined as long sleep onset latency or poor sleep maintenance.
[g] Sleep medication use is defined as 'taking medicine (prescription or over the counter) to help you sleep' at least three times a week during the past month.
[h] Daytime dysfunction is defined as having trouble staying awake while driving, eating, or engaging in social activity at least three times a week during the past month.
[i] Healthcare provider diagnosed sleep apnea is defined as a current doctor or other health professional diagnosis of sleep apnea.

**Supplemental Table 11. Sociodemographic, clinical, and sleep health characteristics among participants who reported trouble sleeping for any reason [a], overall and by perceived indoor temperature extremes, Sister Study, 2017-2019, N = 23,853**

| | Total N = 23,853 (100.0) | Infrequent temperature extremes [b] n = 20,347 (85.3) | Too hot [c] n = 2,682 (11.2) | Too cold [d] n = 406 (1.7) |
|---|---|---|---|---|
| **Sociodemographic characteristics** | | | | |
| Age, years (mean ± SD) | 67.61 ± 8.45 | 68.31 ± 8.31 | 62.78 ± 7.65 | 68.90 ± 9.04 |
| Age (categorical years) | | | | |
| < 67.2 | 11,411 (47.8) | 9,025 (44.4) | 1,920 (71.6) | 164 (40.4) |
| ≥ 67.2 | 12,442 (52.2) | 11,322 (55.6) | 762 (28.4) | 242 (59.6) |
| Race and ethnicity | | | | |
| Hispanic/Latina | 783 (3.3) | 622 (3.1) | 100 (3.7) | 36 (8.9) |
| Non-Hispanic Black | 1,494 (6.3) | 1,150 (5.7) | 276 (10.3) | 31 (7.6) |
| Non-Hispanic White | 21,576 (90.5) | 18,575 (91.3) | 2,306 (86.0) | 339 (83.5) |
| Educational attainment [e] | | | | |
| ≤ High school or GED | 3,205 (13.4) | 2,684 (13.2) | 384 (14.3) | 79 (19.5) |
| Some college/technical | 7,603 (31.9) | 6,394 (31.4) | 913 (34.0) | 150 (36.9) |
| ≥ Bachelor's degree or higher | 13,042 (54.7) | 11,267 (55.4) | 1,385 (51.6) | 177 (43.6) |
| *Missing* | 3 (0.0) | 2 (0.0) | 0 (0.0) | 0 (0.0) |
| Annual household income | | | | |
| < $20,000 - $49,999 | 6,614 (27.7) | 5,741 (28.2) | 585 (21.8) | 176 (43.3) |
| $50,000 - $99,999 | 8,820 (37.0) | 7,605 (37.4) | 943 (35.2) | 119 (29.3) |
| ≥ $100,000 | 8,419 (35.3) | 7,001 (34.4) | 1,154 (43.0) | 111 (27.3) |
| Marital status | | | | |
| Married or living as though married | 16,124 (67.6) | 13,508 (66.4) | 2,065 (77.0) | 251 (61.8) |
| Never married, divorced, widowed, or separated | 7,729 (32.4) | 6,839 (33.6) | 617 (23.0) | 155 (38.2) |
| Region of residence | | | | |
| Northeast | 3,876 (16.2) | 3,334 (16.4) | 430 (16.0) | 55 (13.5) |
| Midwest | 6,290 (26.4) | 5,375 (26.4) | 702 (26.2) | 101 (24.9) |
| South | 7,916 (33.2) | 6,722 (33.0) | 921 (34.3) | 131 (32.3) |
| West | 5,522 (23.2) | 4,725 (23.2) | 605 (22.6) | 96 (23.6) |
| Puerto Rico | 249 (1.0) | 191 (0.9) | 24 (0.9) | 23 (5.7) |
| **Clinical characteristics** | | | | |
| Body Mass Index, kg/$m^2$ | | | | |
| Recommended (18.5 - 24.9) | 9,092 (38.1) | 7,891 (38.8) | 877 (32.7) | 166 (40.9) |

| | | | | |
|---|---|---|---|---|
| Overweight (25.0 - 29.9) | 7,633 (32.0) | 6,507 (32.0) | 895 (33.4) | 101 (24.9) |
| Obesity (≥ 30) | 6,823 (28.6) | 5,691 (28.0) | 878 (32.7) | 133 (32.8) |
| *Missing* | 305 (1.3) | 258 (1.3) | 32 (1.2) | 6 (1.5) |
| Menopausal status | | | | |
| Premenopausal | 791 (3.3) | 588 (2.9) | 158 (5.9) | 20 (4.9) |
| Postmenopausal | 23,057 (96.7) | 19,754 (97.1) | 2,524 (94.1) | 386 (95.1) |
| *Missing or never had period* | 5 (0.0) | 5 (0.0) | 0 (0.0) | 0 (0.0) |
| **Sleep health dimensions** | | | | |
| Average weekly sleep duration [f] | | | | |
| Short (< 7) | 2,844 (11.9) | 2,348 (11.5) | 369 (13.8) | 67 (16.5) |
| Recommended (7-9) | 16,898 (70.8) | 14,567 (71.6) | 1,825 (68.0) | 242 (59.6) |
| Long (> 9) | 3,819 (16.0) | 3,188 (15.7) | 461 (17.2) | 83 (20.4) |
| *Missing* | 292 (1.2) | 244 (1.2) | 27 (1.0) | 14 (3.4) |
| Long sleep onset latency [g] | | | | |
| Yes | 4,183 (17.5) | 3,197 (15.7) | 689 (25.7) | 120 (29.6) |
| No | 19,462 (81.6) | 16,977 (83.4) | 1,973 (73.6) | 278 (68.5) |
| *Missing* | 208 (0.9) | 173 (0.9) | 20 (0.7) | 8 (2.0) |
| Poor sleep maintenance [h] | | | | |
| Yes | 13,260 (55.6) | 10,712 (52.6) | 1,964 (73.2) | 262 (64.5) |
| No | 10,403 (43.6) | 9,465 (46.5) | 705 (26.3) | 142 (35.0) |
| *Missing* | 190 (0.8) | 170 (0.8) | 13 (0.5) | 2 (0.5) |
| Insomnia symptoms [i] | | | | |
| Yes | 14,355 (60.2) | 11,621 (57.1) | 2,102 (78.4) | 279 (68.7) |
| No | 9,431 (39.5) | 8,667 (42.6) | 577 (21.5) | 125 (30.8) |
| *Missing* | 67 (0.3) | 59 (0.3) | 3 (0.1) | 2 (0.5) |
| Sleep medication use [j] | | | | |
| Yes | 5,077 (21.3) | 4,219 (20.7) | 623 (23.2) | 114 (28.1) |
| No | 18,674 (78.3) | 16,047 (78.9) | 2,047 (76.3) | 288 (70.9) |
| *Missing* | 102 (0.4) | 81 (0.4) | 12 (0.4) | 4 (1.0) |
| Daytime dysfunction [k] | | | | |
| Yes | 219 (0.9) | 155 (0.8) | 36 (1.3) | 16 (3.9) |
| No | 23,356 (97.9) | 19,953 (98.1) | 2,615 (97.5) | 386 (95.1) |
| *Missing* | 278 (1.2) | 239 (1.2) | 31 (1.2) | 4 (1.0) |
| Healthcare provider diagnosed sleep apnea [l] | | | | |
| Yes | 2,564 (10.7) | 2,117 (10.4) | 325 (12.1) | 71 (17.5) |

| | | | | |
|---|---|---|---|---|
| No | 21,023 (88.1) | 18,002 (88.5) | 2,338 (87.2) | 324 (79.8) |
| *Missing* | 266 (1.1) | 228 (1.1) | 19 (0.7) | 11 (2.7) |

Abbreviations: SD, Standard Deviation; GED: General Educational Development
Note: N = 418 (1.8%) participants who reported both 'too hot' and 'too cold' temperatures were not included in this sensitivity analysis.
[a] Reasons for trouble sleeping ≥ 3 times per week in the past month include: unable to fall asleep within 30 minutes, waking up in the middle of the night or early morning, waking up to use the bathroom, cannot breathe comfortably, coughing or snoring loudly, having bad dreams, having pain, or other non-specified reasons.
[b] ''Infrequent temperature extremes' is defined as self-reported trouble sleeping due to feeling too hot < 3 times per week and trouble sleeping due to feeling too cold < 3 times per week in the past month.
[c] 'Too hot' is defined as self-reported trouble sleeping due to feeling too hot ≥ 3 times per week in the past month.
[d] 'Too cold' is defined as self-reported trouble sleeping due to feeling too cold ≥ 3 times per week in the past month.
[e] Educational attainment was assessed at baseline.
[f] Sleep duration is based on reported bed and wake times or reported average sleep duration. Participants who reported ≤ 2 or ≥ 23 hours of sleep are excluded. Short: < 7 hours; Recommended: 7-9 hours; Long ≥ 9 hours.
[g] Long sleep onset latency is defined as not falling asleep within 30 minutes at least three times a week during the past month.
[h] Poor sleep maintenance is defined as waking up in the middle of the night or early morning at least three times a week during the past month.
[i] Insomnia symptoms is defined as long sleep onset latency or poor sleep maintenance.
[j] Sleep medication use is defined as 'taking medicine (prescription or over the counter) to help you sleep' at least three times a week during the past month.
[k] Daytime dysfunction is defined as having trouble staying awake while driving, eating, or engaging in social activity at least three times a week during the past month.
[l] Healthcare provider diagnosed sleep apnea is defined as a current doctor or other health professional diagnosis of sleep apnea.

**Supplemental Table 12. Sociodemographic, clinical, and sleep health characteristics among participants who reported trouble sleeping for any reason [a], by race and ethnicity as well as perceived indoor temperature extremes, Sister Study, 2017-2019, N = 23,853**

| Race and ethnicity | Hispanic/Latina [b] | | | | Non-Hispanic Black [c] | | | | Non-Hispanic White [d] | | | |
|---|---|---|---|---|---|---|---|---|---|---|---|---|
| | Total n =783 (3.3%) | Infrequent temperature extremes [e] n = 622 (79.4%) | Too hot [f ¥] n = 100 (12.8%) | Too cold [g §] n = 36 (4.6%) | Total n = 1,494 (6.3%) | Infrequent temperature extremes [e] n = 1,150 (77.0%) | Too hot [f ¥] n = 276 (18.5%) | Too cold [g §] n = 31 (2.1%) | Total n = 21,576 (90.5%) | Infrequent temperature extremes [e] n = 18,575 (86.1%) | Too hot [f ¥] n = 2,306 (10.7%) | Too cold [g §] n = 339 (1.6%) |
| **Sociodemographic characteristics** | | | | | | | | | | | | |
| Age, years [ǂ] | 64.20 ± 8.55 | 64.85 ± 8.50 | 60.35 ± 7.80 | 66.00 ± 8.96 | 65.68 ± 8.16 | 66.60 ± 8.03 | 62.49 ± 7.62 | 65.04 ± 10.43 | 67.86 ± 8.42 | 68.53 ± 8.29 | 62.92 ± 7.63 | 69.56 ± 8.78 |
| Age, years [ǂ] | | | | | | | | | | | | |
| < 67.2 | 502 (64.1) | 381 (61.3) | 82 (82.0) | 21 (58.3) | 839 (56.2) | 599 (52.1) | 197 (71.4) | 15 (48.4) | 10,070 (46.7) | 8,045 (43.3) | 1,641 (71.2) | 128 (37.8) |
| ≥ 67.2 | 281 (35.9) | 241 (38.7) | 18 (18.0) | 15 (41.7) | 655 (43.8) | 551 (47.9) | 79 (28.6) | 16 (51.6) | 11,506 (53.3) | 10,530 (56.7) | 665 (28.8) | 211 (62.2) |
| Educational attainment [ǂ h] | | | | | | | | | | | | |
| < High school or GED | 143 (18.3) | 108 (17.4) | 19 (19.0) | 9 (25.0) | 121 (8.1) | 88 (7.7) | 25 (9.1) | 7 (22.6) | 2,941 (13.6) | 2,488 (13.4) | 340 (14.7) | 63 (18.6) |
| Some college / technical | 256 (32.7) | 196 (31.5) | 37 (37.0) | 11 (30.6) | 469 (31.4) | 344 (29.9) | 101 (36.6) | 10 (32.3) | 6,878 (31.9) | 5,854 (31.5) | 775 (33.6) | 129 (38.1) |
| ≥ Bachelor's or higher | 384 (49.0) | 318 (51.1) | 44 (44.0) | 16 (44.4) | 904 (60.5) | 718 (62.4) | 150 (54.3) | 14 (45.2) | 11,754 (54.5) | 10,231 (55.1) | 1,191 (51.6) | 147 (43.4) |
| *Missing* | *0 (0.0)* | *0 (0.0)* | *0 (0.0)* | *0 (0.0)* | *0 (0.0)* | *0 (0.0)* | *0 (0.0)* | *0 (0.0)* | *3 (0.0)* | *2 (0.0)* | *0 (0.0)* | *0 (0.0)* |
| Annual household income [ǂ] | | | | | | | | | | | | |
| < $20,000 - $49,999 | 386 (49.3) | 301 (48.4) | 38 (38.0) | 31 (86.1) | 536 (35.9) | 426 (37.0) | 79 (28.6) | 16 (51.6) | 5,692 (26.4) | 5,014 (27.0) | 468 (20.3) | 129 (38.1) |
| $50,000 - $99,999 | 209 (26.7) | 170 (27.3) | 29 (29.0) | 3 (8.3) | 568 (38.0) | 427 (37.1) | 121 (43.8) | 6 (19.4) | 8,043 (37.3) | 7,008 (37.7) | 793 (34.4) | 110 (32.4) |
| ≥ $100,000 | 188 (24.0) | 151 (24.3) | 33 (33.0) | 2 (5.6) | 390 (26.1) | 297 (25.8) | 76 (27.5) | 9 (29.0) | 7,841 (36.3) | 6,553 (35.3) | 1,045 (45.3) | 100 (29.5) |
| Marital status [ǂ] | | | | | | | | | | | | |
| Married or living as though married | 499 (63.7) | 388 (62.4) | 74 (74.0) | 18 (50.0) | 681 (45.6) | 499 (43.4) | 157 (56.9) | 9 (29.0) | 14,944 (69.3) | 12,621 67.9) | 1,834 (79.5) | 224 (66.1) |
| Never married, divorced, widowed, or separated | 284 (36.3) | 234 (37.6) | 26 (26.0) | 18 (50.0) | 813 (54.4) | 651 (56.6) | 119 (43.1) | 22 (71.0) | 6,632 (30.7) | 5,954 (32.1) | 472 (20.5) | 115 (33.9) |
| Region of residence [ǂ] | | | | | | | | | | | | |
| Northeast | 63 (8.0) | 45 (7.2) | 14 (14.0) | 1 (2.8) | 129 (8.6) | 102 (8.9) | 22 (8.0) | 2 (6.5) | 3,684 (17.1) | 3,187 (17.2) | 394 (17.1) | 52 (15.3) |
| Midwest | 47 (6.0) | 41 (6.6) | 6 (6.0) | 0 (0.0) | 284 (19.0) | 223 (19.4) | 50 (18.1) | 6 (19.4) | 5,959 (27.6) | 5,111 (27.5) | 646 (28.0) | 95 (28.0) |
| South | 212 (27.1) | 172 (27.7) | 28 (28.0) | 7 (19.4) | 953 (63.8) | 718 (62.4) | 189 (68.5) | 18 (58.1) | 6,751 (31.3) | 5,832 (31.4) | 704 (30.5) | 106 (31.3) |
| West | 215 (27.5) | 176 (28.3) | 28 (28.0) | 5 (13.9) | 128 (8.6) | 107 (9.3) | 15 (5.4) | 5 (16.1) | 5,179 (24.0) | 4,442 (23.9) | 562 (24.4) | 86 (25.4) |
| Puerto Rico | 246 (31.4) | 188 (30.2) | 24 (24.0) | 23 (63.9) | 0 (0.0) | 0 (0.0) | 0 (0.0) | 0 (0.0) | 3 (0.0) | 3 (0.0) | 0 (0.0) | 0 (0.0) |
| **Clinical characteristics** | | | | | | | | | | | | |
| Body Mass Index, kg/m$^2$ [ǂ] | | | | | | | | | | | | |
| Recommended (18.5 - 24.9) | 241 (30.8) | 196 (31.5) | 30 (30.0) | 10 (27.8) | 233 (15.6) | 182 (15.8) | 42 (15.2) | 5 (16.1) | 8,618 (39.9) | 7,513 (40.4) | 805 (34.9) | 151 (44.5) |
| Overweight (25.0 - 29.9) | 273 (34.9) | 215 (34.6) | 33 (33.0) | 14 (38.9) | 488 (32.7) | 385 (33.5) | 79 (28.6) | 9 (29.0) | 6,872 (31.9) | 5,907 (31.8) | 783 (34.0) | 78 (23.0) |
| Obesity (≥ 30) | 261 (33.3) | 205 (33.0) | 36 (36.0) | 12 (33.3) | 756 (50.6) | 570 (49.6) | 154 (55.8) | 16 (51.6) | 5,806 (26.9) | 4,916 (26.5) | 688 (29.8) | 105 (31.0) |
| *Missing* | *8 (1.0)* | *6 (1.0)* | *1 (1.0)* | *0 (0.0)* | *17 (1.1)* | *13 (1.1)* | *1 (0.4)* | *1 (3.2)* | *280 (1.3)* | *239 (1.3)* | *30 (1.3)* | *5 (1.5)* |
| Menopausal status [ǂ] | | | | | | | | | | | | |
| Premenopausal | 50 (6.4) | 42 (6.8) | 7 (7.0) | 1 (2.8) | 85 (5.7) | 53 (4.6) | 24 (8.7) | 5 (16.1) | 656 (3.0) | 493 (2.7) | 127 (5.5) | 14 (4.1) |
| Postmenopausal | 733 (93.6) | 580 (93.2) | 93 (93.0) | 35 (97.2) | 1,407 (94.2) | 1,095 (95.2) | 252 (91.3) | 26 (83.9) | 20,917 (96.9) | 18,079 (97.3) | 2,179 (94.5) | 325 (95.9) |

| | | | | | | | | | | | | |
|---|---|---|---|---|---|---|---|---|---|---|---|---|
| *Missing or never had a period* | *0 (0.0)* | *0 (0.0)* | *0 (0.0)* | *0 (0.0)* | *2 (0.1)* | *2 (0.2)* | *0 (0.0)* | *0 (0.0)* | *3 (0.0)* | *3 (0.0)* | *0 (0.0)* | *0 (0.0)* |
| **Sleep health dimensions** | | | | | | | | | | | | |
| Average weekly sleep duration ǂ [i] | | | | | | | | | | | | |
| Short | 128 (16.3) | 102 (16.4) | 17 (17.0) | 4 (11.1) | 450 (30.1) | 336 (29.2) | 84 (30.4) | 14 (45.2) | 2,266 (10.5) | 1,910 (10.3) | 268 (11.6) | 49 (14.5) |
| Recommended | 502 (64.1) | 408 (65.6) | 60 (60.0) | 22 (61.1) | 817 (54.7) | 639 (55.6) | 151 (54.7) | 12 (38.7) | 15,579 (72.2) | 13,520 (72.8) | 1,614 (70.0) | 208 (61.4) |
| Long | 132 (16.9) | 99 (15.9) | 20 (20.0) | 6 (16.7) | 195 (13.1) | 153 (13.3) | 33 (12.0) | 4 (12.9) | 3,492 (16.2) | 2,936 (15.8) | 408 (17.7) | 73 (21.5) |
| *Missing* | *21 (2.7)* | *13 (2.1)* | *3 (3.0)* | *4 (11.1)* | *32 (2.1)* | *22 (1.9)* | *8 (2.9)* | *1 (3.2)* | *239 (1.1)* | *209 (1.1)* | *16 (0.7)* | *9 (2.7)* |
| Long sleep onset latency ǂ [j] | | | | | | | | | | | | |
| Yes | 216 (27.6) | 146 (23.5) | 41 (41.0) | 14 (38.9) | 327 (21.9) | 210 (18.3) | 90 (32.6) | 7 (22.6) | 3,640 (16.9) | 2,841 (15.3) | 558 (24.2) | 99 (29.2) |
| No | 545 (69.6) | 462 (74.3) | 55 (55.0) | 19 (52.8) | 1,144 (76.6) | 920 (80.0) | 184 (66.7) | 23 (74.2) | 17,773 (82.4) | 15,595 (84.0) | 1,734 (75.2) | 236 (69.6) |
| *Missing* | *22 (2.8)* | *14 (2.3)* | *4 (4.0)* | *3 (8.3)* | *23 (1.5)* | *20 (1.7)* | *2 (0.7)* | *1 (3.2)* | *163 (0.8)* | *139 (0.7)* | *14 (0.6)* | *4 (1.2)* |
| Poor sleep maintenance [k] | | | | | | | | | | | | |
| Yes | 410 (52.4) | 300 (48.2) | 68 (68.0) | 23 (63.9) | 793 (53.1) | 568 (49.4) | 179 (64.9) | 19 (61.3) | 12,057 (55.9) | 9,844 (53.0) | 1,717 (74.5) | 220 (64.9) |
| No | 355 (45.3) | 308 (49.5) | 31 (31.0) | 11 (30.6) | 685 (45.9) | 569 (49.5) | 94 (34.1) | 12 (38.7) | 9,363 (43.4) | 8,588 (46.2) | 580 (25.2) | 119 (35.1) |
| *Missing* | *18 (2.3)* | *14 (2.3)* | *1 (1.0)* | *2 (5.6)* | *16 (1.1)* | *13 (1.1)* | *3 (1.1)* | *0 (0.0)* | *156 (0.7)* | *143 (0.8)* | *9 (0.4)* | *0 (0.0)* |
| Insomnia symptoms [l] | | | | | | | | | | | | |
| Yes | 465 (59.4) | 342 (55.0) | 79 (79.0) | 25 (69.4) | 864 (57.8) | 616 (53.6) | 195 (70.7) | 21 (67.7) | 13,026 (60.4) | 10,663 (57.4) | 1,828 (79.3) | 233 (68.7) |
| No | 305 (39.0) | 271 (43.6) | 20 (20.0) | 9 (25.0) | 625 (41.8) | 529 (46.0) | 81 (29.3) | 10 (32.3) | 8,501 (39.4) | 7,867 (42.4) | 476 (20.6) | 106 (31.3) |
| *Missing* | *13 (1.7)* | *9 (1.4)* | *1 (1.0)* | *2 (5.6)* | *5 (0.3)* | *5 (0.4)* | *0 (0.0)* | *0 (0.0)* | *49 (0.2)* | *45 (0.2)* | *2 (0.1)* | *0 (0.0)* |
| Sleep medication use ǂ [m] | | | | | | | | | | | | |
| Yes | 152 (19.4) | 113 (18.2) | 18 (18.0) | 10 (27.8) | 204 (13.7) | 151 (13.1) | 40 (14.5) | 5 (16.1) | 4,721 (21.9) | 3,955 (21.3) | 565 (24.5) | 99 (29.2) |
| No | 615 (78.5) | 499 (80.2) | 82 (82.0) | 23 (63.9) | 1,280 (85.7) | 993 (86.3) | 233 (84.4) | 26 (83.9) | 16,779 (77.8) | 14,555 (78.4) | 1,732 (75.1) | 239 (70.5) |
| *Missing* | *16 (2.0)* | *10 (1.6)* | *0 (0.0)* | *3 (8.3)* | *10 (0.7)* | *6 (0.5)* | *3 (1.1)* | *0 (0.0)* | *76 (0.4)* | *65 (0.3)* | *9 (0.4)* | *1 (0.3)* |
| Daytime dysfunction [n] | | | | | | | | | | | | |
| Yes | 13 (1.7) | 8 (1.3) | 1 (1.0) | 2 (5.6) | 16 (1.1) | 9 (0.8) | 4 (1.4) | 3 (9.7) | 190 (0.9) | 138 (0.7) | 31 (1.3) | 11 (3.2) |
| No | 754 (96.3) | 601 (96.6) | 98 (98.0) | 33 (91.7) | 1,463 (97.9) | 1,130 (98.3) | 268 (97.1) | 28 (90.3) | 21,139 (98.0) | 18,222 (98.1) | 2,249 (97.5) | 325 (95.9) |
| *Missing* | *16 (2.0)* | *13 (2.1)* | *1 (1.0)* | *1 (2.8)* | *15 (1.0)* | *11 (1.0)* | *4 (1.4)* | *0 (0.0)* | *247 (1.1)* | *215 (1.2)* | *26 (1.1)* | *3 (0.9)* |
| Healthcare provider diagnosed sleep apnea ǂ [o] | | | | | | | | | | | | |
| Yes | 77 (9.8) | 55 (8.8) | 12 (12.0) | 7 (19.4) | 239 (16.0) | 170 (14.8) | 51 (18.5) | 9 (29.0) | 2,248 (10.4) | 1,892 (10.2) | 262 (11.4) | 55 (16.2) |
| No | 693 (88.5) | 558 (89.7) | 88 (88.0) | 28 (77.8) | 1,225 (82.0) | 956 (83.1) | 220 (79.7) | 21 (67.7) | 19,105 (88.5) | 16,488 (88.8) | 2,030 (88.0) | 275 (81.1) |
| *Missing* | *13 (1.7)* | *9 (1.4)* | *0 (0.0)* | *1 (2.8)* | *30 (2.0)* | *24 (2.1)* | *5 (1.8)* | *1 (3.2)* | *223 (1.0)* | *195 (1.0)* | *14 (0.6)* | *9 (2.7)* |

Abbreviations: SD, Standard Deviation; GED: General Educational Development

Note: N = 418 (1.8%) participants who reported both 'too hot' and 'too cold' temperatures were not included in this sensitivity analysis.

[¥] Chi-square test p-values indicate significant differences between the overall prevalence of ‘too hot’ by race and ethnicity.

[§] Chi-square test p-values indicate significant differences between the overall prevalence of ‘too cold’ by race and ethnicity.

[ǂ] Chi-square or Analysis of Variance (ANOVA) test p-values indicate significant differences in characteristics between the total populations of each race and ethnicity.

[a] Reasons for trouble sleeping ≥ 3 times per week in the past month include: unable to fall asleep within 30 minutes, waking up in the middle of the night or early morning, waking up to use the bathroom, cannot breathe comfortably, coughing or snoring loudly, having bad dreams, having pain, or other non-specified reasons.

[b] Excluded for reporting both ‘Too hot’ and ‘Too cold’, Hispanic/Latina: n = 25 (3.2%).

[c] Excluded for reporting both ‘Too hot’ and ‘Too cold’, Non-Hispanic Black: n = 37 (2.5%).

[d] Excluded for reporting both ‘Too hot’ and ‘Too cold’, Non-Hispanic White: n = 356 (1.6%).

[e] ‘Infrequent temperature extremes’ is defined as self-reported trouble sleeping due to feeling too hot < 3 times per week and trouble sleeping due to feeling too cold < 3 times per week in the past month.

[f] ’Too hot’ is defined as self-reported trouble sleeping due to feeling too hot ≥ 3 times per week in the past month.

[g] ’Too cold’ is defined as self-reported trouble sleeping due to feeling too cold ≥ 3 times per week in the past month.

[h] Educational attainment was assessed at baseline.
[i] Sleep duration is based on reported bed and wake times or reported average sleep duration. Participants who reported ≤ 2 or ≥ 23 hours of sleep are excluded. Short: < 7 hours; Recommended: 7-9 hours; Long ≥ 9 hours.
[j] Long sleep onset latency is defined as not falling asleep within 30 minutes at least three times a week during the past month.
[k] Poor sleep maintenance is defined as waking up in the middle of the night or early morning at least three times a week during the past month.
[l] Insomnia symptoms is defined as long sleep onset latency or poor sleep maintenance.
[m] Sleep medication use is defined as 'taking medicine (prescription or over the counter) to help you sleep' at least three times a week during the past month.
[n] Daytime dysfunction is defined as having trouble staying awake while driving, eating, or engaging in social activity at least three times a week during the past month.
[o] Healthcare provider diagnosed sleep apnea is defined as a current doctor or other health professional diagnosis of sleep apnea.

**Supplemental Table 13. Sociodemographic, clinical, and sleep health characteristics among participants who reported trouble sleeping for any reason [a], by annual household income as well as perceived indoor temperature extremes, Sister Study, 2017-2019, N = 23,853**

| Annual household income | < $20,000 - $49,999 [b] | | | | $50,000 - $99,999 [c] | | | | ≥ $100,000 [d] | | | |
|---|---|---|---|---|---|---|---|---|---|---|---|---|
| | Total n = 6,614 (27.7%) | Infrequent temperature extremes [e] n = 5,741 (86.8%) | Too hot [f ¥] n = 585 (8.8%) | Too cold [g §] n = 176 (2.7%) | Total n = 8,820 (37.0%) | Infrequent temperature extremes [e] n = 7,605 (86.2%) | Too hot [f ¥] n = 943 (10.7%) | Too cold [g §] n = 119 (1.3%) | Total n = 8,419 (35.3%) | Infrequent temperature extremes [e] n = 7,001 (83.2%) | Too hot [f ¥] n = 1,154 (13.7%) | Too cold [g §] n = 111 (1.3%) |
| **Sociodemographic characteristics** | | | | | | | | | | | | |
| Age, years (mean ± SD) [ǂ] | 70.79 ± 8.14 | 71.43 ± 7.88 | 65.33 ± 7.97 | 71.39 ± 8.96 | 68.07 ± 7.96 | 68.70 ± 7.83 | 63.81 ± 7.33 | 67.74 ± 8.47 | 64.62 ± 8.18 | 65.33 ± 8.14 | 60.64 ± 7.16 | 66.20 ± 8.79 |
| Age, years [ǂ] | | | | | | | | | | | | |
| < 67.2 | 2,154 (32.6) | 1,682 (29.3) | 348 (59.5) | 56 (31.8) | 3,992 (45.3) | 3,207 (42.2) | 626 (66.4) | 52 (43.7) | 5,265 (62.5) | 4,136 (59.1) | 946 (82.0) | 56 (50.5) |
| ≥ 67.2 | 4,460 (67.4) | 4,059 (70.7) | 237 (40.5) | 120 (68.2) | 4,828 (54.7) | 4,398 (57.8) | 317 (33.6) | 67 (56.3) | 3,154 (37.5) | 2,865 (40.9) | 208 (18.0) | 55 (49.5) |
| Race and ethnicity [ǂ] | | | | | | | | | | | | |
| Hispanic/Latina | 386 (5.8) | 301 (5.2) | 38 (6.5) | 31 (17.6) | 209 (2.4) | 170 (2.2) | 29 (3.1) | 3 (2.5) | 188 (2.2) | 151 (2.2) | 33 (2.9) | 2 (1.8) |
| Non-Hispanic Black | 536 (8.1) | 426 (7.4) | 79 (13.5) | 16 (9.1) | 568 (6.4) | 427 (5.6) | 121 (12.8) | 6 (5.0) | 390 (4.6) | 297 (4.2) | 76 (6.6) | 9 (8.1) |
| Non-Hispanic White | 5,692 (86.1) | 5,014 (87.3) | 468 (80.0) | 129 (73.3) | 8,043 (91.2) | 7,008 (92.1) | 793 (84.1) | 110 (92.4) | 7,841 (93.1) | 6,553 (93.6) | 1,045 (90.6) | 100 (90.1) |
| *Missing* | 386 (5.8) | 301 (5.2) | 38 (6.5) | 31 (17.6) | 209 (2.4) | 170 (2.2) | 29 (3.1) | 3 (2.5) | 188 (2.2) | 151 (2.2) | 33 (2.9) | 2 (1.8) |
| Educational attainment [ǂ h] | | | | | | | | | | | | |
| <High school or GED | 1,600 (24.2) | 1,362 (23.7) | 158 (27.0) | 51 (29.0) | 1,140 (12.9) | 957 (12.6) | 143 (15.2) | 18 (15.1) | 465 (5.5) | 365 (5.2) | 83 (7.2) | 10 (9.0) |
| Some college / technical | 2,678 (40.5) | 2,318 (40.4) | 242 (41.4) | 71 (40.3) | 2,986 (33.9) | 2,509 (33.0) | 370 (39.2) | 47 (39.5) | 1,939 (23.0) | 1,567 (22.4) | 301 (26.1) | 32 (28.8) |
| ≥ Bachelor's or higher | 2,334 (35.3) | 2,059 (35.9) | 185 (31.6) | 54 (30.7) | 4,694 (53.2) | 4,139 (54.4) | 430 (45.6) | 54 (45.4) | 6,014 (71.4) | 5,069 (72.4) | 770 (66.7) | 69 (62.2) |
| *Missing* | 2 (0.0) | 2 (0.0) | 0 (0.0) | 0 (0.0) | 0 (0.0) | 0 (0.0) | 0 (0.0) | 0 (0.0) | 1 (0.0) | 0 (0.0) | 0 (0.0) | 0 (0.0) |
| Marital status [ǂ] | | | | | | | | | | | | |
| Married or living as though married | 2,531 (38.3) | 2,141 (37.3) | 278 (47.5) | 67 (38.1) | 6,221 (70.5) | 5,279 (69.4) | 735 (77.9) | 87 (73.1) | 7,372 (87.6) | 6,088 (87.0) | 1,052 (91.2) | 97 (87.4) |
| Never married, divorced, widowed, or separated | 4,083 (61.7) | 3,600 (62.7) | 307 (52.5) | 109 (61.9) | 2,599 (29.5) | 2,326 (30.6) | 208 (22.1) | 32 (26.9) | 1,047 (12.4) | 913 (13.0) | 102 (8.8) | 14 (12.6) |
| Region of residence [ǂ] | | | | | | | | | | | | |
| Northeast | 905 (13.7) | 797 (13.9) | 81 (13.8) | 18 (10.2) | 1,343 (15.2) | 1,192 (15.7) | 113 (12.0) | 20 (16.8) | 1,628 (19.3) | 1,345 (19.2) | 236 (20.5) | 17 (15.3) |
| Midwest | 1,893 (28.6) | 1,667 (29.0) | 148 (25.3) | 50 (28.4) | 2,475 (28.1) | 2,124 (27.9) | 276 (29.3) | 27 (22.7) | 1,922 (22.8) | 1,584 (22.6) | 278 (24.1) | 24 (21.6) |
| South | 2,184 (33.0) | 1,882 (32.8) | 212 (36.2) | 48 (27.3) | 2,916 (33.1) | 2,486 (32.7) | 336 (35.6) | 41 (34.5) | 2,816 (33.4) | 2,354 (33.6) | 373 (32.3) | 42 (37.8) |
| West | 1,433 (21.7) | 1,247 (21.7) | 125 (21.4) | 38 (21.6) | 2,046 (23.2) | 1,768 (23.2) | 215 (22.8) | 30 (25.2) | 2,043 (24.3) | 1,710 (24.4) | 265 (23.0) | 28 (25.2) |
| Puerto Rico | 199 (3.0) | 148 (2.6) | 19 (3.2) | 22 (12.5) | 40 (0.5) | 35 (0.5) | 3 (0.3) | 1 (0.8) | 10 (0.1) | 8 (0.1) | 2 (0.2) | 0 (0.0) |
| **Clinical characteristics** | | | | | | | | | | | | |
| Body Mass Index, $kg/m^2$ [ǂ] | | | | | | | | | | | | |
| Recommended (18.5 - 24.9) | 2,076 (31.4) | 1,844 (32.1) | 148 (25.3) | 59 (33.5) | 3,294 (37.3) | 2,884 (37.9) | 299 (31.7) | 54 (45.4) | 3,722 (44.2) | 3,163 (45.2) | 430 (37.3) | 53 (47.7) |
| Overweight (25.0 - 29.9) | 2,099 (31.7) | 1,827 (31.8) | 187 (32.0) | 48 (27.3) | 2,877 (32.6) | 2,498 (32.8) | 313 (33.2) | 23 (19.3) | 2,657 (31.6) | 2,182 (31.2) | 395 (34.2) | 30 (27.0) |
| Obesity (≥ 30) | 2,343 (35.4) | 1,984 (34.6) | 245 (41.9) | 66 (37.5) | 2,536 (28.8) | 2,130 (28.0) | 317 (33.6) | 41 (34.5) | 1,944 (23.1) | 1,577 (22.5) | 316 (27.4) | 26 (23.4) |
| *Missing* | 96 (1.5) | 86 (1.5) | 5 (0.9) | 3 (1.7) | 113 (1.3) | 93 (1.2) | 14 (1.5) | 1 (0.8) | 96 (1.1) | 79 (1.1) | 13 (1.1) | 2 (1.8) |
| Menopausal status [ǂ] | | | | | | | | | | | | |
| Premenopausal | 91 (1.4) | 67 (1.2) | 18 (3.1) | 1 (0.6) | 209 (2.4) | 144 (1.9) | 48 (5.1) | 9 (7.6) | 491 (5.8) | 377 (5.4) | 92 (8.0) | 10 (9.0) |
| Postmenopausal | 6,520 (98.6) | 5,671 (98.8) | 567 (96.9) | 175 (99.4) | 8,610 (97.6) | 7,460 (98.1) | 895 (94.9) | 110 (92.4) | 7,927 (94.2) | 6,623 (94.6) | 1,062 (92.0) | 101 (91.0) |

| | | | | | | | | | | | | |
|---|---|---|---|---|---|---|---|---|---|---|---|---|
| *Missing or never had a period* | 3 (0.0) | 3 (0.1) | 0 (0.0) | 0 (0.0) | 1 (0.0) | 1 (0.0) | 0 (0.0) | 0 (0.0) | 1 (0.0) | 1 (0.0) | 0 (0.0) | 0 (0.0) |
| **Sleep health dimensions** | | | | | | | | | | | | |
| Average weekly sleep duration [ƚ i] | | | | | | | | | | | | |
| Short | 932 (14.1) | 792 (13.8) | 85 (14.5) | 33 (18.8) | 1,027 (11.6) | 840 (11.0) | 147 (15.6) | 19 (16.0) | 885 (10.5) | 716 (10.2) | 137 (11.9) | 15 (13.5) |
| Recommended | 4,255 (64.3) | 3,748 (65.3) | 362 (61.9) | 94 (53.4) | 6,327 (71.7) | 5,510 (72.5) | 638 (67.7) | 81 (68.1) | 6,316 (75.0) | 5,309 (75.8) | 825 (71.5) | 67 (60.4) |
| Long | 1,280 (19.4) | 1,084 (18.9) | 125 (21.4) | 38 (21.6) | 1,384 (15.7) | 1,182 (15.5) | 151 (16.0) | 18 (15.1) | 1,155 (13.7) | 922 (13.2) | 185 (16.0) | 27 (24.3) |
| *Missing* | 147 (2.2) | 117 (2.0) | 13 (2.2) | 11 (6.3) | 82 (0.9) | 73 (1.0) | 7 (0.7) | 1 (0.8) | 63 (0.7) | 54 (0.8) | 7 (0.6) | 2 (1.8) |
| Long sleep onset latency [ƚ j] | | | | | | | | | | | | |
| Yes | 1,420 (21.5) | 1,106 (19.3) | 192 (32.8) | 64 (36.4) | 1,561 (17.7) | 1,186 (15.6) | 274 (29.1) | 31 (26.1) | 1,202 (14.3) | 905 (12.9) | 223 (19.3) | 25 (22.5) |
| No | 5,099 (77.1) | 4,559 (79.4) | 382 (65.3) | 107 (60.8) | 7,196 (81.6) | 6,361 (83.6) | 666 (70.6) | 88 (73.9) | 7,167 (85.1) | 6,057 (86.5) | 925 (80.2) | 83 (74.8) |
| *Missing* | 95 (1.4) | 76 (1.3) | 11 (1.9) | 5 (2.8) | 63 (0.7) | 58 (0.8) | 3 (0.3) | 0 (0.0) | 50 (0.6) | 39 (0.6) | 6 (0.5) | 3 (2.7) |
| Poor sleep maintenance [ƚ k] | | | | | | | | | | | | |
| Yes | 3,634 (54.9) | 3,011 (52.4) | 426 (72.8) | 112 (63.6) | 4,832 (54.8) | 3,957 (52.0) | 677 (71.8) | 82 (68.9) | 4,794 (56.9) | 3,744 (53.5) | 861 (74.6) | 68 (61.3) |
| No | 2,900 (43.8) | 2,657 (46.3) | 157 (26.8) | 62 (35.2) | 3,926 (44.5) | 3,592 (47.2) | 262 (27.8) | 37 (31.1) | 3,577 (42.5) | 3,216 (45.9) | 286 (24.8) | 43 (38.7) |
| *Missing* | 80 (1.2) | 73 (1.3) | 2 (0.3) | 2 (1.1) | 62 (0.7) | 56 (0.7) | 4 (0.4) | 0 (0.0) | 48 (0.6) | 41 (0.6) | 7 (0.6) | 0 (0.0) |
| Insomnia symptoms [l] | | | | | | | | | | | | |
| Yes | 3,961 (59.9) | 3,288 (57.3) | 458 (78.3) | 120 (68.2) | 5,263 (59.7) | 4,316 (56.8) | 731 (77.5) | 85 (71.4) | 5,131 (60.9) | 4,017 (57.4) | 913 (79.1) | 74 (66.7) |
| No | 2,619 (39.6) | 2,425 (42.2) | 126 (21.5) | 54 (30.7) | 3,538 (40.1) | 3,270 (43.0) | 212 (22.5) | 34 (28.6) | 3,274 (38.9) | 2,972 (42.5) | 239 (20.7) | 37 (33.3) |
| *Missing* | 34 (0.5) | 28 (0.5) | 1 (0.2) | 2 (1.1) | 19 (0.2) | 19 (0.2) | 0 (0.0) | 0 (0.0) | 14 (0.2) | 12 (0.2) | 2 (0.2) | 0 (0.0) |
| Sleep medication use [ƚ m] | | | | | | | | | | | | |
| Yes | 1,531 (23.1) | 1,276 (22.2) | 160 (27.4) | 55 (31.3) | 1,896 (21.5) | 1,595 (21.0) | 229 (24.3) | 25 (21.0) | 1,650 (19.6) | 1,348 (19.3) | 234 (20.3) | 34 (30.6) |
| No | 5,038 (76.2) | 4,430 (77.2) | 422 (72.1) | 118 (67.0) | 6,893 (78.2) | 5,986 (78.7) | 708 (75.1) | 93 (78.2) | 6,743 (80.1) | 5,631 (80.4) | 917 (79.5) | 77 (69.4) |
| *Missing* | 45 (0.7) | 35 (0.6) | 3 (0.5) | 3 (1.7) | 31 (0.4) | 24 (0.3) | 6 (0.6) | 1 (0.8) | 26 (0.3) | 22 (0.3) | 3 (0.3) | 0 (0.0) |
| Daytime dysfunction [ƚ n] | | | | | | | | | | | | |
| Yes | 82 (1.2) | 63 (1.1) | 10 (1.7) | 5 (2.8) | 71 (0.8) | 48 (0.6) | 13 (1.4) | 7 (5.9) | 66 (0.8) | 44 (0.6) | 13 (1.1) | 4 (3.6) |
| No | 6,447 (97.5) | 5,602 (97.6) | 570 (97.4) | 169 (96.0) | 8,635 (97.9) | 7,460 (98.1) | 915 (97.0) | 111 (93.3) | 8,274 (98.3) | 6,891 (98.4) | 1,130 (97.9) | 106 (95.5) |
| *Missing* | 85 (1.3) | 76 (1.3) | 5 (0.9) | 2 (1.1) | 114 (1.3) | 97 (1.3) | 15 (1.6) | 1 (0.8) | 79 (0.9) | 66 (0.9) | 11 (1.0) | 1 (0.9) |
| Healthcare provider diagnosed sleep apnea [ƚ o] | | | | | | | | | | | | |
| Yes | 879 (13.3) | 737 (12.8) | 94 (16.1) | 33 (18.8) | 942 (10.7) | 780 (10.3) | 113 (12.0) | 22 (18.5) | 743 (8.8) | 600 (8.6) | 118 (10.2) | 16 (14.4) |
| No | 5,615 (84.9) | 4,906 (85.5) | 481 (82.2) | 136 (77.3) | 7,783 (88.2) | 6,740 (88.6) | 826 (87.6) | 93 (78.2) | 7,625 (90.6) | 6,356 (90.8) | 1,031 (89.3) | 95 (85.6) |
| *Missing* | 120 (1.8) | 98 (1.7) | 10 (1.7) | 7 (4.0) | 95 (1.1) | 85 (1.1) | 4 (0.4) | 4 (3.4) | 51 (0.6) | 45 (0.6) | 5 (0.4) | 0 (0.0) |

Abbreviations: SD, Standard Deviation; GED: General Educational Development

Note: N = 418 (1.8%) participants who reported both 'too hot' and 'too cold' temperatures were not included in this sensitivity analysis.

[¥] Chi-square test *p*-values indicate significant differences between the overall prevalence of 'too hot' by annual household income.

[§] Chi-square test *p*-values indicate significant differences between the overall prevalence of 'too cold' by annual household income.

[ƚ] Chi-square or Analysis of Variance (ANOVA) test *p*-values indicate significant differences in characteristics between the total populations of each annual household income category.

[a] Reasons for trouble sleeping ≥ 3 times per week in the past month include: unable to fall asleep within 30 minutes, waking up in the middle of the night or early morning, waking up to use the bathroom, cannot breathe comfortably, coughing or snoring loudly, having bad dreams, having pain, or other non-specified reasons.

[b] Excluded for reporting both 'Too hot' and 'Too cold', < $20,000 - $49,999: n = 112 (1.7%).

[c] Excluded for reporting both 'Too hot' and 'Too cold', $50,000 - $99,999: n = 153 (1.7%).

[d] Excluded for reporting both 'Too hot' and 'Too cold', ≥ $100,000: n = 153 (1.8%).

[e] 'Infrequent temperature extremes' is defined as self-reported trouble sleeping due to feeling too hot < 3 times per week and trouble sleeping due to feeling too cold < 3 times per week in the past month.

[f] 'Too hot' is defined as self-reported trouble sleeping due to feeling too hot ≥ 3 times per week in the past month.

[g] 'Too cold' is defined as self-reported trouble sleeping due to feeling too cold ≥ 3 times per week in the past month.

[h] Educational attainment was assessed at baseline.
[i] Sleep duration is based on reported bed and wake times or reported average sleep duration. Participants who reported ≤ 2 or ≥ 23 hours of sleep are excluded. Short: < 7 hours; Recommended: 7-9 hours; Long ≥ 9 hours.
[j] Long sleep onset latency is defined as not falling asleep within 30 minutes at least three times a week during the past month.
[k] Poor sleep maintenance is defined as waking up in the middle of the night or early morning at least three times a week during the past month.
[l] Insomnia symptoms is defined as long sleep onset latency or poor sleep maintenance.
[m] Sleep medication use is defined as 'taking medicine (prescription or over the counter) to help you sleep' at least three times a week during the past month.
[n] Daytime dysfunction is defined as having trouble staying awake while driving, eating, or engaging in social activity at least three times a week during the past month.
[o] Healthcare provider diagnosed sleep apnea is defined as a current doctor or other health professional diagnosis of sleep apnea.

**Supplemental Table 14. Sociodemographic, clinical, and sleep health characteristics among participants who reported trouble sleeping for any reason [a], by region of residence as well as perceived indoor temperature extremes, Sister Study, 2017-2019, N = 23,853**

| Region | Northeast [b] | | | | Midwest [c] | | | | South [d] | | | | West [e] | | | | Puerto Rico [f] | | | |
|---|---|---|---|---|---|---|---|---|---|---|---|---|---|---|---|---|---|---|---|---|
| | Total n = 3,876 (16.2%) | Infrequent temperature extremes[g] n = 3,334 (86.0%) | Too hot[h ¥] n = 430 (11.1%) | Too cold[i §] n = 55 (1.4%) | Total n = 6,290 (26.4%) | Infrequent temperature extremes[g] n = 5,375 (85.5%) | Too hot[h ¥] n = 702 (11.2%) | Too cold[i §] n = 101 (1.6%) | Total n = 7,916 (33.2%) | Infrequent temperature extremes[g] n = 6,722 (84.9%) | Too hot[h ¥] n = 921 (11.6%) | Too cold[i §] n = 131 (1.7%) | Total n = 5,522 (23.2%) | Infrequent temperature extremes[g] n = 4,725 (85.6%) | Too hot[h ¥] n = 605 (11.0%) | Too cold[i §] n = 96 (1.7%) | Total n = 249 (1.0%) | Infrequent temperature extremes[g] n = 191 (76.7%) | Too hot[h ¥] n = 24 (9.6%) | Too cold[i §] n =23 (9.2%) |
| **Sociodemographic characteristics** | | | | | | | | | | | | | | | | | | | | |
| Age, years (mean ± SD) [ǂ] | 67.23 ± 8.58 | 68.00 ± 8.45 | 61.77 ± 7.41 | 68.83 ± 8.31 | 67.41 ± 8.47 | 68.16 ± 8.32 | 62.24 ± 7.48 | 68.08 ± 9.13 | 67.61 ± 8.44 | 68.31 ± 8.31 | 63.01 ± 7.69 | 68.64 ± 9.74 | 68.17 ± 8.35 | 68.78 ± 8.22 | 63.71 ± 7.84 | 70.80 ± 8.15 | 66.16 ± 8.01 | 66.44 ± 7.97 | 64.12 ± 7.63 | 66.23 ± 8.97 |
| Age, [ǂ] years | | | | | | | | | | | | | | | | | | | | |
| < 67.2 | 1,940 (50.1) | 1,538 (46.1) | 336 (78.1) | 20 (36.4) | 3,068 (48.8) | 2,414 (44.9) | 527 (75.1) | 44 (43.6) | 3,764 (47.5) | 2,963 (44.1) | 640 (69.5) | 57 (43.5) | 2,504 (45.3) | 2,010 (42.5) | 401 (66.3) | 29 (30.2) | 135 (54.2) | 100 (52.4) | 16 (66.7) | 14 (60.9) |
| ≥ 67.2 | 1,936 (49.9) | 1,796 (53.9) | 94 (21.9) | 35 (63.6) | 3,222 (51.2) | 2,961 (55.1) | 175 (24.9) | 57 (56.4) | 4,152 (52.5) | 3,759 (55.9) | 281 (30.5) | 74 (56.5) | 3,018 (54.7) | 2,715 (57.5) | 204 (33.7) | 67 (69.8) | 114 (45.8) | 91 (47.6) | 8 (33.3) | 9 (39.1) |
| Race and ethnicity [ǂ] | | | | | | | | | | | | | | | | | | | | |
| Hispanic/Latina | 63 (1.6) | 45 (1.3) | 14 (3.3) | 1 (1.8) | 47 (0.7) | 41 (0.8) | 6 (0.9) | 0 (0.0) | 212 (2.7) | 172 (2.6) | 28 (3.0) | 7 (5.3) | 215 (3.9) | 176 (3.7) | 28 (4.6) | 5 (5.2) | 246 (98.8) | 188 (98.4) | 24 (100.0) | 23 (100.0) |
| Non-Hispanic Black | 129 (3.3) | 102 (3.1) | 22 (5.1) | 2 (3.6) | 284 (4.5) | 223 (4.1) | 50 (7.1) | 6 (5.9) | 953 (12.0) | 718 (10.7) | 189 (20.5) | 18 (13.7) | 128 (2.3) | 107 (2.3) | 15 (2.5) | 5 (5.2) | 0 (0.0) | 0 (0.0) | 0 (0.0) | 0 (0.0) |
| Non-Hispanic White | 3,684 (95.0) | 3,187 (95.6) | 394 (91.6) | 52 (94.5) | 5,959 (94.7) | 5,111 (95.1) | 646 (92.0) | 95 (94.1) | 6,751 (85.3) | 5,832 (86.8) | 704 (76.4) | 106 (80.9) | 5,179 (93.8) | 4,442 (94.0) | 562 (92.9) | 86 (89.6) | 3 (1.2) | 3 (1.6) | 0 (0.0) | 0 (0.0) |
| Educational attainment [ǂ j] | | | | | | | | | | | | | | | | | | | | |
| <High school or GED | 513 (13.2) | 432 (13.0) | 59 (13.7) | 15 (27.3) | 1,046 (16.6) | 880 (16.4) | 129 (18.4) | 23 (22.8) | 1,021 (12.9) | 866 (12.9) | 113 (12.3) | 26 (19.8) | 572 (10.4) | 469 (9.9) | 75 (12.4) | 11 (11.5) | 53 (21.3) | 37 (19.4) | 8 (33.3) | 4 (17.4) |
| Some college / Technical | 1,082 (27.9) | 910 (27.3) | 133 (30.9) | 19 (34.5) | 2,125 (33.8) | 1,808 (33.6) | 233 (33.2) | 42 (41.6) | 2,526 (31.9) | 2,111 (31.4) | 320 (34.7) | 46 (35.1) | 1,803 (32.7) | 1,517 (32.1) | 220 (36.4) | 36 (37.5) | 67 (26.9) | 48 (25.1) | 7 (29.2) | 7 (30.4) |
| ≥ Bachelor's or Higher | 2,279 (58.8) | 1,991 (59.7) | 238 (55.3) | 21 (38.2) | 3,119 (49.6) | 2,687 (50.0) | 340 (48.4) | 36 (35.6) | 4,368 (55.2) | 3,744 (55.7) | 488 (53.0) | 59 (45.0) | 3,147 (57.0) | 2,739 (58.0) | 310 (51.2) | 49 (51.0) | 129 (51.8) | 106 (55.5) | 9 (37.5) | 12 (52.2) |
| *Missing* | *2 (0.1)* | *1 (0.0)* | *0 (0.0)* | *0 (0.0)* | *0 (0.0)* | *0 (0.0)* | *0 (0.0)* | *0 (0.0)* | *1 (0.0)* | *1 (0.0)* | *0 (0.0)* | *0 (0.0)* | *0 (0.0)* | *0 (0.0)* | *0 (0.0)* | *0 (0.0)* | *0 (0.0)* | *0 (0.0)* | *0 (0.0)* | *0 (0.0)* |
| Annual household income [ǂ] | | | | | | | | | | | | | | | | | | | | |
| < $20,000 - $49,999 | 905 (23.3) | 797 (23.9) | 81 (18.8) | 18 (32.7) | 1,893 (30.1) | 1,667 (31.0) | 148 (21.1) | 50 (49.5) | 2,184 (27.6) | 1,882 (28.0) | 212 (23.0) | 48 (36.6) | 1,433 (26.0) | 1,247 (26.4) | 125 (20.7) | 38 (39.6) | 199 (79.9) | 148 (77.5) | 19 (79.2) | 22 (95.7) |
| $50,000 – $99,999 | 1,343 (34.6) | 1,192 (35.8) | 113 (26.3) | 20 (36.4) | 2,475 (39.3) | 2,124 (39.5) | 276 (39.3) | 27 (26.7) | 2,916 (36.8) | 2,486 (37.0) | 336 (36.5) | 41 (31.3) | 2,046 (37.1) | 1,768 (37.4) | 215 (35.5) | 30 (31.3) | 40 (16.1) | 35 (18.3) | 3 (12.5) | 1 (4.3) |
| ≥ $100,000 | 1,628 (42.0) | 1,345 (40.3) | 236 (54.9) | 17 (30.9) | 1,922 (30.6) | 1,584 (29.5) | 278 (39.6) | 24 (23.8) | 2,816 (35.6) | 2,354 (35.0) | 373 (40.5) | 42 (32.1) | 2,043 (37.0) | 1,710 (36.2) | 265 (43.8) | 28 (29.2) | 10 (4.0) | 8 (4.2) | 2 (8.3) | 0 (0.0) |
| Marital status [ǂ] | | | | | | | | | | | | | | | | | | | | |
| Married or living as though married | 2,643 (68.2) | 2,223 (66.7) | 339 (78.8) | 39 (70.9) | 4,341 (69.0) | 3,642 (67.8) | 553 (78.8) | 65 (64.4) | 5,305 (67.0) | 4,443 (66.1) | 687 (74.6) | 80 (61.1) | 3,676 (66.6) | 3,077 (65.1) | 469 (77.5) | 56 (58.3) | 159 (63.9) | 123 (64.4) | 17 (70.8) | 11 (47.8) |
| Never married, divorced, widowed, or separated | 1,233 (31.8) | 1,111 (33.3) | 91 (21.2) | 16 (29.1) | 1,949 (31.0) | 1,733 (32.2) | 149 (21.2) | 36 (35.6) | 2,611 (33.0) | 2,279 (33.9) | 234 (25.4) | 51 (38.9) | 1,846 (33.4) | 1,648 (34.9) | 136 (22.5) | 40 (41.7) | 90 (36.1) | 68 (35.6) | 7 (29.2) | 12 (52.2) |

| | | | | | | | | | | | | | | | | | | | | |
|---|---|---|---|---|---|---|---|---|---|---|---|---|---|---|---|---|---|---|---|---|
| **Clinical characteristics** | | | | | | | | | | | | | | | | | | | | |
| Body Mass Index, kg/m$^2$ [‡] | | | | | | | | | | | | | | | | | | | | |
| Recommended (18.5 - 24.9) | 1,571 (40.5) | 1,377 (41.3) | 146 (34.0) | 21 (38.2) | 2,152 (34.2) | 1,854 (34.5) | 221 (31.5) | 38 (37.6) | 2,882 (36.4) | 2,530 (37.6) | 260 (28.2) | 47 (35.9) | 2,416 (43.8) | 2,080 (44.0) | 240 (39.7) | 53 (55.2) | 71 (28.5) | 50 (26.2) | 10 (41.7) | 7 (30.4) |
| Overweight (25.0 - 29.9) | 1,216 (31.4) | 1,048 (31.4) | 138 (32.1) | 12 (21.8) | 2,068 (32.9) | 1,764 (32.8) | 237 (33.8) | 30 (29.7) | 2,548 (32.2) | 2,161 (32.1) | 309 (33.6) | 31 (23.7) | 1,698 (30.7) | 1,452 (30.7) | 203 (33.6) | 19 (19.8) | 103 (41.4) | 82 (42.9) | 8 (33.3) | 9 (39.1) |
| Obesity (≥ 30) | 1,041 (26.9) | 868 (26.0) | 141 (32.8) | 21 (38.2) | 1,998 (31.8) | 1,696 (31.6) | 233 (33.2) | 33 (32.7) | 2,372 (30.0) | 1,934 (28.8) | 343 (37.2) | 50 (38.2) | 1,340 (24.3) | 1,136 (24.0) | 155 (25.6) | 22 (22.9) | 72 (28.9) | 57 (29.8) | 6 (25.0) | 7 (30.4) |
| *Missing* | *48 (1.2)* | *41 (1.2)* | *5 (1.2)* | *1 (1.8)* | *72 (1.1)* | *61 (1.1)* | *11 (1.6)* | *0 (0.0)* | *114 (1.4)* | *97 (1.4)* | *9 (1.0)* | *3 (2.3)* | *68 (1.2)* | *57 (1.2)* | *7 (1.2)* | *2 (2.1)* | *3 (1.2)* | *2 (1.0)* | *0 (0.0)* | *0 (0.0)* |
| Menopausal status | | | | | | | | | | | | | | | | | | | | |
| Premenopausal | 131 (3.4) | 98 (2.9) | 26 (6.0) | 2 (3.6) | 210 (3.3) | 157 (2.9) | 42 (6.0) | 6 (5.9) | 270 (3.4) | 197 (2.9) | 56 (6.1) | 8 (6.1) | 166 (3.0) | 124 (2.6) | 33 (5.5) | 3 (3.1) | 14 (5.6) | 12 (6.3) | 1 (4.2) | 1 (4.3) |
| Postmenopausal | 3,745 (96.6) | 3,236 (97.1) | 404 (94.0) | 53 (96.4) | 6,080 (96.7) | 5,218 (97.1) | 660 (94.0) | 95 (94.1) | 7,642 (96.5) | 6,521 (97.0) | 865 (93.9) | 123 (93.9) | 5,355 (97.0) | 4,600 (97.4) | 572 (94.5) | 93 (96.9) | 235 (94.4) | 179 (93.7) | 23 (95.8) | 22 (95.7) |
| *Missing or never had a period* | *0 (0.0)* | *0 (0.0)* | *0 (0.0)* | *0 (0.0)* | *0 (0.0)* | *0 (0.0)* | *0 (0.0)* | *0 (0.0)* | *4 (0.1)* | *4 (0.1)* | *0 (0.0)* | *0 (0.0)* | *1 (0.0)* | *1 (0.0)* | *0 (0.0)* | *0 (0.0)* | *0 (0.0)* | *0 (0.0)* | *0 (0.0)* | *0 (0.0)* |
| **Sleep health dimensions** | | | | | | | | | | | | | | | | | | | | |
| Average weekly sleep duration [‡ k] | | | | | | | | | | | | | | | | | | | | |
| Short | 522 (13.5) | 442 (13.3) | 58 (13.5) | 15 (27.3) | 704 (11.2) | 564 (10.5) | 108 (15.4) | 17 (16.8) | 1,004 (12.7) | 824 (12.3) | 129 (14.0) | 22 (16.8) | 572 (10.4) | 486 (10.3) | 68 (11.2) | 11 (11.5) | 42 (16.9) | 32 (16.8) | 6 (25.0) | 2 (8.7) |
| Recommended | 2,715 (70.0) | 2,344 (70.3) | 303 (70.5) | 29 (52.7) | 4,539 (72.2) | 3,930 (73.1) | 477 (67.9) | 66 (65.3) | 5,502 (69.5) | 4,719 (70.2) | 621 (67.4) | 77 (58.8) | 3,986 (72.2) | 3,446 (72.9) | 416 (68.8) | 55 (57.3) | 156 (62.7) | 128 (67.0) | 8 (33.3) | 15 (65.2) |
| Long | 584 (15.1) | 498 (14.9) | 66 (15.3) | 9 (16.4) | 972 (15.5) | 815 (15.2) | 112 (16.0) | 15 (14.9) | 1,316 (16.6) | 1,104 (16.4) | 160 (17.4) | 27 (20.6) | 908 (16.4) | 747 (15.8) | 116 (19.2) | 28 (29.2) | 39 (15.7) | 24 (12.6) | 7 (29.2) | 4 (17.4) |
| *Missing* | *55 (1.4)* | *50 (1.5)* | *3 (0.7)* | *2 (3.6)* | *75 (1.2)* | *66 (1.2)* | *5 (0.7)* | *3 (3.0)* | *94 (1.2)* | *75 (1.1)* | *11 (1.2)* | *5 (3.8)* | *56 (1.0)* | *46 (1.0)* | *5 (0.8)* | *2 (2.1)* | *12 (4.8)* | *7 (3.7)* | *3 (12.5)* | *2 (8.7)* |
| Long sleep onset latency [‡ l] | | | | | | | | | | | | | | | | | | | | |
| Yes | 634 (16.4) | 499 (15.0) | 102 (23.7) | 13 (23.6) | 1,064 (16.9) | 822 (15.3) | 161 (22.9) | 34 (33.7) | 1,501 (19.0) | 1,126 (16.8) | 278 (30.2) | 34 (26.0) | 910 (16.5) | 700 (14.8) | 141 (23.3) | 28 (29.2) | 74 (29.7) | 50 (26.2) | 7 (29.2) | 11 (47.8) |
| No | 3,211 (82.8) | 2,809 (84.3) | 324 (75.3) | 41 (74.5) | 5,177 (82.3) | 4,511 (83.9) | 537 (76.5) | 66 (65.3) | 6,344 (80.1) | 5,535 (82.3) | 638 (69.3) | 95 (72.5) | 4,571 (82.8) | 3,989 (84.4) | 461 (76.2) | 67 (69.8) | 159 (63.9) | 133 (69.6) | 13 (54.2) | 9 (39.1) |
| *Missing* | *31 (0.8)* | *26 (0.8)* | *4 (0.9)* | *1 (1.8)* | *49 (0.8)* | *42 (0.8)* | *4 (0.6)* | *1 (1.0)* | *71 (0.9)* | *61 (0.9)* | *5 (0.5)* | *2 (1.5)* | *41 (0.7)* | *36 (0.8)* | *3 (0.5)* | *1 (1.0)* | *16 (6.4)* | *8 (4.2)* | *4 (16.7)* | *3 (13.0)* |
| Poor sleep maintenance [‡ m] | | | | | | | | | | | | | | | | | | | | |
| Yes | 2,228 (57.5) | 1,808 (54.2) | 339 (78.8) | 33 (60.0) | 3,437 (54.6) | 2,781 (51.7) | 501 (71.4) | 70 (69.3) | 4,336 (54.8) | 3,477 (51.7) | 661 (71.8) | 85 (64.9) | 3,141 (56.9) | 2,565 (54.3) | 447 (73.9) | 59 (61.5) | 118 (47.4) | 81 (42.4) | 16 (66.7) | 15 (65.2) |
| No | 1,620 (41.8) | 1,499 (45.0) | 90 (20.9) | 22 (40.0) | 2,809 (44.7) | 2,557 (47.6) | 196 (27.9) | 31 (30.7) | 3,515 (44.4) | 3,184 (47.4) | 257 (27.9) | 46 (35.1) | 2,341 (42.4) | 2,124 (45.0) | 155 (25.6) | 37 (38.5) | 118 (47.4) | 101 (52.9) | 7 (29.2) | 6 (26.1) |
| *Missing* | *28 (0.7)* | *27 (0.8)* | *1 (0.2)* | *0 (0.0)* | *44 (0.7)* | *37 (0.7)* | *5 (0.7)* | *0 (0.0)* | *65 (0.8)* | *61 (0.9)* | *3 (0.3)* | *0 (0.0)* | *40 (0.7)* | *36 (0.8)* | *3 (0.5)* | *0 (0.0)* | *13 (5.2)* | *9 (4.7)* | *1 (4.2)* | *2 (8.7)* |
| Insomnia symptoms [‡ n] | | | | | | | | | | | | | | | | | | | | |
| Yes | 2,380 (61.4) | 1,939 (58.2) | 356 (82.8) | 34 (61.8) | 3,739 (59.4) | 3,035 (56.5) | 536 (76.4) | 74 (73.3) | 4,742 (59.9) | 3,814 (56.7) | 712 (77.3) | 92 (70.2) | 3,357 (60.8) | 2,737 (57.9) | 480 (79.3) | 62 (64.6) | 137 (55.0) | 96 (50.3) | 18 (75.0) | 17 (73.9) |
| No | 1,489 (38.4) | 1,388 (41.6) | 74 (17.2) | 21 (38.2) | 2,536 (40.3) | 2,328 (43.3) | 164 (23.4) | 27 (26.7) | 3,154 (39.8) | 2,889 (43.0) | 209 (22.7) | 39 (29.8) | 2,151 (39.0) | 1,974 (41.8) | 125 (20.7) | 34 (35.4) | 101 (40.6) | 88 (46.1) | 5 (20.8) | 4 (17.4) |
| *Missing* | *7 (0.2)* | *7 (0.2)* | *0 (0.0)* | *0 (0.0)* | *15 (0.2)* | *12 (0.2)* | *2 (0.3)* | *0 (0.0)* | *20 (0.3)* | *19 (0.3)* | *0 (0.0)* | *0 (0.0)* | *14 (0.3)* | *14 (0.3)* | *0 (0.0)* | *0 (0.0)* | *11 (4.4)* | *7 (3.7)* | *1 (4.2)* | *2 (8.7)* |

| | | | | | | | | | | | | | | | | | | | | |
|---|---|---|---|---|---|---|---|---|---|---|---|---|---|---|---|---|---|---|---|---|
| Sleep medication use [ǀ] [o] | | | | | | | | | | | | | | | | | | | | |
| Yes | 732 (18.9) | 608 (18.2) | 94 (21.9) | 18 (32.7) | 1,297 (20.6) | 1,085 (20.2) | 148 (21.1) | 31 (30.7) | 1,744 (22.0) | 1,435 (21.3) | 231 (25.1) | 33 (25.2) | 1,239 (22.4) | 1,047 (22.2) | 143 (23.6) | 22 (22.9) | 65 (26.1) | 44 (23.0) | 7 (29.2) | 10 (43.5) |
| No | 3,129 (80.7) | 2,715 (81.4) | 333 (77.4) | 37 (67.3) | 4,968 (79.0) | 4,268 (79.4) | 551 (78.5) | 70 (69.3) | 6,139 (77.6) | 5,262 (78.3) | 685 (74.4) | 96 (73.3) | 4,261 (77.2) | 3,659 (77.4) | 461 (76.2) | 73 (76.0) | 177 (71.1) | 143 (74.9) | 17 (70.8) | 12 (52.2) |
| *Missing* | *15 (0.4)* | *11 (0.3)* | *3 (0.7)* | *0 (0.0)* | *25 (0.4)* | *22 (0.4)* | *3 (0.4)* | *0 (0.0)* | *33 (0.4)* | *25 (0.4)* | *5 (0.5)* | *2 (1.5)* | *22 (0.4)* | *19 (0.4)* | *1 (0.2)* | *1 (1.0)* | *7 (2.8)* | *4 (2.1)* | *0 (0.0)* | *1 (4.3)* |
| Daytime dysfunction [ǀ] [p] | | | | | | | | | | | | | | | | | | | | |
| Yes | 31 (0.8) | 24 (0.7) | 5 (1.2) | 2 (3.6) | 53 (0.8) | 43 (0.8) | 7 (1.0) | 1 (1.0) | 87 (1.1) | 55 (0.8) | 16 (1.7) | 8 (6.1) | 46 (0.8) | 33 (0.7) | 7 (1.2) | 4 (4.2) | 2 (0.8) | 0 (0.0) | 1 (4.2) | 1 (4.3) |
| No | 3,801 (98.1) | 3,270 (98.1) | 421 (97.9) | 53 (96.4) | 6,160 (97.9) | 5,268 (98.0) | 684 (97.4) | 99 (98.0) | 7,744 (97.8) | 6,596 (98.1) | 893 (97.0) | 122 (93.1) | 5,414 (98.0) | 4,636 (98.1) | 595 (98.3) | 90 (93.8) | 237 (95.2) | 183 (95.8) | 22 (91.7) | 22 (95.7) |
| *Missing* | *44 (1.1)* | *40 (1.2)* | *4 (0.9)* | *0 (0.0)* | *77 (1.2)* | *64 (1.2)* | *11 (1.6)* | *1 (1.0)* | *85 (1.1)* | *71 (1.1)* | *12 (1.3)* | *1 (0.8)* | *62 (1.1)* | *56 (1.2)* | *3 (0.5)* | *2 (2.1)* | *10 (4.0)* | *8 (4.2)* | *1 (4.2)* | *0 (0.0)* |
| Healthcare provider diagnosed sleep apnea [ǀ] [q] | | | | | | | | | | | | | | | | | | | | |
| Yes | 353 (9.1) | 294 (8.8) | 47 (10.9) | 9 (16.4) | 662 (10.5) | 560 (10.4) | 77 (11.0) | 9 (8.9) | 890 (11.2) | 709 (10.5) | 132 (14.3) | 30 (22.9) | 644 (11.7) | 547 (11.6) | 66 (10.9) | 18 (18.8) | 15 (6.0) | 7 (3.7) | 3 (12.5) | 5 (21.7) |
| No | 3,478 (89.7) | 3,000 (90.0) | 379 (88.1) | 45 (81.8) | 5,552 (88.3) | 4,754 (88.4) | 618 (88.0) | 87 (86.1) | 6,939 (87.7) | 5,935 (88.3) | 785 (85.2) | 97 (74.0) | 4,827 (87.4) | 4,134 (87.5) | 535 (88.4) | 77 (80.2) | 227 (91.2) | 179 (93.7) | 21 (87.5) | 18 (78.3) |
| *Missing* | *45 (1.2)* | *40 (1.2)* | *4 (0.9)* | *1 (1.8)* | *76 (1.2)* | *61 (1.1)* | *7 (1.0)* | *5 (5.0)* | *87 (1.1)* | *78 (1.2)* | *4 (0.4)* | *4 (3.1)* | *51 (0.9)* | *44 (0.9)* | *4 (0.7)* | *1 (1.0)* | *7 (2.8)* | *5 (2.6)* | *0 (0.0)* | *0 (0.0)* |

Abbreviations: SD, Standard Deviation; GED: General Educational Development

Note: N = 418 (1.8%) participants who reported both 'too hot' and 'too cold' temperatures were not included in this sensitivity analysis.

[¥] Chi-square test *p*-values indicate significant differences between the overall prevalence of 'too hot' by region of residence.

[§] Chi-square test *p*-values indicate significant differences between the overall prevalence of 'too cold' by region of residence.

[ǀ] Chi-square or Analysis of Variance (ANOVA) test *p*-values indicate significant differences in characteristics between the total populations of each region of residence.

[a] Reasons for trouble sleeping ≥ 3 times per week in the past month include: unable to fall asleep within 30 minutes, waking up in the middle of the night or early morning, waking up to use the bathroom, cannot breathe comfortably, coughing or snoring loudly, having bad dreams, having pain, or other non-specified reasons.

[b] Excluded for reporting both 'Too hot' and 'Too cold', Northeast: n = 57 (1.5%).

[c] Excluded for reporting both 'Too hot' and 'Too cold', Midwest: n = 112 (1.8%).

[d] Excluded for reporting both 'Too hot' and 'Too cold', South: n = 142 (1.8%).

[e] Excluded for reporting both 'Too hot' and 'Too cold', West: n = 96 (1.7%).

[f] Excluded for reporting both 'Too hot' and 'Too cold', Puerto Rico: n = 11 (4.4%).

[g] 'Infrequent temperature extremes' is defined as self-reported trouble sleeping due to feeling too hot < 3 times per week and trouble sleeping due to feeling too cold < 3 times per week in the past month.

[h] 'Too hot' is defined as self-reported trouble sleeping due to feeling too hot ≥ 3 times per week in the past month.

[i] 'Too cold' is defined as self-reported trouble sleeping due to feeling too cold ≥ 3 times per week in the past month.

[j] Educational attainment was assessed at baseline.

[k] Sleep duration is based on reported bed and wake times or reported average sleep duration. Participants who reported ≤ 2 or ≥ 23 hours of sleep are excluded. Short: < 7 hours; Recommended: 7-9 hours; Long ≥ 9 hours.

[l] Long sleep onset latency is defined as not falling asleep within 30 minutes at least three times a week during the past month.

[m] Poor sleep maintenance is defined as waking up in the middle of the night or early morning at least three times a week during the past month.

[n] Insomnia symptoms is defined as long sleep onset latency or poor sleep maintenance.

[o] Sleep medication use is defined as 'taking medicine (prescription or over the counter) to help you sleep' at least three times a week during the past month.

[p] Daytime dysfunction is defined as having trouble staying awake while driving, eating, or engaging in social activity at least three times a week during the past month.

[q] Healthcare provider diagnosed sleep apnea is defined as a current doctor or other health professional diagnosis of sleep apnea.

**Supplemental Table 15. Sociodemographic, clinical, and sleep health characteristics among participants who reported trouble sleeping for any reason [a], by menopausal status as well as perceived indoor temperature extremes, Sister Study, 2017-2019, N = 23,853**

| **Menopausal status** | **Premenopausal [b]** | | | | **Postmenopausal [c]** | | | |
|---|---|---|---|---|---|---|---|---|
| | Total<br>n = 791 (3.3%) | Infrequent temperature extremes [d]<br>n = 588 (74.3%) | Too hot [e ¥]<br>n = 158 (20.0%) | Too cold [f §]<br>n = 20 (2.5%) | Total<br>n = 23,057 (96.7%) | Infrequent temperature extremes [d]<br>n = 19,754 (85.7%) | Too hot [e ¥]<br>n = 2,524 (10.9%) | Too cold [f §]<br>n = 386 (1.7%) |
| **Sociodemographic characteristics** | | | | | | | | |
| Age, years (mean ± SD) [ƚ] | 51.34 ± 2.60 | 51.21 ± 2.60 | 51.74 ± 2.58 | 51.33 ± 2.79 | 68.16 ± 8.02 | 68.82 ± 7.88 | 63.47 ± 7.33 | 69.81 ± 8.28 |
| Age, years (n (%)) [ƚ] | | | | | | | | |
| < 67.2 | 791 (100.0) | 588 (100.0) | 158 (100.0) | 20 (100.0) | 10,620 (46.1) | 8,437 (42.7) | 1,762 (69.8) | 144 (37.3) |
| ≥ 67.2 | 0 (0.0) | 0 (0.0) | 0 (0.0) | 0 (0.0) | 12,437 (53.9) | 11,317 (57.3) | 762 (30.2) | 242 (62.7) |
| Race and ethnicity [ƚ] | | | | | | | | |
| Hispanic/Latina | 50 (6.3) | 42 (7.1) | 7 (4.4) | 1 (5.0) | 733 (3.2) | 580 (2.9) | 93 (3.7) | 35 (9.1) |
| Non-Hispanic Black | 85 (10.7) | 53 (9.0) | 24 (15.2) | 5 (25.0) | 1,407 (6.1) | 1,095 (5.5) | 252 (10.0) | 26 (6.7) |
| Non-Hispanic White | 656 (82.9) | 493 (83.8) | 127 (80.4) | 14 (70.0) | 20,917 (90.7) | 18,079 (91.5) | 2,179 (86.3) | 325 (84.2) |
| Educational attainment [ƚ g] | | | | | | | | |
| <High school or GED | 70 (8.8) | 48 (8.2) | 15 (9.5) | 5 (25.0) | 3,134 (13.6) | 2,635 (13.3) | 369 (14.6) | 74 (19.2) |
| Some college /technical | 222 (28.1) | 155 (26.4) | 48 (30.4) | 8 (40.0) | 7,379 (32.0) | 6,237 (31.6) | 865 (34.3) | 142 (36.8) |
| ≥ Bachelor's or higher | 499 (63.1) | 385 (65.5) | 95 (60.1) | 7 (35.0) | 12,541 (54.4) | 10,880 (55.1) | 1,290 (51.1) | 170 (44.0) |
| *Missing* | *0 (0.0)* | *0 (0.0)* | *0 (0.0)* | *0 (0.0)* | *3 (0.0)* | *2 (0.0)* | *0 (0.0)* | *0 (0.0)* |
| Annual household income [ƚ] | | | | | | | | |
| < $20,000 - $49,999 | 91 (11.5) | 67 (11.4) | 18 (11.4) | 1 (5.0) | 6,520 (28.3) | 5,671 (28.7) | 567 (22.5) | 175 (45.3) |
| $50,000 – $99,999 | 209 (26.4) | 144 (24.5) | 48 (30.4) | 9 (45.0) | 8,610 (37.3) | 7,460 (37.8) | 895 (35.5) | 110 (28.5) |
| ≥ $100,000 | 491 (62.1) | 377 (64.1) | 92 (58.2) | 10 (50.0) | 7,927 (34.4) | 6,623 (33.5) | 1,062 (42.1) | 101 (26.2) |
| Marital status [ƚ] | | | | | | | | |
| Married or living as though married | 598 (75.6) | 445 (75.7) | 122 (77.2) | 16 (80.0) | 15,524 (67.3) | 13,061 (66.1) | 1,943 (77.0) | 235 (60.9) |
| Never married, divorced, widowed, or separated | 193 (24.4) | 143 (24.3) | 36 (22.8) | 4 (20.0) | 7,533 (32.7) | 6,693 (33.9) | 581 (23.0) | 151 (39.1) |
| Region of residence | | | | | | | | |
| Northeast | 131 (16.6) | 98 (16.7) | 26 (16.5) | 2 (10.0) | 3,745 (16.2) | 3,236 (16.4) | 404 (16.0) | 53 (13.7) |
| Midwest | 210 (26.5) | 157 (26.7) | 42 (26.6) | 6 (30.0) | 6,080 (26.4) | 5,218 (26.4) | 660 (26.1) | 95 (24.6) |
| South | 270 (34.1) | 197 (33.5) | 56 (35.4) | 8 (40.0) | 7,642 (33.1) | 6,521 (33.0) | 865 (34.3) | 123 (31.9) |
| West | 166 (21.0) | 124 (21.1) | 33 (20.9) | 3 (15.0) | 5,355 (23.2) | 4,600 (23.3) | 572 (22.7) | 93 (24.1) |
| Puerto Rico | 14 (1.8) | 12 (2.0) | 1 (0.6) | 1 (5.0) | 235 (1.0) | 179 (0.9) | 23 (0.9) | 22 (5.7) |
| **Clinical characteristics** | | | | | | | | |
| Body Mass Index, $kg/m^2$ [ƚ] | | | | | | | | |
| Recommended (18.5 - 24.9) | 271 (34.3) | 208 (35.4) | 44 (27.8) | 7 (35.0) | 8,819 (38.2) | 7,681 (38.9) | 833 (33.0) | 159 (41.2) |
| Overweight (25.0 - 29.9) | 229 (29.0) | 171 (29.1) | 47 (29.7) | 4 (20.0) | 7,403 (32.1) | 6,335 (32.1) | 848 (33.6) | 97 (25.1) |
| Obesity (≥ 30) | 282 (35.7) | 204 (34.7) | 63 (39.9) | 9 (45.0) | 6,539 (28.4) | 5,485 (27.8) | 815 (32.3) | 124 (32.1) |
| *Missing* | *9 (1.1)* | *5 (0.9)* | *4 (2.5)* | *0 (0.0)* | *296 (1.3)* | *253 (1.3)* | *28 (1.1)* | *6 (1.6)* |
| **Sleep health dimensions** | | | | | | | | |

| | | | | | | | | |
|---|---|---|---|---|---|---|---|---|
| Average weekly sleep duration [ǂ h] | | | | | | | | |
| Short | 112 (14.2) | 79 (13.4) | 26 (16.5) | 5 (25.0) | 2,732 (11.8) | 2,269 (11.5) | 343 (13.6) | 62 (16.1) |
| Recommended | 573 (72.4) | 435 (74.0) | 108 (68.4) | 12 (60.0) | 16,321 (70.8) | 14,128 (71.5) | 1,717 (68.0) | 230 (59.6) |
| Long | 101 (12.8) | 71 (12.1) | 23 (14.6) | 3 (15.0) | 3,717 (16.1) | 3,116 (15.8) | 438 (17.4) | 80 (20.7) |
| *Missing* | *5 (0.6)* | *3 (0.5)* | *1 (0.6)* | *0 (0.0)* | *287 (1.2)* | *241 (1.2)* | *26 (1.0)* | *14 (3.6)* |
| Long sleep onset latency [i] | | | | | | | | |
| Yes | 125 (15.8) | 76 (12.9) | 36 (22.8) | 2 (10.0) | 4,057 (17.6) | 3,120 (15.8) | 653 (25.9) | 118 (30.6) |
| No | 659 (83.3) | 508 (86.4) | 120 (75.9) | 18 (90.0) | 18,799 (81.5) | 16,465 (83.4) | 1,853 (73.4) | 260 (67.4) |
| *Missing* | *7 (0.9)* | *4 (0.7)* | *2 (1.3)* | *0 (0.0)* | *201 (0.9)* | *169 (0.9)* | *18 (0.7)* | *8 (2.1)* |
| Poor sleep maintenance [ǂ j] | | | | | | | | |
| Yes | 478 (60.4) | 330 (56.1) | 117 (74.1) | 14 (70.0) | 12,779 (55.4) | 10,379 (52.5) | 1,847 (73.2) | 248 (64.2) |
| No | 308 (38.9) | 256 (43.5) | 38 (24.1) | 6 (30.0) | 10,093 (43.8) | 9,207 (46.6) | 667 (26.4) | 136 (35.2) |
| *Missing* | *5 (0.6)* | *2 (0.3)* | *3 (1.9)* | *0 (0.0)* | *185 (0.8)* | *168 (0.9)* | *10 (0.4)* | *2 (0.5)* |
| Insomnia symptoms [ǂ k] | | | | | | | | |
| Yes | 512 (64.7) | 354 (60.2) | 125 (79.1) | 14 (70.0) | 13,840 (60.0) | 11,264 (57.0) | 1,977 (78.3) | 265 (68.7) |
| No | 277 (35.0) | 232 (39.5) | 33 (20.9) | 6 (30.0) | 9,152 (39.7) | 8,433 (42.7) | 544 (21.6) | 119 (30.8) |
| *Missing* | *2 (0.3)* | *2 (0.3)* | *0 (0.0)* | *0 (0.0)* | *65 (0.3)* | *57 (0.3)* | *3 (0.1)* | *2 (0.5)* |
| Sleep medication use [ǂ l] | | | | | | | | |
| Yes | 138 (17.4) | 100 (17.0) | 28 (17.7) | 5 (25.0) | 4,939 (21.4) | 4,119 (20.9) | 595 (23.6) | 109 (28.2) |
| No | 650 (82.2) | 485 (82.5) | 130 (82.3) | 15 (75.0) | 18,019 (78.1) | 15,557 (78.8) | 1,917 (76.0) | 273 (70.7) |
| *Missing* | *3 (0.4)* | *3 (0.5)* | *0 (0.0)* | *0 (0.0)* | *99 (0.4)* | *78 (0.4)* | *12 (0.5)* | *4 (1.0)* |
| Daytime dysfunction [m] | | | | | | | | |
| Yes | 7 (0.9) | 4 (0.7) | 1 (0.6) | 2 (10.0) | 211 (0.9) | 150 (0.8) | 35 (1.4) | 14 (3.6) |
| No | 777 (98.2) | 579 (98.5) | 155 (98.1) | 18 (90.0) | 22,575 (97.9) | 19,370 (98.1) | 2,460 (97.5) | 368 (95.3) |
| *Missing* | *7 (0.9)* | *5 (0.9)* | *2 (1.3)* | *0 (0.0)* | *271 (1.2)* | *234 (1.2)* | *29 (1.1)* | *4 (1.0)* |
| Healthcare provider diagnosed sleep apnea [ǂ n] | | | | | | | | |
| Yes | 60 (7.6) | 45 (7.7) | 11 (7.0) | 1 (5.0) | 2,503 (10.9) | 2,071 (10.5) | 314 (12.4) | 70 (18.1) |
| No | 726 (91.8) | 538 (91.5) | 147 (93.0) | 19 (95.0) | 20,293 (88.0) | 17,460 (88.4) | 2,191 (86.8) | 305 (79.0) |
| *Missing* | *5 (0.6)* | *5 (0.9)* | *0 (0.0)* | *0 (0.0)* | *261 (1.1)* | *223 (1.1)* | *19 (0.8)* | *11 (2.8)* |

Abbreviations: SD, Standard Deviation; GED: General Educational Development

Note: N = 418 (1.8%) participants who reported both 'too hot' and 'too cold' temperatures were not included in this sensitivity analysis.

[¥] Chi-square test *p*-values indicate significant differences between the overall prevalence of ‘too hot’ by menopausal status.

[§] Chi-square test *p*-values indicate significant differences between the overall prevalence of ‘too cold’ by menopausal status.

[ǂ] Chi-square or Analysis of Variance (ANOVA) test *p*-values indicate significant differences in characteristics between the total populations of each menopausal status category.

[a] Reasons for trouble sleeping ≥ 3 times per week in the past month include: unable to fall asleep within 30 minutes, waking up in the middle of the night or early morning, waking up to use the bathroom, cannot breathe comfortably, coughing or snoring loudly, having bad dreams, having pain, or other non-specified reasons.

[b] Excluded for reporting both ‘Too hot’ and ‘Too cold’, Premenopausal: n = 25 (3.2%).

[c] Excluded for reporting both ‘Too hot’ and ‘Too cold’, Postmenopausal: n = 393 (1.7%).

[d] ‘Infrequent temperature extremes’ is defined as self-reported trouble sleeping due to feeling too hot < 3 times per week and trouble sleeping due to feeling too cold < 3 times per week in the past month.

[e] ’Too hot’ is defined as self-reported trouble sleeping due to feeling too hot ≥ 3 times per week in the past month.

[f] ’Too cold’ is defined as self-reported trouble sleeping due to feeling too cold ≥ 3 times per week in the past month.

[g] Educational attainment was assessed at baseline.

[h] Sleep duration is based on reported bed and wake times or reported average sleep duration. Participants who reported ≤ 2 or ≥ 23 hours of sleep are excluded. Short: < 7 hours; Recommended: 7-9 hours; Long ≥ 9 hours.

[i] Long sleep onset latency is defined as not falling asleep within 30 minutes at least three times a week during the past month.

[j] Poor sleep maintenance is defined as waking up in the middle of the night or early morning at least three times a week during the past month.
[k] Insomnia symptoms is defined as long sleep onset latency or poor sleep maintenance.
[l] Sleep medication use is defined as 'taking medicine (prescription or over the counter) to help you sleep' at least three times a week during the past month.
[m] Daytime dysfunction is defined as having trouble staying awake while driving, eating, or engaging in social activity at least three times a week during the past month.
[n] Healthcare provider diagnosed sleep apnea is defined as a current doctor or other health professional diagnosis of sleep apnea.

**Supplemental Table 16. Prevalence ratios (95% confidence intervals) for associations between perceived indoor temperature extremes and sleep health dimensions among participants who reported trouble sleeping for any reason [a], Sister Study, 2017-2019**

| | Indoor temperature: Too hot [b] | Indoor temperature: Too cold [c] |
|---|---|---|
| Weekly sleep duration [d] | | |
| Short | **1.14 (1.03,1.27)‡** | **1.43 (1.16,1.76)‡** |
| Recommended | 1.00 (ref) | 1.00 (ref) |
| Long | **1.23 (1.12,1.35)‡** | **1.36 (1.13,1.65)‡** |
| Long sleep onset latency [e] | | |
| Yes | **1.80 (1.66,1.96)‡** | **1.87 (1.58,2.21)‡** |
| No | 1.00 (ref) | 1.00 (ref) |
| Poor sleep maintenance [f] | | |
| Yes | **1.43 (1.38,1.47)‡** | **1.26 (1.16,1.36)‡** |
| No | 1.00 (ref) | 1.00 (ref) |
| Insomnia symptoms [g] | | |
| Yes | **1.42 (1.38,1.46)‡** | **1.24 (1.15,1.34)‡** |
| No | 1.00 (ref) | 1.00 (ref) |
| Sleep medication use [h] | | |
| Yes | **1.18 (1.09,1.27)‡** | **1.34 (1.14,1.57)‡** |
| No | 1.00 (ref) | 1.00 (ref) |
| Daytime dysfunction [i] | | |
| Yes | **1.75 (1.21,2.54)‡** | **4.86 (2.91,8.09)‡** |
| No | 1.00 (ref) | 1.00 (ref) |
| Healthcare professional diagnosed sleep apnea [j] | | |
| Yes | **1.16 (1.04,1.29)‡** | **1.65 (1.35,2.03)‡** |
| No | 1.00 (ref) | 1.00 (ref) |

Note: N = 418 (1.8%) participants who reported both 'too hot' and 'too cold' temperatures were not included in this sensitivity analysis.

Models are adjusted for age (continuous), age$^2$, race and ethnicity (Hispanic/Latina, non-Hispanic Black, non-Hispanic White), annual household income (<$20,000 - $49,999, $50,000 - $99,999, ≥$100,000), marital status (married or living as though married, divorced/widowed/separated/never married), region of residence (Northeast, Midwest, South, West, and Puerto Rico), body mass index (BMI: underweight, recommended, overweight, obesity), and menopausal status (premenopausal, postmenopausal).

[a] In models for weekly sleep duration, sleep medication use, daytime dysfunction, and healthcare professional diagnosed sleep apnea, reasons for trouble sleeping for any reason other than feeling too hot or feeling too cold ≥ 3 times per week in the past month include: unable to fall asleep within 30 minutes, waking up in the middle of the night or early morning, waking up to use the bathroom, cannot breathe comfortably, coughing or snoring loudly, having bad dreams, having pain, or other non-specified reasons (n = 23,853). In models for long sleep onset latency, poor sleep maintenance, and insomnia symptoms, reasons for trouble sleeping for any reason other than feeling too hot or feeling too cold ≥ 3 times per week in the past month include waking up to use the bathroom, cannot breathe comfortably, coughing or snoring loudly, having bad dreams, having pain, or other non-specified reasons (n = 21,641).
[b] 'Too hot' is defined as self-reported trouble sleeping due to feeling too hot ≥ 3 times per week vs. trouble sleeping due to feeling too hot < 3 times per week and trouble sleeping due to feeling too cold < 3 times per week.

[c] 'Too cold' is defined as self-reported trouble sleeping due to feeling too cold ≥ 3 times per week vs. trouble sleeping due to feeling too hot < 3 times per week and trouble sleeping due to feeling too cold < 3 times per week.
[d] Sleep duration is based on reported bed and wake times or reported average sleep duration. Participants who reported ≤ 2 or ≥ 23 hours of sleep are excluded. Short: <7 hours; Recommended: 7-9 hours; Long ≥ 9 hours.
[e] Long sleep onset latency is defined as not falling asleep within 30 minutes at least three times a week during the past month.
[f] Poor sleep maintenance is defined as waking up in the middle of the night or early morning at least three times a week during the past month.
[g] Insomnia symptoms is defined as long sleep onset latency or poor sleep maintenance.
[h] Sleep medication use is defined as 'taking medicine (prescription or over the counter) to help you sleep' at least three times a week during the past month.
[i] Daytime dysfunction is defined as having trouble staying awake while driving, eating, or engaging in social activity at least three times a week during the past month.
[j] Healthcare provider diagnosed sleep apnea is defined as a current doctor or other health professional diagnosis of sleep apnea.
[ł] Remained significant after false discovery rate correction. False discovery rate-corrected *p*-value was considered statistically significant at the $\alpha = 0.05$ level.

**Supplemental Table 17A. Prevalence ratios (95% confidence intervals) for associations between perceived indoor temperature extremes and sleep health dimensions by race and ethnicity, among participants who reported trouble sleeping for any reason [a], Sister Study, 2017-2019**

| | Indoor temperature: Too hot [b] | | | | Indoor temperature: Too cold [c] | | | |
|---|---|---|---|---|---|---|---|---|
| | Hispanic/Latina | Non-Hispanic Black | Non-Hispanic White | Wald *p*-values | Hispanic/Latina | Non-Hispanic Black | Non-Hispanic White | Wald *p*-values |
| Weekly sleep duration [d] | | | | | | | | |
| Short | 1.15 (0.73,1.82) | 1.06 (0.87,1.29) | **1.17 (1.03,1.32)‡** | 0.7029 | 0.73 (0.28,1.88) | **1.54 (1.07,2.19)‡** | **1.51 (1.18,1.94)‡** | 0.3298 |
| Recommended | 1.00 (Ref) | 1.00 (Ref) | 1.00 (Ref) | --- | 1.00 (Ref) | 1.00 (Ref) | 1.00 (Ref) | --- |
| Long | 1.42 (0.93,2.16) | 0.98 (0.70,1.38) | **1.24 (1.13,1.37)‡** | 0.3402 | 1.15 (0.56,2.38) | 1.46 (0.62,3.41) | **1.38 (1.13,1.69)‡** | 0.8859 |
| Long sleep onset latency [e] | | | | | | | | |
| Yes | **2.03 (1.51,2.72)‡** | **2.02 (1.61,2.52)‡** | **1.76 (1.61,1.93)‡** | 0.3987 | 1.50 (0.92,2.45) | 1.13 (0.51,2.51) | **1.99 (1.66,2.39)‡** | 0.2492 |
| No | 1.00 (Ref) | 1.00 (Ref) | 1.00 (Ref) | --- | 1.00 (Ref) | 1.00 (Ref) | 1.00 (Ref) | --- |
| Poor sleep maintenance [f] | | | | | | | | |
| Yes | **1.45 (1.22,1.72)‡** | **1.36 (1.21,1.53)‡** | **1.43 (1.39,1.48)‡** | 0.6953 | **1.46 (1.10,1.94)‡** | **1.37 (1.03,1.82)‡** | **1.23 (1.13,1.35)‡** | 0.4502 |
| No | 1.00 (Ref) | 1.00 (Ref) | 1.00 (Ref) | --- | 1.00 (Ref) | 1.00 (Ref) | 1.00 (Ref) | --- |
| Insomnia symptoms [g] | | | | | | | | |
| Yes | **1.50 (1.30,1.74)‡** | **1.39 (1.25,1.55)‡** | **1.42 (1.38,1.46)‡** | 0.6751 | **1.38 (1.06,1.79)‡** | **1.38 (1.06,1.80)‡** | **1.22 (1.12,1.32)‡** | 0.4668 |
| No | 1.00 (Ref) | 1.00 (Ref) | 1.00 (Ref) | --- | 1.00 (Ref) | 1.00 (Ref) | 1.00 (Ref) | --- |
| Sleep medication use [h] | | | | | | | | |
| Yes | 1.06 (0.68,1.67) | 1.15 (0.83,1.58) | **1.19 (1.10,1.28)‡** | 0.8813 | 1.37 (0.82,2.30) | 1.03 (0.41,2.61) | **1.35 (1.14,1.60)‡** | 0.8529 |
| No | 1.00 (Ref) | 1.00 (Ref) | 1.00 (Ref) | --- | 1.00 (Ref) | 1.00 (Ref) | 1.00 (Ref) | --- |
| Daytime dysfunction [i] | | | | | | | | |
| Yes | 0.69 (0.08,5.95) | 2.01 (0.60,6.69) | **1.81 (1.22,2.68)‡** | 0.6754 | **6.40 (1.56,26.28)‡** | **12.60 (3.37,47.10)‡** | **4.05 (2.19,7.51)‡** | 0.2910 |
| No | 1.00 (Ref) | 1.00 (Ref) | 1.00 (Ref) | --- | 1.00 (Ref) | 1.00 (Ref) | 1.00 (Ref) | --- |
| Healthcare professional diagnosed sleep apnea [j] | | | | | | | | |
| Yes | 1.39 (0.79,2.44) | 1.27 (0.96,1.67) | **1.13 (1.00,1.28)‡** | 0.6088 | **2.71 (1.39,5.27)‡** | **2.11 (1.19,3.75)‡** | **1.53 (1.21,1.93)‡** | 0.2008 |
| No | 1.00 (Ref) | 1.00 (Ref) | 1.00 (Ref) | --- | 1.00 (Ref) | 1.00 (Ref) | 1.00 (Ref) | --- |

Note: N = 418 (1.8%) participants who reported both 'too hot' and 'too cold' temperatures were not included in this sensitivity analysis.

Models are adjusted for age (continuous), age$^2$, annual household income (<$20,000 - $49,999, $50,000 - $99,999, ≥$100,000), marital status (married or living as though married, divorced/widowed/separated/never married), region of residence (Northeast, Midwest, South, West, and Puerto Rico), body mass index (BMI: underweight, recommended, overweight, obesity), and menopausal status (premenopausal, postmenopausal).

[a] In models for weekly sleep duration, sleep medication use, daytime dysfunction, and healthcare professional diagnosed sleep apnea, reasons for trouble sleeping for any reason other than feeling too hot or feeling too cold ≥ 3 times per week in the past month include: unable to fall asleep within 30 minutes, waking up in the middle of the night or early morning, waking up to use the bathroom, cannot breathe comfortably, coughing or snoring loudly, having bad dreams, having pain, or other non-specified reasons (n = 23,853). In models for long sleep onset latency, poor sleep maintenance, and insomnia symptoms, reasons for trouble sleeping for any reason other than feeling too hot or feeling too cold ≥ 3 times per week in the past month include waking up to use the bathroom, cannot breathe comfortably, coughing or snoring loudly, having bad dreams, having pain, or other non-specified reasons (n = 21,641).
[b] Too hot' is defined as self-reported trouble sleeping due to feeling too hot ≥ 3 times per week vs. trouble sleeping due to feeling too hot < 3 times per week and trouble sleeping due to feeling too cold < 3 times per week.
[c] 'Too cold' is defined as self-reported trouble sleeping due to feeling too cold ≥ 3 times per week vs. trouble sleeping due to feeling too hot < 3 times per week and trouble sleeping due to feeling too cold < 3 times per week.
[d] Sleep duration is based on reported bed and wake times or reported average sleep duration. Participants who reported ≤ 2 or ≥ 23 hours of sleep are excluded. Short: <7 hours; Recommended: 7-9 hours; Long ≥ 9 hours.
[e] Long sleep onset latency is defined as not falling asleep within 30 minutes at least three times a week during the past month.
[f] Poor sleep maintenance is defined as waking up in the middle of the night or early morning at least three times a week during the past month.
[g] Insomnia symptoms is defined as long sleep onset latency or poor sleep maintenance.
[h] Sleep medication use is defined as 'taking medicine (prescription or over the counter) to help you sleep' at least three times a week during the past month.

[i] Daytime dysfunction is defined as having trouble staying awake while driving, eating, or engaging in social activity at least three times a week during the past month.
[j] Healthcare provider diagnosed sleep apnea is defined as a current doctor or other health professional diagnosis of sleep apnea.
[ł] Remained significant after false discovery rate correction. False discovery rate-corrected *p*-value was considered statistically significant at the $\alpha = 0.05$ level.

**Supplemental Table 17B. Relative excess risk due to interaction (RERI) between perceived indoor temperature extremes and race and ethnicity, among participants who reported trouble sleeping for any reason [a], Sister Study, 2017-2019**

| | Indoor temperature: Too hot [a] | | | Indoor temperature: Too cold [b] | | |
|---|---|---|---|---|---|---|
| | **Hispanic/Latina** | **Non-Hispanic Black** | **Non-Hispanic White** | **Hispanic/Latina** | **Non-Hispanic Black** | **Non-Hispanic White** |
| Weekly sleep duration [d] | | | | | | |
| Short | 0.05 (-0.71, 0.81) | -0.02 (-0.54, 0.50) | | -0.93 (-2.07, 0.21) | 0.80 (-0.56, 2.16) | |
| Recommended | --- | --- | 1.00 (Ref) | --- | --- | 1.00 (Ref) |
| Long | 0.23 (-0.41, 0.87) | -0.26 (-0.62, 0.10) | | -0.20 (-1.21, 0.81) | 0.09 (-1.19, 1.37) | |
| Long sleep onset latency [e] | | | | | | |
| Yes | 0.71 (-0.05, 1.47) | 0.27 (-0.14, 0.68) | 1.00 (Ref) | -0.33 (-1.34, 0.68) | -0.86 (-1.85, 0.13) | 1.00 (Ref) |
| No | --- | --- | | --- | --- | |
| Poor sleep maintenance [f] | | | | | | |
| Yes | -0.02 (-0.22, 0.18) | -0.11 (-0.24, 0.02) | 1.00 (Ref) | 0.19 (-0.18, 0.56) | 0.10 (-0.26, 0.46) | 1.00 (Ref) |
| No | --- | --- | | --- | --- | |
| Insomnia symptoms [g] | | | | | | |
| Yes | 0.06 (-0.11, 0.23) | -0.07 (-0.19, 0.05) | 1.00 (Ref) | 0.15 (-0.18, 0.48) | 0.12 (-0.21, 0.45) | 1.00 (Ref) |
| No | --- | --- | | --- | --- | |
| Sleep medication use [h] | | | | | | |
| Yes | -0.14 (-0.49, 0.21) | -0.10 (-0.33, 0.13) | 1.00 (Ref) | -0.09 (-0.62, 0.44) | -0.33 (-0.95, 0.29) | 1.00 (Ref) |
| No | --- | --- | | --- | --- | |
| Daytime dysfunction [i] | | | | | | |
| Yes | -1.42 (-4.44, 1.60) | -0.04 (-1.77, 1.69) | 1.00 (Ref) | 8.21 (-8.76, 25.18) | 6.01 (-5.59, 17.61) | 1.00 (Ref) |
| No | --- | --- | | --- | --- | |
| Healthcare professional diagnosed sleep apnea [j] | | | | | | |
| Yes | 0.21 (-0.43, 0.85) | 0.14 (-0.22, 0.50) | 1.00 (Ref) | 0.94 (-0.54, 2.42) | 0.60 (-0.65, 1.85) | 1.00 (Ref) |
| No | --- | --- | | --- | --- | |

Note: N = 418 (1.8%) participants who reported both 'too hot' and 'too cold' temperatures were not included in this sensitivity analysis.
RERI = 0: no additive interaction; RERI >0: positive additive interaction; RERI <0: negative additive interaction.

Models are adjusted for age (continuous), age$^2$, annual household income (<$20,000 - $49,999, $50,000 - $99,999, ≥$100,000), marital status (married or living as though married, divorced/widowed/separated/never married), region of residence (Northeast, Midwest, South, West, and Puerto Rico), body mass index (BMI: underweight, recommended, overweight, obesity), and menopausal status (premenopausal, postmenopausal).

[a] In models for weekly sleep duration, sleep medication use, daytime dysfunction, and healthcare professional diagnosed sleep apnea, reasons for trouble sleeping for any reason other than feeling too hot or feeling too cold ≥ 3 times per week in the past month include: unable to fall asleep within 30 minutes, waking up in the middle of the night or early morning, waking up to use the bathroom, cannot breathe comfortably, coughing or snoring loudly, having bad dreams, having pain, or other non-specified reasons (n = 23,853). In models for long sleep onset latency, poor sleep maintenance, and insomnia symptoms, reasons for trouble sleeping for any reason other than feeling too hot or feeling too cold ≥ 3 times per week in the past month include waking up to use the bathroom, cannot breathe comfortably, coughing or snoring loudly, having bad dreams, having pain, or other non-specified reasons (n = 21,641).
[b] Too hot' is defined as self-reported trouble sleeping due to feeling too hot ≥ 3 times per week vs. trouble sleeping due to feeling too hot < 3 times per week and trouble sleeping due to feeling too cold < 3 times per week.
[c] 'Too cold' is defined as self-reported trouble sleeping due to feeling too cold ≥ 3 times per week vs. trouble sleeping due to feeling too hot < 3 times per week and trouble sleeping due to feeling too cold < 3 times per week.
[d] Sleep duration is based on reported bed and wake times or reported average sleep duration. Participants who reported ≤ 2 or ≥ 23 hours of sleep are excluded. Short: <7 hours; Recommended: 7-9 hours; Long ≥ 9 hours.
[e] Long sleep onset latency is defined as not falling asleep within 30 minutes at least three times a week during the past month.
[f] Poor sleep maintenance is defined as waking up in the middle of the night or early morning at least three times a week during the past month.
[g] Insomnia symptoms is defined as long sleep onset latency or poor sleep maintenance.
[h] Sleep medication use is defined as 'taking medicine (prescription or over the counter) to help you sleep' at least three times a week during the past month.
[i] Daytime dysfunction is defined as having trouble staying awake while driving, eating, or engaging in social activity at least three times a week during the past month.

[j] Healthcare provider diagnosed sleep apnea is defined as a current doctor or other health professional diagnosis of sleep apnea.

**Supplemental Table 18A. Prevalence ratios (95% confidence intervals) for associations between perceived indoor temperature extremes and sleep health dimensions by annual household income, among participants who reported trouble sleeping for any reason [a], Sister Study, 2017-2019**

| | Indoor temperature: Too hot [b] | | | | Indoor temperature: Too cold [c] | | | |
|---|---|---|---|---|---|---|---|---|
| | **<$20,000 - $49,999** | **$50,000 - $99,999** | **≥ $100,000** | **Wald *p*-values** | **<$20,000 - $49,999** | **$50,000 - $99,999** | **≥ $100,000** | **Wald *p*-values** |
| Weekly sleep duration [d] | | | | | | | | |
| Short | 1.02 (0.83,1.25) | **1.26 (1.07,1.48)‡** | 1.12 (0.94,1.33) | 0.2460 | **1.44 (1.07,1.93)‡** | 1.43 (0.96,2.14) | 1.41 (0.92,2.17) | 0.9973 |
| Recommended | 1.00 (Ref) | 1.00 (Ref) | 1.00 (Ref) | --- | 1.00 (Ref) | 1.00 (Ref) | 1.00 (Ref) | --- |
| Long | **1.22 (1.04,1.43)‡** | 1.13 (0.97,1.33) | **1.33 (1.15,1.54)‡** | 0.3291 | 1.31 (0.99,1.73) | 1.04 (0.68,1.58) | **1.88 (1.35,2.60)‡** | 0.0730 |
| Long sleep onset latency [e] | | | | | | | | |
| Yes | **1.72 (1.49,1.98)‡** | **1.98 (1.75,2.25)‡** | **1.68 (1.46,1.95)‡** | 0.1590 | **1.84 (1.47,2.31)‡** | **1.85 (1.33,2.56)‡** | **1.95 (1.33,2.88)‡** | 0.9654 |
| No | 1.00 (Ref) | 1.00 (Ref) | 1.00 (Ref) | --- | 1.00 (Ref) | 1.00 (Ref) | 1.00 (Ref) | --- |
| Poor sleep maintenance [f] | | | | | | | | |
| Yes | **1.41 (1.32,1.50)‡** | **1.42 (1.35,1.50)‡** | **1.45 (1.38,1.52)‡** | 0.7509 | **1.27 (1.12,1.43)‡** | **1.31 (1.14,1.52)‡** | **1.18 (1.00,1.39)‡** | 0.6297 |
| No | 1.00 (Ref) | 1.00 (Ref) | 1.00 (Ref) | --- | 1.00 (Ref) | 1.00 (Ref) | 1.00 (Ref) | --- |
| Insomnia symptoms [g] | | | | | | | | |
| Yes | **1.39 (1.31,1.47)‡** | **1.42 (1.36,1.49)‡** | **1.43 (1.37,1.50)‡** | 0.6617 | **1.24 (1.11,1.39)‡** | **1.27 (1.11,1.46)‡** | **1.21 (1.03,1.41)‡** | 0.8874 |
| No | 1.00 (Ref) | 1.00 (Ref) | 1.00 (Ref) | --- | 1.00 (Ref) | 1.00 (Ref) | 1.00 (Ref) | --- |
| Sleep medication use [h] | | | | | | | | |
| Yes | **1.26 (1.09,1.45)‡** | **1.21 (1.07,1.37)‡** | 1.10 (0.97,1.25) | 0.3580 | **1.39 (1.11,1.75)‡** | 1.01 (0.71,1.43) | **1.62 (1.23,2.15)‡** | 0.1151 |
| No | 1.00 (Ref) | 1.00 (Ref) | 1.00 (Ref) | --- | 1.00 (Ref) | 1.00 (Ref) | 1.00 (Ref) | --- |
| Daytime dysfunction [i] | | | | | | | | |
| Yes | 1.47 (0.76,2.84) | **2.18 (1.17,4.05)‡** | 1.67 (0.89,3.13) | 0.6793 | **2.66 (1.10,6.42)‡** | **9.17 (4.23,19.86)‡** | **5.64 (2.09,15.19)‡** | 0.1176 |
| No | 1.00 (Ref) | 1.00 (Ref) | 1.00 (Ref) | --- | 1.00 (Ref) | 1.00 (Ref) | 1.00 (Ref) | --- |
| Healthcare professional diagnosed sleep apnea [j] | | | | | | | | |
| Yes | 1.18 (0.97,1.44) | 1.13 (0.95,1.35) | 1.16 (0.96,1.39) | 0.9457 | **1.56 (1.16,2.09)‡** | **1.84 (1.27,2.66)‡** | **1.63 (1.03,2.57)‡** | 0.7786 |
| No | 1.00 (Ref) | 1.00 (Ref) | 1.00 (Ref) | --- | 1.00 (Ref) | 1.00 (Ref) | 1.00 (Ref) | --- |

Note: N = 418 (1.8%) participants who reported both 'too hot' and 'too cold' temperatures were not included in this sensitivity analysis.

Models are adjusted for age (continuous), age$^2$, race and ethnicity (Hispanic/Latina, non-Hispanic Black, non-Hispanic White), marital status (married or living as though married, divorced/widowed/separated/never married), region of residence (Northeast, Midwest, South, West, and Puerto Rico), body mass index (BMI: underweight, recommended, overweight, obesity), and menopausal status (premenopausal, postmenopausal).

[a] In models for weekly sleep duration, sleep medication use, daytime dysfunction, and healthcare professional diagnosed sleep apnea, reasons for trouble sleeping for any reason other than feeling too hot or feeling too cold ≥ 3 times per week in the past month include: unable to fall asleep within 30 minutes, waking up in the middle of the night or early morning, waking up to use the bathroom, cannot breathe comfortably, coughing or snoring loudly, having bad dreams, having pain, or other non-specified reasons (n = 23,853). In models for long sleep onset latency, poor sleep maintenance, and insomnia symptoms, reasons for trouble sleeping for any reason other than feeling too hot or feeling too cold ≥ 3 times per week in the past month include waking up to use the bathroom, cannot breathe comfortably, coughing or snoring loudly, having bad dreams, having pain, or other non-specified reasons (n = 21,641).
[b] Too hot' is defined as self-reported trouble sleeping due to feeling too hot ≥ 3 times per week vs. trouble sleeping due to feeling too hot < 3 times per week and trouble sleeping due to feeling too cold < 3 times per week.
[c] 'Too cold' is defined as self-reported trouble sleeping due to feeling too cold ≥ 3 times per week vs. trouble sleeping due to feeling too hot < 3 times per week and trouble sleeping due to feeling too cold < 3 times per week.
[d] Sleep duration is based on reported bed and wake times or reported average sleep duration. Participants who reported ≤ 2 or ≥ 23 hours of sleep are excluded. Short: <7 hours; Recommended: 7-9 hours; Long ≥ 9 hours.
[e] Long sleep onset latency is defined as not falling asleep within 30 minutes at least three times a week during the past month.
[f] Poor sleep maintenance is defined as waking up in the middle of the night or early morning at least three times a week during the past month.
[g] Insomnia symptoms is defined as long sleep onset latency or poor sleep maintenance.
[h] Sleep medication use is defined as 'taking medicine (prescription or over the counter) to help you sleep' at least three times a week during the past month.

[i] Daytime dysfunction is defined as having trouble staying awake while driving, eating, or engaging in social activity at least three times a week during the past month.
[j] Healthcare provider diagnosed sleep apnea is defined as a current doctor or other health professional diagnosis of sleep apnea.
[ƚ] Remained significant after false discovery rate correction. False discovery rate-corrected *p*-value was considered statistically significant at the $\alpha = 0.05$ level.

**Supplemental Table 18B. Relative excess risk due to interaction (RERI) between perceived temperature extremes and annual household income, among participants who reported trouble sleeping for any reason [a], Sister Study, 2017-2019**

| | Indoor temperature: Too hot [b] | | | Indoor temperature: Too cold [c] | | |
|---|---|---|---|---|---|---|
| | **< $20,000 - $49,999** | **$50,000 - $99,999** | **≥ $100,000** | **< $20,000 - $49,999** | **$50,000 - $99,999** | **≥ $100,000** |
| Weekly sleep duration [d] | | | | | | |
| Short | -0.09 (-0.40, 0.22) | 0.16 (-0.11, 0.43) | | 0.15 (-0.65, 0.95) | 0.05 (-0.80, 0.90) | |
| Recommended | --- | --- | 1.00 (Ref) | --- | --- | 1.00 (Ref) |
| Long | -0.06 (-0.40, 0.28) | -0.28 (-0.56, 0.00) | | -0.43 (-1.22, 0.36) | **-0.83 (-1.62, -0.04)** | |
| Long sleep onset latency [e] | | | | | | |
| Yes | **0.39 (0.01, 0.77)** | **0.49 (0.16, 0.82)** | 1.00 (Ref) | 0.35 (-0.62, 1.32) | 0.07 (-0.97, 1.11) | 1.00 (Ref) |
| No | --- | --- | | --- | --- | |
| Poor sleep maintenance [f] | | | | | | |
| Yes | -0.04 (-0.12, 0.04) | -0.03 (-0.10, 0.04) | 1.00 (Ref) | 0.09 (-0.15, 0.33) | 0.13 (-0.14, 0.40) | 1.00 (Ref) |
| No | --- | --- | | --- | --- | |
| Insomnia symptoms [g] | | | | | | |
| Yes | -0.01 (-0.07, 0.05) | -0.01 (-0.06, 0.04) | 1.00 (Ref) | 0.04 (-0.19, 0.27) | 0.07 (-0.18, 0.32) | 1.00 (Ref) |
| No | --- | --- | | --- | --- | |
| Sleep medication use [h] | | | | | | |
| Yes | 0.20 (-0.04, 0.44) | 0.12 (-0.08, 0.32) | 1.00 (Ref) | -0.16 (-0.74, 0.42) | **-0.61 (-1.20, -0.02)** | 1.00 (Ref) |
| No | --- | --- | | --- | --- | |
| Daytime dysfunction [i] | | | | | | |
| Yes | 0.12 (-1.72, 1.96) | 0.46 (-1.03, 1.95) | 1.00 (Ref) | -1.81 (-8.52, 4.90) | 3.35 (-5.15, 11.85) | 1.00 (Ref) |
| No | --- | --- | | --- | --- | |
| Healthcare professional diagnosed sleep apnea [j] | | | | | | |
| Yes | 0.04 (-0.28, 0.36) | -0.02 (-0.30, 0.26) | 1.00 (Ref) | -0.02 (-0.90, 0.86) | 0.24 (-0.76, 1.24) | 1.00 (Ref) |
| No | --- | --- | | --- | --- | |

Note: N = 418 (1.8%) participants who reported both 'too hot' and 'too cold' temperatures were not included in this sensitivity analysis.
RERI = 0: no additive interaction; RERI >0: positive additive interaction; RERI <0: negative additive interaction.

Models are adjusted for age (continuous), age$^2$, race and ethnicity (Hispanic/Latina, non-Hispanic Black, non-Hispanic White), marital status (married or living as though married, divorced/widowed/separated/never married), region of residence (Northeast, Midwest, South, West, and Puerto Rico), body mass index (BMI: underweight, recommended, overweight, obesity), and menopausal status (premenopausal, postmenopausal).

[a] In models for weekly sleep duration, sleep medication use, daytime dysfunction, and healthcare professional diagnosed sleep apnea, reasons for trouble sleeping for any reason other than feeling too hot or feeling too cold ≥ 3 times per week in the past month include: unable to fall asleep within 30 minutes, waking up in the middle of the night or early morning, waking up to use the bathroom, cannot breathe comfortably, coughing or snoring loudly, having bad dreams, having pain, or other non-specified reasons (n = 23,853). In models for long sleep onset latency, poor sleep maintenance, and insomnia symptoms, reasons for trouble sleeping for any reason other than feeling too hot or feeling too cold ≥ 3 times per week in the past month include waking up to use the bathroom, cannot breathe comfortably, coughing or snoring loudly, having bad dreams, having pain, or other non-specified reasons (n = 21,641).
[b] Too hot' is defined as self-reported trouble sleeping due to feeling too hot ≥ 3 times per week vs. trouble sleeping due to feeling too hot < 3 times per week and trouble sleeping due to feeling too cold < 3 times per week.
[c] 'Too cold' is defined as self-reported trouble sleeping due to feeling too cold ≥ 3 times per week vs. trouble sleeping due to feeling too hot < 3 times per week and trouble sleeping due to feeling too cold < 3 times per week.

[d] Sleep duration is based on reported bed and wake times or reported average sleep duration. Participants who reported ≤ 2 or ≥ 23 hours of sleep are excluded. Short: <7 hours; Recommended: 7-9 hours; Long ≥ 9 hours.
[e] Long sleep onset latency is defined as not falling asleep within 30 minutes at least three times a week during the past month.
[f] Poor sleep maintenance is defined as waking up in the middle of the night or early morning at least three times a week during the past month.
[g] Insomnia symptoms is defined as long sleep onset latency or poor sleep maintenance.
[h] Sleep medication use is defined as 'taking medicine (prescription or over the counter) to help you sleep' at least three times a week during the past month.
[i] Daytime dysfunction is defined as having trouble staying awake while driving, eating, or engaging in social activity at least three times a week during the past month.
[j] Healthcare provider diagnosed sleep apnea is defined as a current doctor or other health professional diagnosis of sleep apnea.

**Supplemental Table 19A. Prevalence ratios (95% confidence intervals) for associations between perceived indoor temperature extremes and sleep health dimensions by region of residence, among participants who reported trouble sleeping for any reason [a], Sister Study, 2017-2019**

| | Indoor temperature: Too hot [b] | | | | | | Indoor temperature: Too cold [c] | | | | | |
|---|---|---|---|---|---|---|---|---|---|---|---|---|
| | **Northeast** | **Midwest** | **South** | **West** | **Puerto Rico** | **Wald *p*-values** | **Northeast** | **Midwest** | **South** | **West** | **Puerto Rico** | **Wald *p*-values** |
| Weekly sleep duration [d] | | | | | | | | | | | | |
| Short | 0.96 (0.75,1.24) | **1.44 (1.19,1.74)‡** | 1.01 (0.86,1.20) | 1.15 (0.91,1.46) | **2.33 (1.18,4.58)‡** | **0.0075** | **2.04 (1.37,3.02)‡** | **1.52 (1.01,2.30)‡** | 1.38 (0.96,1.99) | 1.23 (0.73,2.05) | 0.55 (0.14,2.13) | 0.2669 |
| Recommended | 1.00 (Ref) | 1.00 (Ref) | 1.00 (Ref) | 1.00 (Ref) | 1.00 (Ref) | --- | 1.00 (Ref) | 1.00 (Ref) | 1.00 (Ref) | 1.00 (Ref) | 1.00 (Ref) | --- |
| Long | 1.14 (0.90,1.43) | **1.24 (1.03,1.48)‡** | **1.16 (1.00,1.35)‡** | **1.33 (1.12,1.58)‡** | **3.23 (1.75,5.95)‡** | **0.0231** | 1.28 (0.72,2.27) | 1.06 (0.67,1.67) | 1.35 (0.98,1.86) | **1.73 (1.24,2.40)‡** | 1.29 (0.49,3.39) | 0.5237 |
| Long sleep onset latency [e] | | | | | | | | | | | | |
| Yes | **1.71 (1.40,2.09)‡** | **1.63 (1.38,1.93)‡** | **2.02 (1.79,2.28)‡** | **1.74 (1.46,2.07)‡** | 1.24 (0.58,2.64) | 0.1880 | 1.37 (0.78,2.38) | **2.17 (1.61,2.93)‡** | **1.67 (1.21,2.31)‡** | **2.06 (1.48,2.89)‡** | **1.97 (1.16,3.36)‡** | 0.5585 |
| No | 1.00 (Ref) | 1.00 (Ref) | 1.00 (Ref) | 1.00 (Ref) | 1.00 (Ref) | --- | 1.00 (Ref) | 1.00 (Ref) | 1.00 (Ref) | 1.00 (Ref) | 1.00 (Ref) | --- |
| Poor sleep maintenance [f] | | | | | | | | | | | | |
| Yes | **1.52 (1.42,1.62)‡** | **1.40 (1.31,1.49)‡** | **1.43 (1.36,1.51)‡** | **1.39 (1.30,1.48)‡** | **1.63 (1.16,2.29)‡** | 0.2606 | 1.06 (0.81,1.39) | **1.41 (1.23,1.62)‡** | **1.23 (1.06,1.43)‡** | 1.16 (0.97,1.38) | **1.70 (1.23,2.34)‡** | 0.0770 |
| No | 1.00 (Ref) | 1.00 (Ref) | 1.00 (Ref) | 1.00 (Ref) | 1.00 (Ref) | --- | 1.00 (Ref) | 1.00 (Ref) | 1.00 (Ref) | 1.00 (Ref) | 1.00 (Ref) | --- |
| Insomnia symptoms [g] | | | | | | | | | | | | |
| Yes | **1.49 (1.40,1.58)‡** | **1.39 (1.31,1.47)‡** | **1.42 (1.35,1.49)‡** | **1.40 (1.33,1.48)‡** | **1.57 (1.17,2.12)‡** | 0.4580 | 1.04 (0.81,1.35) | **1.37 (1.20,1.56)‡** | **1.25 (1.09,1.43)‡** | 1.13 (0.96,1.34) | **1.63 (1.24,2.16)‡** | 0.0729 |
| No | 1.00 (Ref) | 1.00 (Ref) | 1.00 (Ref) | 1.00 (Ref) | 1.00 (Ref) | --- | 1.00 (Ref) | 1.00 (Ref) | 1.00 (Ref) | 1.00 (Ref) | 1.00 (Ref) | --- |
| Sleep medication use [h] | | | | | | | | | | | | |
| Yes | **1.26 (1.03,1.53)‡** | 1.10 (0.94,1.28) | **1.27 (1.12,1.43)‡** | 1.09 (0.93,1.27) | 1.24 (0.63,2.44) | 0.4515 | **1.78 (1.21,2.62)‡** | **1.49 (1.11,2.01)‡** | 1.20 (0.89,1.63) | 1.01 (0.69,1.47) | **1.91 (1.13,3.23)‡** | 0.1408 |
| No | 1.00 (Ref) | 1.00 (Ref) | 1.00 (Ref) | 1.00 (Ref) | 1.00 (Ref) | --- | 1.00 (Ref) | 1.00 (Ref) | 1.00 (Ref) | 1.00 (Ref) | 1.00 (Ref) | --- |
| Daytime dysfunction [i] | | | | | | | | | | | | |
| Yes | --- | --- | --- | --- | --- | --- | --- | --- | --- | --- | --- | --- |
| No | 1.00 (Ref) | 1.00 (Ref) | 1.00 (Ref) | 1.00 (Ref) | 1.00 (Ref) | --- | 1.00 (Ref) | 1.00 (Ref) | 1.00 (Ref) | 1.00 (Ref) | 1.00 (Ref) | --- |
| Healthcare professional diagnosed sleep apnea [j] | | | | | | | | | | | | |
| Yes | 1.22 (0.92,1.61) | 1.11 (0.89,1.39) | **1.26 (1.06,1.49)‡** | 0.98 (0.78,1.24) | **3.86 (1.13,13.20)‡** | 0.1482 | 1.61 (0.92,2.81) | 0.90 (0.49,1.66) | **1.91 (1.40,2.61)‡** | **1.75 (1.17,2.61)‡** | **5.52 (1.95,15.59)‡** | **0.0467** |
| No | 1.00 (Ref) | 1.00 (Ref) | 1.00 (Ref) | 1.00 (Ref) | 1.00 (Ref) | --- | 1.00 (Ref) | 1.00 (Ref) | 1.00 (Ref) | 1.00 (Ref) | 1.00 (Ref) | --- |

Note: N = 418 (1.8%) participants who reported both 'too hot' and 'too cold' temperatures were not included in this sensitivity analysis.

Models are adjusted age (continuous), age$^2$, race and ethnicity (Hispanic/Latina, non-Hispanic Black, non-Hispanic White), annual household income (<\$20,000 - \$49,999, \$50,000 - \$99,999, ≥\$100,000), marital status (married or living as though married, divorced/widowed/separated/never married), body mass index (BMI: underweight, recommended, overweight, obesity), and menopausal status (premenopausal, postmenopausal).

[a] In models for weekly sleep duration, sleep medication use, daytime dysfunction, and healthcare professional diagnosed sleep apnea, reasons for trouble sleeping for any reason other than feeling too hot or feeling too cold ≥ 3 times per week in the past month include: unable to fall asleep within 30 minutes, waking up in the middle of the night or early morning, waking up to use the bathroom, cannot breathe comfortably, coughing or snoring loudly, having bad dreams, having pain, or other non-specified reasons (n = 23,853). In models for long sleep onset latency, poor sleep maintenance, and insomnia symptoms, reasons for trouble sleeping for any reason other than feeling too hot or feeling too cold ≥ 3 times per week in the past month include waking up to use the bathroom, cannot breathe comfortably, coughing or snoring loudly, having bad dreams, having pain, or other non-specified reasons (n = 21,641).

[b] Too hot' is defined as self-reported trouble sleeping due to feeling too hot ≥ 3 times per week vs. trouble sleeping due to feeling too hot < 3 times per week and trouble sleeping due to feeling too cold < 3 times per week.

[c] 'Too cold' is defined as self-reported trouble sleeping due to feeling too cold ≥ 3 times per week vs. trouble sleeping due to feeling too hot < 3 times per week and trouble sleeping due to feeling too cold < 3 times per week.
[d] Sleep duration is based on reported bed and wake times or reported average sleep duration. Participants who reported ≤ 2 or ≥ 23 hours of sleep are excluded. Short: <7 hours; Recommended: 7-9 hours; Long ≥ 9 hours.
[e] Long sleep onset latency is defined as not falling asleep within 30 minutes at least three times a week during the past month.
[f] Poor sleep maintenance is defined as waking up in the middle of the night or early morning at least three times a week during the past month.
[g] Insomnia symptoms is defined as long sleep onset latency or poor sleep maintenance.
[h] Sleep medication use is defined as 'taking medicine (prescription or over the counter) to help you sleep' at least three times a week during the past month.
[i] Daytime dysfunction is defined as having trouble staying awake while driving, eating, or engaging in social activity at least three times a week during the past month.
[j] Healthcare provider diagnosed sleep apnea is defined as a current doctor or other health professional diagnosis of sleep apnea.
[ł] Remained significant after false discovery rate correction. False discovery rate-corrected *p*-value was considered statistically significant at the α = 0.05 level.

**Supplemental Table 19B. Relative excess risk due to interaction (RERI) between perceived temperature extremes and region of residence, among participants who reported trouble sleeping for any reason [a], Sister Study, 2017-2019**

| | Indoor temperature: Too hot [b] | | | | | Indoor temperature: Too cold [c] | | | | |
|---|---|---|---|---|---|---|---|---|---|---|
| | **Northeast** | **Midwest** | **South** | **West** | **Puerto Rico** | **Northeast** | **Midwest** | **South** | **West** | **Puerto Rico** |
| Weekly sleep duration [d] | | | | | | | | | | |
| Short | -0.06 (-0.40, 0.28) | **0.39 (0.11, 0.67)** | | 0.13 (-0.17, 0.43) | 1.29 (-0.14, 2.72) | 0.89 (-0.19, 1.97) | 0.10 (-0.66, 0.86) | | -0.17 (-0.95, 0.61) | -0.81 (-1.72, 0.10) |
| Recommended | --- | --- | 1.00 (Ref) | --- | --- | --- | --- | 1.00 (Ref) | --- | --- |
| Long | -0.03 (-0.33, 0.27) | 0.05 (-0.20, 0.30) | | 0.15 (-0.12, 0.42) | **1.22 (0.18, 2.26)** | -0.08 (-0.90, 0.74) | -0.30 (-0.91, 0.31) | | 0.34 (-0.34, 1.02) | -0.17 (-1.03, 0.69) |
| Long sleep onset latency [e] | | | | | | | | | | |
| Yes | -0.31 (-0.69, 0.07) | **-0.43 (-0.75, -0.11)** | 1.00 (Ref) | -0.33 (-0.67, 0.01) | -0.79 (-1.69, 0.11) | -0.31 (-1.22, 0.60) | 0.41 (-0.38, 1.20) | 1.00 (Ref) | 0.32 (-0.51, 1.15) | 0.34 (-0.78, 1.46) |
| No | --- | --- | | --- | --- | --- | --- | | --- | --- |
| Poor sleep maintenance [f] | | | | | | | | | | |
| Yes | 0.10 (0.00, 0.20) | -0.04 (-0.13, 0.05) | 1.00 (Ref) | -0.03 (-0.12, 0.06) | 0.15 (-0.29, 0.59) | -0.17 (-0.51, 0.17) | 0.17 (-0.09, 0.43) | 1.00 (Ref) | -0.07 (-0.34, 0.20) | 0.40 (-0.05, 0.85) |
| No | --- | --- | | --- | --- | --- | --- | | --- | --- |
| Insomnia symptoms [g] | | | | | | | | | | |
| Yes | 0.08 (-0.01, 0.17) | -0.04 (-0.12, 0.04) | 1.00 (Ref) | -0.01 (-0.09, 0.07) | 0.10 (-0.26, 0.46) | -0.21 (-0.52, 0.10) | 0.11 (-0.12, 0.34) | 1.00 (Ref) | -0.12 (-0.37,0.13) | 0.33 (-0.04, 0.70) |
| No | | | | | | | | | | |
| Sleep medication use [h] | | | | | | | | | | |
| Yes | -0.06 (-0.31, 0.19) | -0.18 (-0.39, 0.03) | 1.00 (Ref) | -0.18 (-0.40, 0.04) | 0.06 (-1.07, 1.19) | 0.45 (-0.22, 1.12) | 0.25 (-0.29, 0.79) | 1.00 (Ref) | -0.20 (-0.73, 0.33) | 1.06 (-0.26, 2.38) |
| No | --- | --- | | --- | --- | --- | --- | | --- | --- |
| Daytime dysfunction [i] | | | | | | | | | | |
| Yes | --- | --- | 1.00 (Ref) | --- | --- | --- | --- | 1.00 (Ref) | --- | --- |
| No | --- | --- | | --- | --- | --- | --- | | --- | --- |
| Healthcare professional diagnosed sleep apnea [j] | | | | | | | | | | |
| Yes | -0.06 (-0.42, 0.30) | -0.15 (-0.45, 0.15) | 1.00 (Ref) | -0.28 (-0.62, 0.06) | 0.78 (-0.67, 2.23) | -0.37 (-1.36, 0.62) | **-1.01 (-1.80, -0.22)** | 1.00 (Ref) | -0.00 (-1.01, 1.01) | 0.76 (-0.93, 2.45) |
| No | --- | --- | | --- | --- | --- | --- | | --- | --- |

Note: N = 418 (1.8%) participants who reported both 'too hot' and 'too cold' temperatures were not included in this sensitivity analysis.
RERI = 0: no additive interaction; RERI >0: positive additive interaction; RERI <0: negative additive interaction.

Models are adjusted for age (continuous), age$^2$, race and ethnicity (Hispanic/Latina, non-Hispanic Black, non-Hispanic White), annual household income (<$20,000 - $49,999, $50,000 - $99,999, ≥$100,000), marital status (married or living as though married, divorced/widowed/separated/never married), body mass index (BMI: underweight, recommended, overweight, obesity), and menopausal status (premenopausal, postmenopausal).

[a] In models for weekly sleep duration, sleep medication use, daytime dysfunction, and healthcare professional diagnosed sleep apnea, reasons for trouble sleeping for any reason other than feeling too hot or feeling too cold ≥ 3 times per week in the past month include: unable to fall asleep within 30 minutes, waking up in the middle of the night or early morning, waking up to use the bathroom, cannot breathe comfortably, coughing or snoring loudly, having bad dreams, having pain, or other non-specified reasons (n = 23,853). In models for long sleep onset latency, poor sleep maintenance, and insomnia symptoms, reasons for trouble sleeping for any reason other than feeling too hot or feeling too cold ≥ 3 times per week in the past month include waking up to use the bathroom, cannot breathe comfortably, coughing or snoring loudly, having bad dreams, having pain, or other non-specified reasons (n = 21,641).
[b] Too hot' is defined as self-reported trouble sleeping due to feeling too hot ≥ 3 times per week vs. trouble sleeping due to feeling too hot < 3 times per week and trouble sleeping due to feeling too cold < 3 times per week.
[c] 'Too cold' is defined as self-reported trouble sleeping due to feeling too cold ≥ 3 times per week vs. trouble sleeping due to feeling too hot < 3 times per week and trouble sleeping due to feeling too cold < 3 times per week.
[d] Sleep duration is based on reported bed and wake times or reported average sleep duration. Participants who reported ≤ 2 or ≥ 23 hours of sleep are excluded. Short: <7 hours; Recommended: 7-9 hours; Long ≥ 9 hours.
[e] Long sleep onset latency is defined as not falling asleep within 30 minutes at least three times a week during the past month.
[f] Poor sleep maintenance is defined as waking up in the middle of the night or early morning at least three times a week during the past month.
[g] Insomnia symptoms is defined as long sleep onset latency or poor sleep maintenance.
[h] Sleep medication use is defined as 'taking medicine (prescription or over the counter) to help you sleep' at least three times a week during the past month.
[i] Daytime dysfunction is defined as having trouble staying awake while driving, eating, or engaging in social activity at least three times a week during the past month.

[j] Healthcare provider diagnosed sleep apnea is defined as a current doctor or other health professional diagnosis of sleep apnea.

**Supplemental Table 20A. Prevalence ratios (95% confidence intervals) for associations between perceived indoor temperature extremes and sleep health dimensions by menopausal status, among participants who reported trouble sleeping for any reason [a], Sister Study, 2017-2019**

| | Indoor temperature: Too hot [b] | | | Indoor temperature: Too cold [c] | | |
|---|---|---|---|---|---|---|
| | **Premenopausal** | **Postmenopausal** | **Wald *p*-values** | **Premenopausal** | **Postmenopausal** | **Wald *p*-values** |
| Weekly sleep duration [d] | | | | | | |
| Short | 1.13 (0.75,1.70) | **1.14 (1.03,1.27)‡** | 0.9567 | 1.55 (0.74,3.22) | **1.42 (1.14,1.77)‡** | 0.8276 |
| Recommended | 1.0 (Ref) | 1.0 (Ref) | --- | 1.0 (Ref) | 1.0 (Ref) | --- |
| Long | 1.15 (0.72,1.83) | **1.25 (1.13,1.37)‡** | 0.7407 | 1.35 (0.47,3.84) | **1.36 (1.12,1.66)‡** | 0.9855 |
| Long sleep onset latency [e] | | | | | | |
| Yes | **2.16 (1.45,3.21)‡** | **1.79 (1.65,1.95)‡** | 0.3677 | 0.94 (0.25,3.60) | **1.90 (1.61,2.25)‡** | 0.3085 |
| No | 1.0 (Ref) | 1.0 (Ref) | --- | 1.0 (Ref) | 1.0 (Ref) | --- |
| Poor sleep maintenance [f] | | | | | | |
| Yes | **1.45 (1.25,1.68)‡** | **1.43 (1.38,1.47)‡** | 0.8261 | **1.40 (1.00,1.96)‡** | **1.25 (1.15,1.36)‡** | 0.5299 |
| No | 1.0 (Ref) | 1.0 (Ref) | --- | 1.0 (Ref) | 1.0 (Ref) | --- |
| Insomnia symptoms [g] | | | | | | |
| Yes | **1.32 (1.19,1.47)‡** | **1.35 (1.32,1.39)‡** | 0.6549 | 1.18 (0.88,1.59) | **1.21 (1.12,1.29)‡** | 0.9033 |
| No | 1.0 (Ref) | 1.0 (Ref) | --- | 1.0 (Ref) | 1.0 (Ref) | --- |
| Sleep medication use [h] | | | | | | |
| Yes | 1.03 (0.70,1.50) | **1.19 (1.10,1.28)‡** | 0.4659 | 1.50 (0.71,3.16) | **1.33 (1.13,1.56)‡** | 0.7535 |
| No | 1.0 (Ref) | 1.0 (Ref) | --- | 1.0 (Ref) | 1.0 (Ref) | --- |
| Daytime dysfunction [i] | | | | | | |
| Yes | 0.94 (0.11,8.39) | **1.79 (1.23,2.61)‡** | 0.5693 | **15.61 (2.92,83.52)‡** | **4.43 (2.54,7.74)‡** | 0.1706 |
| No | 1.0 (Ref) | 1.0 (Ref) | --- | 1.0 (Ref) | 1.0 (Ref) | --- |
| Healthcare professional diagnosed sleep apnea [j] | | | | | | |
| Yes | 0.87 (0.47,1.60) | **1.17 (1.04,1.31)‡** | 0.3517 | 0.61 (0.09,4.03) | **1.69 (1.38,2.08)‡** | 0.2919 |
| No | 1.0 (Ref) | 1.0 (Ref) | --- | 1.0 (Ref) | 1.0 (Ref) | --- |

Note: N = 418 (1.8%) participants who reported both 'too hot' and 'too cold' temperatures were not included in this sensitivity analysis.

Models are adjusted for age (continuous), age$^2$, race and ethnicity (Hispanic/Latina, non-Hispanic Black, non-Hispanic White), income (<$20,000 - $49,999, $50,000 - $99,999, ≥$100,000), marital status (married or living as though married, divorced/widowed/separated/never married), region of residence (Northeast, Midwest, South, West, and Puerto Rico), and body mass index (BMI: underweight, recommended, overweight, obesity).

[a] In models for weekly sleep duration, sleep medication use, daytime dysfunction, and healthcare professional diagnosed sleep apnea, reasons for trouble sleeping for any reason other than feeling too hot or feeling too cold ≥ 3 times per week in the past month include: unable to fall asleep within 30 minutes, waking up in the middle of the night or early morning, waking up to use the bathroom, cannot breathe comfortably, coughing or snoring loudly, having bad dreams, having pain, or other non-specified reasons (n = 23,853). In models for long sleep onset latency, poor sleep maintenance, and insomnia symptoms, reasons for trouble sleeping for any reason other than feeling too hot or feeling too cold ≥ 3 times per week in the past month include waking up to use the bathroom, cannot breathe comfortably, coughing or snoring loudly, having bad dreams, having pain, or other non-specified reasons (n = 21,641).
[b] Too hot' is defined as self-reported trouble sleeping due to feeling too hot ≥ 3 times per week vs. trouble sleeping due to feeling too hot < 3 times per week and trouble sleeping due to feeling too cold < 3 times per week.
[c] 'Too cold' is defined as self-reported trouble sleeping due to feeling too cold ≥ 3 times per week vs. trouble sleeping due to feeling too hot < 3 times per week and trouble sleeping due to feeling too cold < 3 times per week.
[d] Sleep duration is based on reported bed and wake times or reported average sleep duration. Participants who reported ≤ 2 or ≥ 23 hours of sleep are excluded. Short: <7 hours; Recommended: 7-9 hours; Long ≥ 9 hours.
[e] Long sleep onset latency is defined as not falling asleep within 30 minutes at least three times a week during the past month.
[f] Poor sleep maintenance is defined as waking up in the middle of the night or early morning at least three times a week during the past month.
[g] Insomnia symptoms is defined as long sleep onset latency or poor sleep maintenance.
[h] Sleep medication use is defined as 'taking medicine (prescription or over the counter) to help you sleep' at least three times a week during the past month.
[i] Daytime dysfunction is defined as having trouble staying awake while driving, eating, or engaging in social activity at least three times a week during the past month.
[j] Healthcare provider diagnosed sleep apnea is defined as a current doctor or other health professional diagnosis of sleep apnea.

[‡] Remained significant after false discovery rate correction. False discovery rate-corrected *p*-value was considered statistically significant at the $\alpha = 0.05$ level.

**Supplemental Table 20B. Relative excess risk due to interaction (RERI) between perceived temperature extremes and menopausal status, among participants who reported trouble sleeping for any reason [a], Sister Study, 2017-2019**

| | Indoor temperature: Too hot [b] | | Indoor temperature: Too cold [c] | |
|---|---|---|---|---|
| | **Premenopausal** | **Postmenopausal** | **Premenopausal** | **Postmenopausal** |
| Weekly sleep duration [d] | | | | |
| Short | | 0.01 (-0.42, 0.44) | | -0.12 (-1.26, 1.02) |
| Recommended | 1.0 (Ref) | --- | 1.0 (Ref) | --- |
| Long | | -0.04 (-0.53, 0.45) | | -0.08 (-1.52, 1.36) |
| Long sleep onset latency [e] | | | | |
| Yes | 1.0 (Ref) | -0.15 (-0.81, 0.51) | 1.0 (Ref) | **1.32 (0.04, 2.60)** |
| No | | --- | | --- |
| Poor sleep maintenance [f] | | | | |
| Yes | 1.0 (Ref) | N/A* | 1.0 (Ref) | -0.14 (-0.58, 0.30) |
| No | | --- | | --- |
| Insomnia symptoms [g] | | | | |
| Yes | 1.0 (Ref) | N/A* | 1.0 (Ref) | -0.07 (-0.48, 0.34) |
| No | | --- | | --- |
| Sleep medication use [h] | | | | |
| Yes | 1.0 (Ref) | 0.16 (-0.20, 0.52) | 1.0 (Ref) | -0.18 (-1.29, 0.93) |
| No | | --- | | --- |
| Daytime dysfunction [i] | | | | |
| Yes | 1.0 (Ref) | 1.21 (-0.97, 3.39) | 1.0 (Ref) | -9.57 (-33.73, 14.59) |
| No | | --- | | --- |
| Healthcare professional diagnosed sleep apnea [j] | | | | |
| Yes | 1.0 (Ref) | 0.33 (-0.14, 0.80) | 1.0 (Ref) | **1.26 (0.08, 2.44)** |
| No | | --- | | --- |

Note: N = 418 (1.8%) participants who reported both 'too hot' and 'too cold' temperatures were not included in this sensitivity analysis.
RERI = 0: no additive interaction; RERI >0: positive additive interaction; RERI <0: negative additive interaction.

Models are adjusted for age (continuous), age$^2$, race and ethnicity (Hispanic/Latina, non-Hispanic Black, non-Hispanic White), income (<$20,000 - $49,999, $50,000 - $99,999, ≥$100,000), marital status (married or living as though married, divorced/widowed/separated/never married), region of residence (Northeast, Midwest, South, West, and Puerto Rico), and body mass index (BMI: underweight, recommended, overweight, obesity).

[a] In models for weekly sleep duration, sleep medication use, daytime dysfunction, and healthcare professional diagnosed sleep apnea, reasons for trouble sleeping for any reason other than feeling too hot or feeling too cold ≥ 3 times per week in the past month include: unable to fall asleep within 30 minutes, waking up in the middle of the night or early morning, waking up to use the bathroom, cannot breathe comfortably, coughing or snoring loudly, having bad dreams, having pain, or other non-specified reasons (n = 23,853). In models for long sleep onset latency, poor sleep maintenance, and insomnia symptoms, reasons for trouble sleeping for any reason other than feeling too hot or feeling too cold ≥ 3 times per week in the past month include waking up to use the bathroom, cannot breathe comfortably, coughing or snoring loudly, having bad dreams, having pain, or other non-specified reasons (n = 21,641).
[b] Too hot' is defined as self-reported trouble sleeping due to feeling too hot ≥ 3 times per week vs. trouble sleeping due to feeling too hot < 3 times per week and trouble sleeping due to feeling too cold < 3 times per week.
[c] 'Too cold' is defined as self-reported trouble sleeping due to feeling too cold ≥ 3 times per week vs. trouble sleeping due to feeling too hot < 3 times per week and trouble sleeping due to feeling too cold < 3 times per week.

[d] Sleep duration is based on reported bed and wake times or reported average sleep duration. Participants who reported ≤ 2 or ≥ 23 hours of sleep are excluded. Short: <7 hours; Recommended: 7-9 hours; Long ≥ 9 hours.
[e] Long sleep onset latency is defined as not falling asleep within 30 minutes at least three times a week during the past month.
[f] Poor sleep maintenance is defined as waking up in the middle of the night or early morning at least three times a week during the past month.
[g] Insomnia symptoms is defined as long sleep onset latency or poor sleep maintenance.
[h] Sleep medication use is defined as 'taking medicine (prescription or over the counter) to help you sleep' at least three times a week during the past month.
[i] Daytime dysfunction is defined as having trouble staying awake while driving, eating, or engaging in social activity at least three times a week during the past month.
[j] Healthcare provider diagnosed sleep apnea is defined as a current doctor or other health professional diagnosis of sleep apnea.
* RERI estimates are invalid because the covariances between ‘too hot’ and postmenopausal status do not fall within the plausible bounds of the 95% CI bound.
